# Re-emergence of cholera in Haiti linked to environmental *V. cholerae* O1 Ogawa strains

**DOI:** 10.1101/2022.11.21.22282526

**Authors:** Carla N. Mavian, Massimiliano S. Tagliamonte, Meer T. Alam, S. Nazmus Sakib, Melanie N. Cash, Alberto Riva, V. Madsen Beau De Rochars, Vanessa Rouzier, Jean William Pape, J. Glenn Morris, Marco Salemi, Afsar Ali

## Abstract

**BACKGROUND:** On September 25^th^, 2022, cholera re-emerged in Haiti.

**OBJECTIVES/METHODS:** Toxigenic *Vibrio cholerae* O1 Ogawa were isolated on October 3^rd^ & 4^th^, 2022, from cholera case patients in Port-au-Prince. The two new genomes were compared with genomes from 2,129 *V. cholerae* O1 isolated worldwide, including 292 Haitian strains from 2010-2018.

**RESULTS:** Phylogenies conclusively show the 2022 strains clustering within the Haitian monophyletic clade dating back to the 2010 outbreak. Strains shared a most recent common ancestor with a 2018 Haitian Ogawa strain isolated from the aquatic ecosystem, and cluster with the Ogawa clade that was circulating in 2015-2016.

**CONCLUSIONS:** Re-emergence of cholera in Haiti is the likely result of a spill-over event at the aquatic-human interface related to persistence of *V. cholerae* O1 in the environment.

**One-Sentence Summary:** We analyzed the full genome of two *V. cholerae* strains isolated from Haitian patients infected during the early days of the current 2022 epidemic, with data indicating that they originated from strains that have been circulating undetected at sub-epidemic levels in the aquatic environment.

## Main Text

Cholera was detected in Haiti for the first time in over 100 years in October of 2010. This followed the major earthquake that occurred in January 2010, which resulted in almost complete destruction of the nation’s public health infrastructure. The 2010 epidemic appears to have been caused by a toxigenic *Vibrio cholerae* O1 strain originally introduced into Haiti by peace-keeping troops from Nepal, through contamination of water in the Artibonite River by sewage outflows from the camp used by the peace-keeping contingent (*1, 2*). Initial transmission of the disease was associated with exposure to river water (*3*), and subsequent work by our group has underscored the critical role played by the aquatic environment in transmission and evolution of epidemic strains in Haiti (*4-7*). Between October 2010 and February 2019 this epidemic resulted in over 820,000 reported cholera cases and close to 10,000 deaths (*8*).

Despite ongoing surveillance, no clinical cholera cases were reported in Haiti between February 2019 and the beginning of September 2022, leading to the assumption that cholera had been eradicated. However, in September of 2022 cholera cases were again identified in the Port-au-Prince region, with toxigenic *V. cholerae* O1 serotype Ogawa isolated from case patients. Between September 25^th^ and November 16^th^, 2022, the Haiti Ministry of Public Health and Population (MSPP) reported 807 laboratory-confirmed cases of toxigenic *V. cholerae* O1 Ogawa serotype with 9,317 suspected cases and 8146 hospitalizations, with cases concentrated in Port-au-Prince, Carrefour and Cité Soleil in the Ouest Department; 35% of cases were in children aged nine years or younger (*9*).

Two strains (designated as VCN3833 and VCN3834) isolated from adult patients seen at the GHESKIO Cholera Treatment Center in Port-au-Prince on October 3 and 4, respectively, were further evaluated at the University of Florida Emerging Pathogens Institute; strains were de-identified, and their use was approved by the University of Florida IRB. Both strains were serotype Ogawa and carried the CTX cholera toxin genes, as well as other key genes associated with cholera pathogenicity and virulence (Supplementary Table S1). Both were susceptible to antimicrobial agents commonly used for treatment of cholera, including doxycycline, ciprofloxacin, and azithromycin (Supplementary Table S2); however, ciprofloxacin susceptibility was borderline, with molecular analysis showing a DNA gyrase mutation in *gyrA* (Ser83Ile) and *parC* (Ser85Leu), as has been previously described in clinical isolates from Haiti (*10, 11*).

We employed next generation sequencing to obtain the full genome of the two 2022 strains, as well as toxigenic *V. cholerae* O1 obtained in Haiti in 2018 (*n*=31)(Supplementary Table S3). VCN3833 and VCN3834 only differ in four SNPs, of which one is a nonsynonymous mutation, i.e., causing a change in amino acid in the encoded protein (Supplementary Table S4). We then explored the phylogenetic relationships of these strains with 1) toxigenic *V. cholerae* O1 full genome sequences that span each epidemic wave from 2010 through 2017 (*n*=261) from *Mavian et al*. (*7*)(Supplemental Table S3); and 2) with 2,096 strains, including strains from Europe (*n*=22), the Americas (*n*=856, including previously available 261 from *Mavian et al*. (*7*)), Asia (*n*=389), the Middle East (*n*=86), and Africa (*n*=743) that were collected between 1937-2022 (Table Supplemental Table S5). Our maximum likelihood phylogeny inferred from the genome-wide single nucleotide polymorphisms’ (SNPs) alignment of 2,131 worldwide strains indicates that the 2022 VCN3833 and VCN3834 isolates cluster within the well supported (bootstrap > 90%) monophyletic Haitian clade that emerged in 2010 and includes all Haitian strains isolated to date (Figure 1). The Haitian clade also comprises a sub-clade of 2013 Mexican strains suggesting a possible spill-over of cholera from Haiti to Mexico.

**Figure 1.**
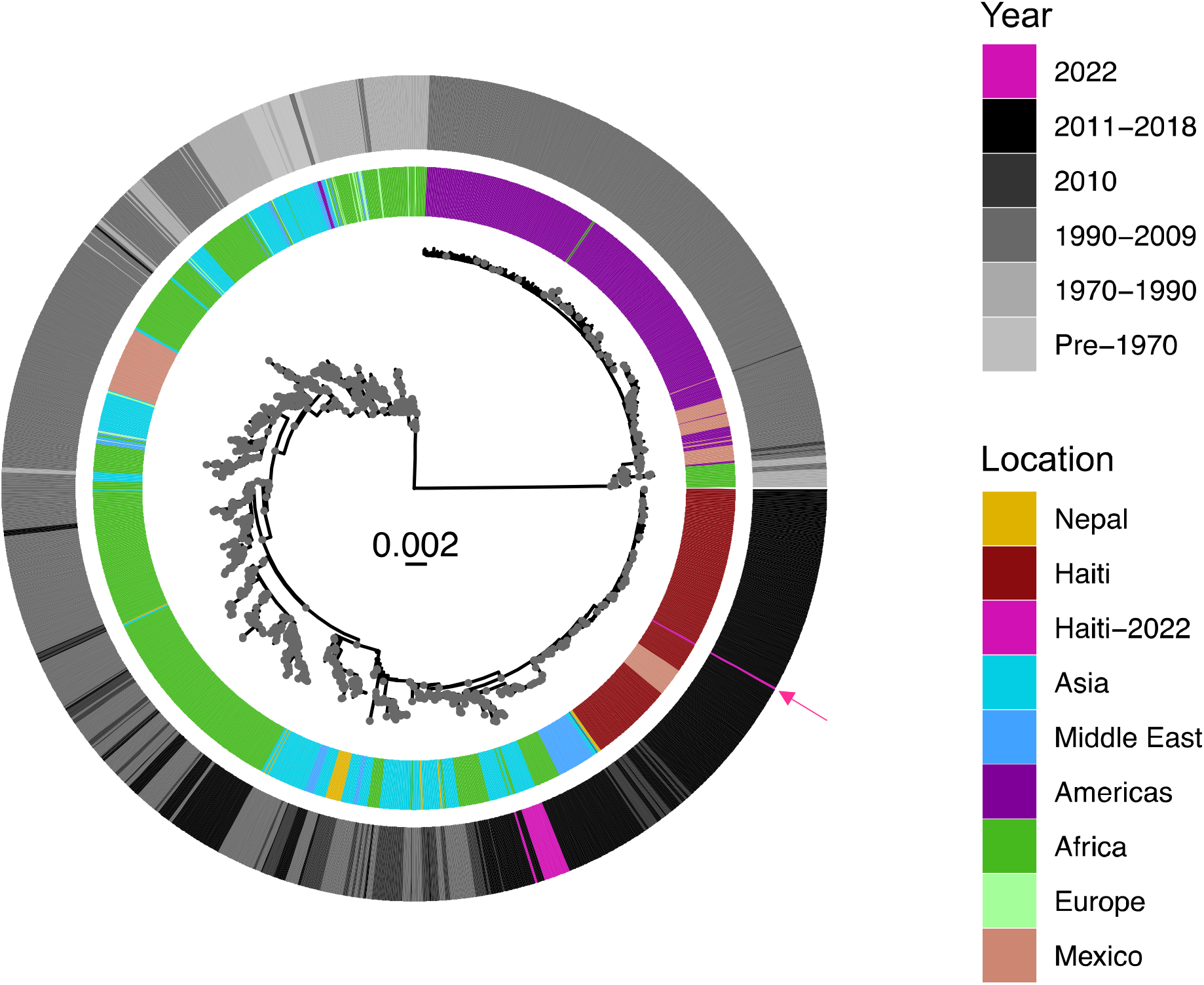
V. cholerae O1 strains from 2022 cluster within the Haitian monophyletic clade. Maximum likelihood phylogeny of global V. cholerae O1 strains obtained between 1937 and 2022 worldwide. The maximum likelihood tree was obtained using IQTREE using best-fit model according to BIC (TVM+F+ASC+R4). Heatmap denote location (geographic region or country) and year (or year range) of collection, following colors as indicated in the legend.

We next investigated the origin of the recent cholera strains (2022) within Haiti and the time to their most recent common ancestor by the Bayesian phylodynamic framework with a molecular clock. Haiti strains collected since 2015 cluster in two highly supported (posterior probability > 0.9) clades: Ogawa and Inaba. The VCN3833 and VCN3834 isolates cluster within the Ogawa clade that contains previous Ogawa strains circulating during the 2015-2016 wave and emerged after a lull period during the epidemic fuelled by the environmental reservoir that was established in the aquatic ecosystem between 2013 and 2014 (*7*).

Our collection included clinical strains collected in 2018. Yet, the strains obtained from the 2022 outbreak (VCN3833 and VCN3834) did not cluster with these clinical isolates (Figure 2A), but directly and with high statistical significance (posterior probability > 0.9) to an environmental Ogawa strain (EnvJ515) sampled in 2018 at a site on the Jacmel Estuary on Haiti’s South-eastern coast (Figure 2A). A total of 20 SNPs differentiate VCN3833 and VCN38334 from EnvJ515 (Table S6), 10 of which are nonsynonymous mutations (three affecting hypothetical proteins) (Figure 2B). Furthermore, at the base of the Ogawa clade is found an environmental strain isolated in 2016 (Env5156) from a river site in Leogane, located on the northern side of the Southern peninsula, with a 2015 environmental isolate from Jacmel Estuary (Env4303), where ENVJ515 was isolated, seeding both the Ogawa and the Inaba clades. The time to the most common ancestor of VCN3833 and VCN38334 was August of 2022 with a high posterior density 95% (HPD) interval of January and October 2022, sharing a common ancestor with EnvJ515 in July 2018 (HPD interval May and July 2018) (Figure 2A).

**Figure 2.**
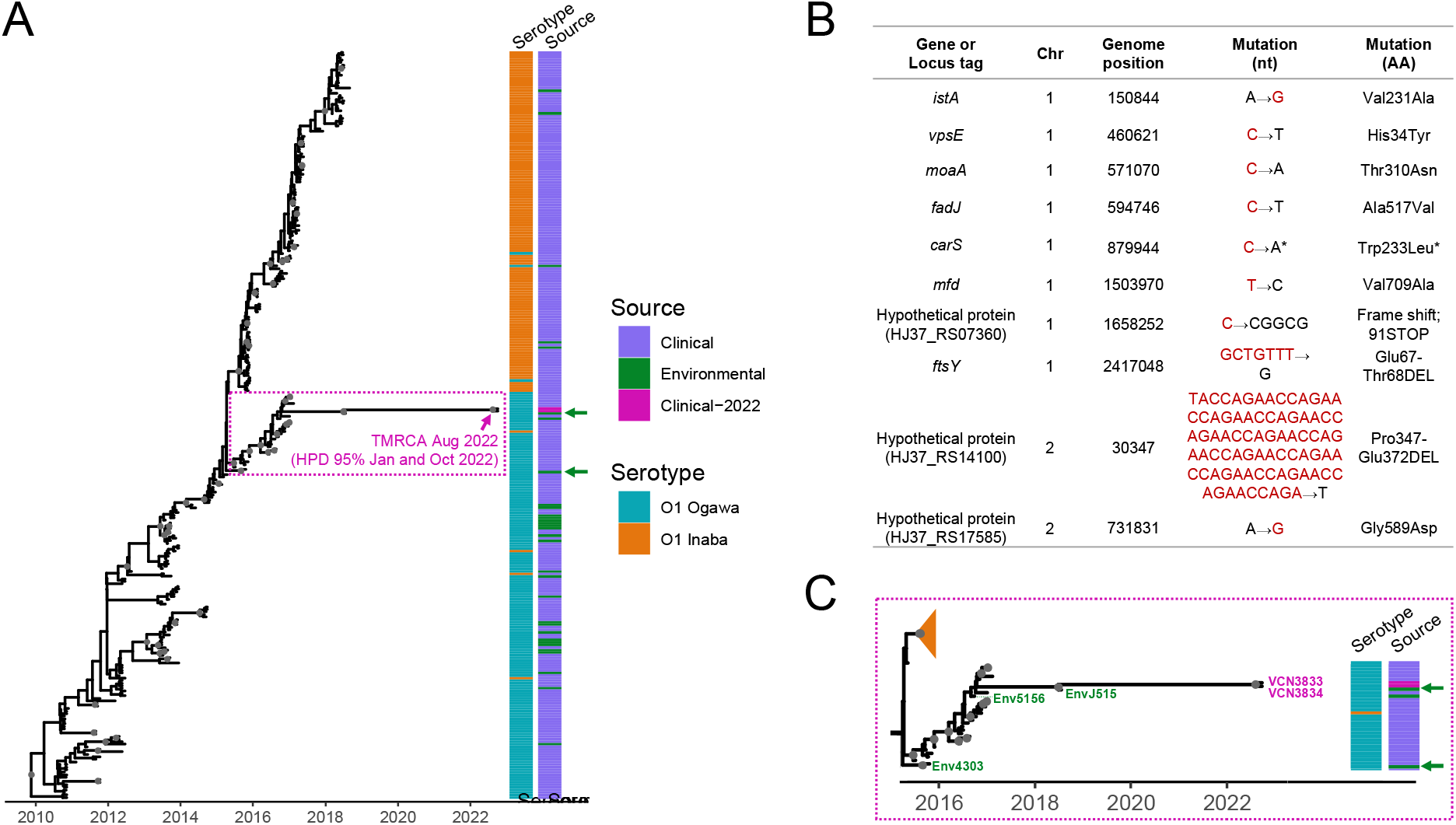
Reemergence of V. cholerae O1 epidemic in Haiti is related to Ogawa strains circulating in humans and in the aquatic ecosystem. (A) MCC time-scaled phylogeny was inferred by enforcing a relaxed clock and Bayesian skyline demographic prior in BEAST v. 1.10.4. Posterior probability was greater than 0.9 is shown with gray circles at nodes. Branch lengths are scaled in time. The time to the most common ancestor (TMRCA) of main divergence events are shown at the nodes. Heatmaps denote source (clinical or environmental) and serotype (O1 Ogawa or Inaba) of the strains. Arrow indicates the position two strains from the 2022 outbreak, VCN3833 and VCN3834, and environmental strains EnvJ515 and Env4303 isolated in 2018 and 2015, respectively. (B) Mutations distinguishing the 2022 strains from EnvJ515 (Ogawa environmental strain collected in 2018). *= mutation only present in VCN3833 (C) Zoom of the phylogenetic tree focusing on the Ogawa clade from which the 2022 *V. cholerae* epidemic strains were derived. Environmental strains are numbered: strain EnvJ515 was isolated in 2018 from Jacmel Estuary, Env5156 was isolated from a river in Leogane in 2016, and Env4303 was isolated in 2015, again, from Jacmel Estuary.

Overall, our analyses demonstrate that strains from the current outbreak are phylogenetically related to the toxigenic *V. cholerae* O1 population that was previously circulating in Haiti and are highly likely to have been derived from an Ogawa 2018 strain isolated from the aquatic ecosystem. Environmental strains isolated in 2015-2016 (including a 2015 isolate from the Jacmel Estuary site), at the base of both Inaba and Ogawa clades, are consistent with establishment of environmental foci which appear to have persisted across a multi-year period. In laboratory settings, we have shown that *V. cholerae* can survive in nutrient-poor environments for over 700 days (*12*). While the ecologic and/or strain factors driving persistence of *V. cholerae* strains within environmental reservoirs remain to be fully elucidated (*13*), subsequent spill-over of the EnvJ515 environmental strain into human populations is clearly plausible. Notably, ten days before the first cholera case was reported September 25^th^, 2022, catastrophic flooding occurred in the island in the aftermath of hurricane Fiona (*13*), potentially facilitating spill-over events igniting a new epidemic wave. It is likely that spread of the epidemic strain was further facilitated by an abrupt interruption of the water supply by DINEPA, the national water company, due to a lack of fuel related to gang warfare and political unrest (*14,15*); this resulted in an inability to provide potable water to shantytown areas of Port-au-Prince, one of the key areas where the epidemic was first identified.

In the current epidemic infection rates appear to be substantially higher in pediatric populations, and, in particular, among children aged 0-9 years; this would be consistent with a lack of immunity to *V. cholerae* in this age group due to the lack of exposure to clinical cases in the preceding 3-4 years. It should also be noted that clinical cases in the 2015-2019 time period were due almost exclusively to *V. cholerae* strains of serotype Inaba; while field-based studies suggest that initial Inaba infections protect against subsequent Ogawa infections (*16*), there may have been issues with cross-protection with the two serotypes, as well as with waning cholera immunity in the general population, leading to increased susceptibility to infection.

From a prevention standpoint, mathematical models developed by our group have indicated that, given the presence of environmental reservoirs, eradication of cholera in Haiti will be difficult without substantive improvements to drinking water and sanitation infrastructure, with clear potential for recurrence of epidemic disease (*7,17,18*). Our modeling has also underscored the potentially critical role that can be played by mass cholera vaccination in controlling epidemics (*18*). While oral killed cholera vaccine has been used successfully in targeted campaigns in Haiti (*19*), efforts have not been made to immunize the entire country or to develop a long-term vaccination strategy (*20*). A major focus of prevention efforts in Haiti has been implementation of rapid response teams, which go to homes of cholera patients and seek to minimize transmission within households with sanitation and chlorination of household water. While these efforts are clearly important, the ongoing risk of recurrent outbreaks linked with environmental reservoirs documented in this study highlights the urgent need for improvements in public health infrastructure, combined, potentially, with periodic mass vaccination campaigns to maintain protective levels of population immunity.

## Materials and Methods

### Sample collection and isolation of V. cholerae

From 2010 to 2022, over 850 toxigenic *V. cholerae* O1 strains were isolated and identified from patients with cholera attending cholera treatment centers, clinics, and hospitals in Haiti (*4, 7, 22, 23*). To isolate toxigenic *V. cholerae* O1 from cholera patients, stool samples were collected and immediately transported to the laboratory in Haiti. The samples were enriched in Alkaline Peptone Water (APW) and inoculated onto TCBS agar plates as described previously (*1-4*). For the isolation and detection of toxigenic *V. cholerae* from environmental samples, water samples were collected monthly from sentinel sites across Haiti and the samples were processed as described above. Each isolate was further characterized using serology and genetic characterization was performed using PCR techniques targeting genes present in toxigenic *V. cholerae* as described previously (*5-6*).

### Antibiogram assay

The susceptibility of two *V. cholerae* strains isolated recently (2022) from Haiti was tested against antimicrobial agents using Kirby-Bauer Disk Diffusion method following Clinical and Laboratory Standards Institute (CLSI) guidelines as described previously (*5*). The antibiotic susceptibility test results of the two strains are shown in Table S2.

### Whole genome mapping and hqSNP calling

In this study, we have analyzed a total of 294 *V. cholerae* O1 strains from Haiti collected from cholera patients’ stool samples and aquatic (water) reservoirs (also referred to as environmental (*7*)): of 294 strains, 261 were from *Mavian et al*. (*7*) (PRJNA510624), 31 new strains were from 2018 and 2 new isolates obtained from the 2022 outbreak (de-identified samples). All new strains (n=33) from 2018 and 2022 were selected for high-quality full genome next generation sequencing (NGS) using in-house protocols previously described (*5-7, 22*) (Table S3). Bacterial genomic DNA extraction was performed as previously described (*7*). Sample library construction was performed using the Nextera XT DNA Library Preparation Kit was performed according to the manufacturer’s instructions (Illumina). Two new 2022 strains (de-identified samples) were obtained from Haiti using UF IRB. Whole genome sequencing on all isolates was executed using the MiSeq Reagent Kit V3 for 600 cycles on the Illumina MiSeq System (Illumina). Raw reads and genome assemblies were downloaded from the NCBI and ENA databases; reads quality assessment and trimming with fastp v. /0.22.0 (*24*); data were analyzed by reference mapping, using the N16961 strain (acc. No. NZ_CP028827.1 and NZ_CP028828.1) as reference for the global dataset and the 2010EL-1786 strain (acc. No. NC_016445.1 and NC_016446.1) for the Haitian clade dataset analysis. The Snippy v 4.6.0 pipeline (*25*) was used to generate synthetic reads from genome assemblies, mapping to the reference genome, and call variants. Variant calling thresholds FreeBayes (*26*) were set as minimum 10x site coverage, minimum mapping quality 60, and minimum 90% base concordance. Individual vcf files were merged using bcftools v.1.15 (*27, 28*). Consensus genomes alignments were scanned for recombination with Gubbins v.3.2.1 (*29*). For the global dataset, the alignment was first split into clusters identified with fastBaps v.1.0.8 (*30*) prior to recombination analysis. Fasta files manipulation was performed with seqkit v.2.0.0 (*31*) and Biostrings v.2.58.0 (*32*); parsimony informative sites for phylogenetic analyses were extracted from consensus genome alignments using Mega-X v.10.0.3 (*33*).

### Phylogenetic and phylodynamic inference

We investigated the phylogenetic relationships of the new outbreak toxigenic *V. cholerae* O1 strains to previous Haitian and international cholera strains. We analyzed two datasets: 1) an Haitian dataset composed of 294 toxigenic *V. cholerae* O1 strains from Haiti: of 294 strains, 261 from *Mavian et al*. (*7*), and 31 strains were from 2018 and 2 strains from the 2022 outbreak (Table S3); 2); and a global dataset encompassing 2,131 strains, including strains from Europe (*n*=22), the Americas (*n*=856), [Americas strains includes 261 from *Mavian et al*. (*7*), and 31 new strains from 2018 and 2 strains from 2022), Asia (*n*=389), the Middle East (*n*=86), and Africa (*n*=743) that were collected between 1937-2022 (Table S5). Samples from the Americas include strains from the outbreak in Argentina in the 90’s and in Mexico from the 90’s up to 2013. Samples from Asia include strains from Bangladesh from 2022 and Nepal from 2010, as well as a wide range of strains from India collected between the 90’s to 2017. The collection of African and Middle Eastern strains includes samples from the recent outbreak in the Democratic Republic of Congo (*34*) and Yemen in 2017 (*35*).

Phylogenetic signal was determined using likelihood mapping test in IQ-TREE (*36*). Maximum likelihood (ML) phylogenetic inferences based on global and local datasets was performed using IQ-TREE (*36*). Temporal signal within the ML phylogeny was estimated by plotting the root-to-tip divergence using TempEst (*37*) (Figure S1). Molecular clock calibration for estimation of time to the most common ancestor (TMRCA) was carried out with BEAST v1.10.4 software package (*38*) enforcing an uncorrelated relaxed molecular clock and a Bayesian Skyline demographic prior as used previously for cholera in Mavian *et al*. (*7*). Phylogenies inferred using strict clock and constant demographic priors gave similar tree topology and time to the most common ancestor (TMRCA) at the root. The maximum clade credibility (MCC) tree was obtained from the posterior distribution of trees with TreeAnnotator v1.8.4 using best burn-in (*38*). The MCC phylogeny was manipulated in R using the package ggtree (*39*) for publishing purposes.

### Additional data

The genomic sequences have been deposited with NCBI under Sequence Read Archive (SRA) under BioProject ID: PRJNA900623. All other data are available in the Supplementary Information. Xml, global maximal likelihood and MCC trees are available at https://github.com/cmavian/CholeraHaiti2022.

## Author contributions

Conceptualization: CNM, JGMJ, MS, AA, VR, JWP

Methodology: CNM, MST, MTA, MNC, AR, SNS

Investigation: CNM, MST, MTA, MS

Visualization: CNM

Funding acquisition: JGMJ

Project administration: CNM, MTA, VR, JWP, JGMJ, MS, AA

Supervision: CNM, JGMJ, MS, AA

Writing – original draft: CNM

Writing – review & editing: CNM, MST, MTA, MNC, VMBDR, VR, JWP, JGMJ, MS, AA

