## Supplementary material for "Re-emergence of cholera in Haiti linked to environmental *V. cholerae* O1 Ogawa strains": Figure S1, Supplemental Table S1, Supplemental Table S2, Supplemental Table S3, Supplemental Table S4, Supplemental Table S5, Supplemental Table S6

**Fig. S1.**

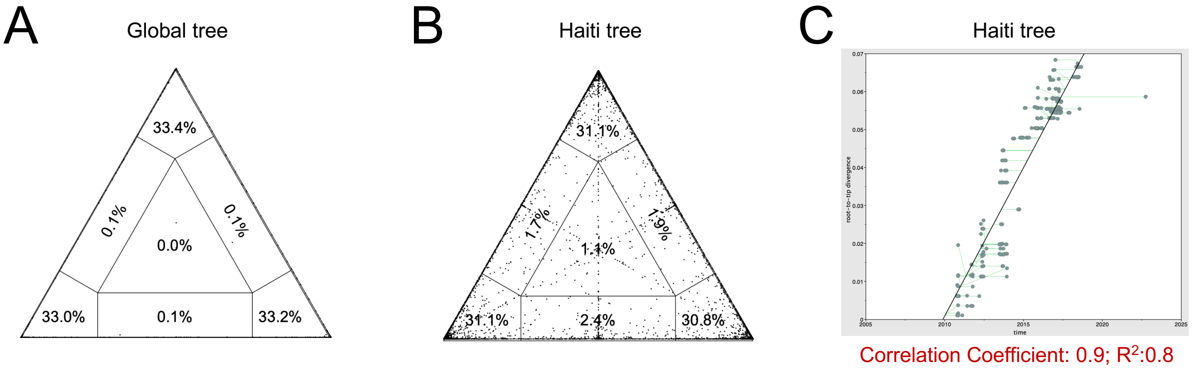


**Phylogenetic quality of the data sets.** (A) Presence of phylogenetic signal was evaluated by likelihood mapping checking for alternative topologies (tips), unresolved quartets (center) and partly resolved quartets (edges) for the data set. (B) Linear regression of root-to-tip genetic distance within the ML phylogeny against sampling time for each taxon: temporal resolution was assessed using the slope of the regression, with positive slope indicating sufficient temporal signal.

**Table S1.**

Major pathogenicity islands and critical genes among 2022 Ogawa *V. cholerae* O1 strains (VCN3383 and VCN3384) and the 2018 Ogawa *V. cholerae* O1 strain (EnvJ515) isolated from aquatic reservoir in Jacmel, Haiti, as compared to the genome of *Vibrio cholerae* O1 2010EL-1786 (reference strain).

| Pathogenicity island/gene | Description | Chromosome | Nucleotide bp (Start - end) | VCN3833 | VCN3834 | EnvJ515 |
|---|---|---|---|---|---|---|
| *tcpA* | Toxin coregulated pilin | I | 367950-368624 | - | - | - |
| *ctxB* | Cholera enterotoxin subunit B | I | 1041238-1041612 | - | - | - |
| *wbeT* | RfbT related protein | I | 2687324-2688226 | - | - | - |
| *ideA* | ICE encoded DNase | I | 152890-153573 | - | - | - |
| *gyrA* | DNA gyrase A subunit | I | 807201-809885 | - | - | - |
| *parC* | Topoisomerase subunit A | I | 2073463-2075748 | - | - | - |
| VPI-1 | Vibrio pathogenicity Island | I | 350786-391625 | 357595(A>G)[a] | - | - |
| VPI-2 | Vibrio pathogenicity Island | I | 1367093-1424290 | NS[b] | NS[b] | NS[b] |
| VSP-1 | Vibrio pathogenicity Island | I | 2600611-2614636 | - | - | - |
| VSP-2 | Vibrio pathogenicity Island | I | 2947311-2961303 | - | - | - |

[a]Mutation found in intergenic region

[b]We detected a nonsynonymous mutation GTGG>CGGC in the annotated gene HJ37_RS06215 (Vch1786_I1284) resulting in the change of amino acids from Proline and Proline to Alanine and Alanine at position 285.

**Table S2.**

Antibiotic susceptibility test perfomed on *V. cholerae* strains recently (2022) isolated from Haiti using standard disc diffusion assay**.**

| Antimicrobial agent with antibiotic content on the disc (μg) | Abbreviated name of each antibiotic | Zone of inhibition in diameter (mm) | | Interpretation[a] |
|---|---|---|---|---|
| | | VCN3833 | VCN3834 | |
| Cephalothin (30) | KF | 18 | 20 | S |
| Ceftriaxone (30) | CRO | 25 | 25 | S |
| Cloramphenicol (30) | C | 15 | 15 | I |
| Cefixime (5) | CFM | 20 | 21 | S |
| Erythromycin (15) | E | 16 | 15 | I |
| Amikacin (30) | AK | 15 | 16 | I |
| Cefotaxime (30) | CTX | 26 | 27 | S |
| Ciprofloxacin (5)[b] | CIP | 22 | 21 | S |
| Azithromycin (15) | AZM | 18 | 19 | S |
| Streptomycin (10) | S | 7 | 7 | R |
| Ceftazidime (30) | CAZ | 23 | 23 | S |
| Sulfisoxazole (1000) | G | 7 | 7 | R |
| Sulphonamide (1000) | SUL | 7 | 7 | R |
| Doxycycline (30) | DO | 20 | 20 | S |
| Ampicillin (10) | AMP | 11 | 12 | R |
| Sulphamethoxazole-trimethoprim (25) | SXT | 7 | 7 | R |
| Nalidixic acid (30) | NA | 7 | 7 | R |
| Tetracycline (30) | TE | 22 | 22 | S |
| Gentamycin (120) | CN | 23 | 21 | S |

[a]S, Susceptible; R, Resistant; I, Intermediate.
[b]Ciprofloxacin, a commonly used antibiotic in Haiti exhibited a borderline sensitivity againt *V. cholerae* strains (VCN3833 and VCN3834) with zone of inhibition (in diameter [mm]) found 22 and 21, respectively compared to standard upper threshold of the antibiotic (≥ 21 mm)

**Table S3.**

Metadata and accession numbers of Haiti toxigenic V. cholerae O1 strains in this study.

| Name | Serogroup | Source | Date | Year |
|---|---|---|---|---|
| SRR771360 | O1 Ogawa | Clinical | 10/19/10 | 2010 |
| SRR773656 | O1 Ogawa | Clinical | 10/19/10 | 2010 |
| SRR770779 | O1 Ogawa | Clinical | 10/20/10 | 2010 |
| SRR771214 | O1 Ogawa | Clinical | 10/20/10 | 2010 |
| SRR771222 | O1 Ogawa | Clinical | 10/20/10 | 2010 |
| SRR771582 | O1 Ogawa | Clinical | 10/20/10 | 2010 |
| SRR771645 | O1 Ogawa | Clinical | 10/21/10 | 2010 |
| SRR772254 | O1 Ogawa | Clinical | 10/21/10 | 2010 |
| SRR773657 | O1 Ogawa | Clinical | 11/3/10 | 2010 |
| SRR773658 | O1 Ogawa | Clinical | 11/3/10 | 2010 |
| AA-142 | O1 Ogawa | Clinical | 11/9/10 | 2010 |
| AA-143 | O1 Ogawa | Clinical | 11/9/10 | 2010 |
| AA-144 | O1 Ogawa | Clinical | 11/9/10 | 2010 |
| AA-145 | O1 Ogawa | Clinical | 11/9/10 | 2010 |
| AA-146 | O1 Ogawa | Clinical | 11/9/10 | 2010 |
| AA-147 | O1 Ogawa | Clinical | 11/9/10 | 2010 |
| AA-148 | O1 Ogawa | Clinical | 11/9/10 | 2010 |
| AA-150 | O1 Ogawa | Clinical | 11/9/10 | 2010 |
| AA-151 | O1 Ogawa | Clinical | 11/9/10 | 2010 |
| SRR772256 | O1 Ogawa | Clinical | 11/9/10 | 2010 |
| SRR773660 | O1 Ogawa | Clinical | 11/27/10 | 2010 |
| SRR772892 | O1 Ogawa | Clinical | 2/12/11 | 2011 |
| SRR772893 | O1 Ogawa | Clinical | 6/6/11 | 2011 |
| SRR773027 | O1 Ogawa | Clinical | 6/9/11 | 2011 |
| SRR773028 | O1 Ogawa | Clinical | 9/2/11 | 2011 |
| SRR773104 | O1 Ogawa | Clinical | 9/2/11 | 2011 |
| SRR773107 | O1 Ogawa | Clinical | 9/2/11 | 2011 |
| SRR773179 | O1 Ogawa | Clinical | 9/12/11 | 2011 |
| SRR773175 | O1 Ogawa | Clinical | 9/20/11 | 2011 |
| SRR773315 | O1 Ogawa | Clinical | 10/11/11 | 2011 |
| SRR773317 | O1 Ogawa | Clinical | 10/11/11 | 2011 |
| SRR773321 | O1 Ogawa | Clinical | 10/12/11 | 2011 |
| SRR773389 | O1 Ogawa | Clinical | 10/12/11 | 2011 |
| SRR773393 | O1 Ogawa | Clinical | 10/12/11 | 2011 |
| SRR773397 | O1 Ogawa | Clinical | 3/13/12 | 2012 |
| HC-07 | O1 Ogawa | Clinical | 4/21/12 | 2012 |
| HC-08 | O1 Ogawa | Clinical | 4/21/12 | 2012 |
| HC-10 | O1 Ogawa | Clinical | 4/21/12 | 2012 |
| HC-11 | O1 Ogawa | Clinical | 5/20/12 | 2012 |
| HC-12 | O1 Ogawa | Clinical | 5/20/12 | 2012 |

| | | | | |
|---|---|---|---|---|
| HC-15 | O1 Ogawa | Clinical | 5/20/12 | 2012 |
| HC-16 | O1 Ogawa | Clinical | 5/20/12 | 2012 |
| HC-17 | O1 Ogawa | Clinical | 5/20/12 | 2012 |
| HC-18 | O1 Ogawa | Clinical | 5/20/12 | 2012 |
| HC-21 | O1 Ogawa | Clinical | 5/20/12 | 2012 |
| HC-22 | O1 Ogawa | Clinical | 5/20/12 | 2012 |
| HC-24 | O1 Ogawa | Clinical | 5/20/12 | 2012 |
| HC-19 | O1 Ogawa | Clinical | 5/22/12 | 2012 |
| env-90 | O1 Ogawa | Environment | 5/24/12 | 2012 |
| env-94 | O1 Ogawa | Environment | 5/24/12 | 2012 |
| HC-31 | O1 Ogawa | Clinical | 6/20/12 | 2012 |
| HC-32 | O1 Ogawa | Clinical | 6/20/12 | 2012 |
| HC-33 | O1 Ogawa | Clinical | 6/20/12 | 2012 |
| HC-34 | O1 Ogawa | Clinical | 6/20/12 | 2012 |
| HC-35 | O1 Inaba | Clinical | 6/20/12 | 2012 |
| env-131 | O1 Ogawa | Environment | 6/22/12 | 2012 |
| env-326 | O1 Ogawa | Environment | 8/27/12 | 2012 |
| SRR8364252 | O1 Ogawa | Environment | 6/27/13 | 2013 |
| SRR8364253 | O1 Ogawa | Environment | 6/27/13 | 2013 |
| SRR8364258 | O1 Ogawa | Environment | 6/27/13 | 2013 |
| SRR8364281 | O1 Ogawa | Clinical | 6/27/13 | 2013 |
| SRR8364340 | O1 Ogawa | Clinical | 7/2/13 | 2013 |
| SRR8364342 | O1 Ogawa | Clinical | 7/11/13 | 2013 |
| SRR8364345 | O1 Ogawa | Clinical | 7/16/13 | 2013 |
| SRR8364346 | O1 Inaba | Clinical | 7/16/13 | 2013 |
| SRR8364369 | O1 Ogawa | Environment | 7/29/13 | 2013 |
| SRR8364371 | O1 Ogawa | Environment | 7/29/13 | 2013 |
| SRR8364372 | O1 Ogawa | Environment | 7/29/13 | 2013 |
| SRR8364339 | O1 Ogawa | Clinical | 8/1/13 | 2013 |
| SRR8364341 | O1 Ogawa | Clinical | 8/1/13 | 2013 |
| SRR8364344 | O1 Ogawa | Clinical | 8/8/13 | 2013 |
| SRR8364337 | O1 Ogawa | Clinical | 8/12/13 | 2013 |
| SRR8364343 | O1 Ogawa | Clinical | 8/22/13 | 2013 |
| SRR8364265 | O1 Ogawa | Clinical | 8/26/13 | 2013 |
| SRR8364289 | O1 Ogawa | Environment | 8/26/13 | 2013 |
| SRR8364262 | O1 Ogawa | Clinical | 8/28/13 | 2013 |
| SRR8364263 | O1 Ogawa | Clinical | 8/29/13 | 2013 |
| SRR8364269 | O1 Ogawa | Clinical | 9/1/13 | 2013 |
| SRR8364270 | O1 Ogawa | Clinical | 9/1/13 | 2013 |
| SRR8364338 | O1 Ogawa | Clinical | 9/4/13 | 2013 |
| SRR8364264 | O1 Ogawa | Clinical | 9/10/13 | 2013 |
| SRR8364266 | O1 Ogawa | Clinical | 9/10/13 | 2013 |
| SRR8364267 | O1 Ogawa | Clinical | 9/16/13 | 2013 |

| | | | | |
|---|---|---|---|---|
| SRR8364268 | O1 Ogawa | Clinical | 9/17/13 | 2013 |
| SRR8364271 | O1 Ogawa | Clinical | 9/19/13 | 2013 |
| SRR8364288 | O1 Ogawa | Environment | 9/23/13 | 2013 |
| SRR8364294 | O1 Ogawa | Environment | 9/23/13 | 2013 |
| SRR8364295 | O1 Ogawa | Environment | 9/23/13 | 2013 |
| SRR8364434 | O1 Ogawa | Environment | 9/23/13 | 2013 |
| SRR8364254 | O1 Ogawa | Environment | 9/25/13 | 2013 |
| SRR8364310 | O1 Ogawa | Clinical | 10/1/13 | 2013 |
| SRR8364298 | O1 Ogawa | Environment | 10/28/13 | 2013 |
| SRR8364299 | O1 Ogawa | Environment | 10/28/13 | 2013 |
| SRR8364430 | O1 Ogawa | Environment | 10/28/13 | 2013 |
| SRR8364437 | O1 Ogawa | Environment | 10/28/13 | 2013 |
| SRR8364309 | O1 Ogawa | Clinical | 10/29/13 | 2013 |
| SRR8364312 | O1 Inaba | Clinical | 11/6/13 | 2013 |
| SRR8364311 | O1 Ogawa | Clinical | 11/15/13 | 2013 |
| SRR8364314 | O1 Ogawa | Clinical | 11/21/13 | 2013 |
| SRR8364257 | O1 Ogawa | Environment | 11/25/13 | 2013 |
| SRR8364436 | O1 Ogawa | Environment | 11/25/13 | 2013 |
| SRR8364313 | O1 Ogawa | Clinical | 11/29/13 | 2013 |
| SRR8364316 | O1 Ogawa | Clinical | 12/4/13 | 2013 |
| SRR8364315 | O1 Ogawa | Clinical | 12/6/13 | 2013 |
| SRR8364318 | O1 Ogawa | Clinical | 12/10/13 | 2013 |
| SRR8364317 | O1 Ogawa | Clinical | 12/15/13 | 2013 |
| SRR8364357 | O1 Ogawa | Clinical | 12/15/13 | 2013 |
| SRR8364358 | O1 Ogawa | Clinical | 12/15/13 | 2013 |
| SRR8364370 | O1 Ogawa | Clinical | 12/16/13 | 2013 |
| SRR8364256 | O1 Ogawa | Environment | 5/1/14 | 2014 |
| SRR8364255 | O1 Ogawa | Environment | 6/1/14 | 2014 |
| SRR8364375 | O1 Ogawa | Clinical | 8/30/14 | 2014 |
| SRR8364373 | O1 Ogawa | Clinical | 9/16/14 | 2014 |
| SRR8364376 | O1 Ogawa | Clinical | 9/16/14 | 2014 |
| SRR8364374 | O1 Ogawa | Clinical | 9/26/14 | 2014 |
| SRR8364377 | O1 Ogawa | Clinical | 9/26/14 | 2014 |
| SRR8364378 | O1 Ogawa | Clinical | 10/16/14 | 2014 |
| SRR8364348 | O1 Ogawa | Clinical | 10/24/14 | 2014 |
| SRR8364347 | O1 Ogawa | Clinical | 11/12/14 | 2014 |
| SRR8364350 | O1 Ogawa | Clinical | 11/27/14 | 2014 |
| SRR8364349 | O1 Ogawa | Clinical | 11/28/14 | 2014 |
| SRR8364352 | O1 Ogawa | Clinical | 11/28/14 | 2014 |
| SRR8364351 | O1 Ogawa | Clinical | 12/17/14 | 2014 |
| SRR8364354 | O1 Ogawa | Clinical | 12/23/14 | 2014 |
| SRR8364353 | O1 Ogawa | Clinical | 1/30/15 | 2015 |
| SRR8364356 | O1 Ogawa | Clinical | 2/19/15 | 2015 |

| | | | | |
|---|---|---|---|---|
| SRR8364355 | O1 Ogawa | Clinical | 3/11/15 | 2015 |
| SRR8364413 | O1 Ogawa | Clinical | 4/14/15 | 2015 |
| SRR8364414 | O1 Ogawa | Clinical | 5/12/15 | 2015 |
| SRR8364415 | O1 Ogawa | Clinical | 7/9/15 | 2015 |
| SRR8364261 | O1 Ogawa | Environment | 9/14/15 | 2015 |
| SRR8364416 | O1 Ogawa | Clinical | 9/25/15 | 2015 |
| SRR8364417 | O1 Ogawa | Clinical | 10/13/15 | 2015 |
| SRR8364418 | O1 Inaba | Clinical | 10/15/15 | 2015 |
| SRR8364419 | O1 Ogawa | Clinical | 10/21/15 | 2015 |
| SRR8364420 | O1 Inaba | Clinical | 10/29/15 | 2015 |
| SRR8364421 | O1 Inaba | Clinical | 10/29/15 | 2015 |
| SRR8364422 | O1 Inaba | Clinical | 10/30/15 | 2015 |
| SRR8364394 | O1 Inaba | Clinical | 11/5/15 | 2015 |
| SRR8364395 | O1 Inaba | Clinical | 11/5/15 | 2015 |
| SRR8364396 | O1 Inaba | Clinical | 11/5/15 | 2015 |
| SRR8364397 | O1 Inaba | Clinical | 11/5/15 | 2015 |
| SRR8364390 | O1 Inaba | Clinical | 11/11/15 | 2015 |
| SRR8364391 | O1 Inaba | Clinical | 11/11/15 | 2015 |
| SRR8364392 | O1 Inaba | Clinical | 11/11/15 | 2015 |
| SRR8364393 | O1 Inaba | Clinical | 11/11/15 | 2015 |
| SRR8364399 | O1 Ogawa | Clinical | 11/11/15 | 2015 |
| SRR8364398 | O1 Inaba | Clinical | 11/12/15 | 2015 |
| SRR8364446 | O1 Inaba | Clinical | 11/12/15 | 2015 |
| SRR8364447 | O1 Inaba | Clinical | 11/12/15 | 2015 |
| SRR8364444 | O1 Inaba | Clinical | 11/13/15 | 2015 |
| SRR8364445 | O1 Inaba | Clinical | 11/19/15 | 2015 |
| SRR8364438 | O1 Inaba | Clinical | 11/20/15 | 2015 |
| SRR8364440 | O1 Inaba | Clinical | 11/20/15 | 2015 |
| SRR8364441 | O1 Inaba | Clinical | 11/20/15 | 2015 |
| SRR8364442 | O1 Inaba | Clinical | 11/20/15 | 2015 |
| SRR8364443 | O1 Inaba | Clinical | 11/20/15 | 2015 |
| SRR8364431 | O1 Inaba | Clinical | 11/25/15 | 2015 |
| SRR8364439 | O1 Inaba | Clinical | 11/25/15 | 2015 |
| SRR8364433 | O1 Inaba | Clinical | 11/26/15 | 2015 |
| SRR8364435 | O1 Inaba | Clinical | 11/26/15 | 2015 |
| SRR8364259 | O1 Inaba | Environment | 11/29/15 | 2015 |
| SRR8364260 | O1 Inaba | Environment | 11/29/15 | 2015 |
| SRR8364425 | O1 Ogawa | Clinical | 12/3/15 | 2015 |
| SRR8364426 | O1 Ogawa | Clinical | 12/3/15 | 2015 |
| SRR8364427 | O1 Inaba | Clinical | 12/3/15 | 2015 |
| SRR8364428 | O1 Inaba | Clinical | 12/3/15 | 2015 |
| SRR8364429 | O1 Inaba | Clinical | 12/3/15 | 2015 |
| SRR8364432 | O1 Inaba | Clinical | 12/3/15 | 2015 |

| | | | | |
|---|---|---|---|---|
| SRR8364321 | O1 Inaba | Clinical | 2/15/16 | 2016 |
| SRR8364322 | O1 Inaba | Clinical | 2/15/16 | 2016 |
| SRR8364323 | O1 Inaba | Clinical | 2/15/16 | 2016 |
| SRR8364324 | O1 Inaba | Clinical | 2/15/16 | 2016 |
| SRR8364424 | O1 Inaba | Clinical | 2/15/16 | 2016 |
| SRR8364327 | O1 Inaba | Clinical | 3/8/16 | 2016 |
| SRR8364379 | O1 Inaba | Clinical | 3/8/16 | 2016 |
| SRR8364319 | O1 Inaba | Clinical | 3/11/16 | 2016 |
| SRR8364320 | O1 Ogawa | Clinical | 6/13/16 | 2016 |
| SRR8364308 | O1 Ogawa | Clinical | 6/22/16 | 2016 |
| SRR8364336 | O1 Ogawa | Clinical | 7/1/16 | 2016 |
| SRR8364401 | O1 Ogawa | Clinical | 7/6/16 | 2016 |
| SRR8364400 | O1 Ogawa | Clinical | 8/10/16 | 2016 |
| SRR8364273 | O1 Ogawa | Clinical | 8/16/16 | 2016 |
| SRR8364272 | O1 Inaba | Clinical | 8/18/16 | 2016 |
| SRR8364279 | O1 Ogawa | Clinical | 8/23/16 | 2016 |
| Env4926 | O1 Ogawa | Environment | 8/29/16 | 2016 |
| SRR8364278 | O1 Inaba | Clinical | 9/6/16 | 2016 |
| SRR8364277 | O1 Ogawa | Clinical | 9/15/16 | 2016 |
| SRR8364276 | O1 Inaba | Clinical | 9/22/16 | 2016 |
| SRR8364423 | O1 Inaba | Clinical | 9/27/16 | 2016 |
| Env5156 | O1 Ogawa | Environment | 9/29/16 | 2016 |
| SRR8364412 | O1 Ogawa | Clinical | 10/11/16 | 2016 |
| SRR8364282 | O1 Inaba | Clinical | 10/18/16 | 2016 |
| SRR8364283 | O1 Inaba | Clinical | 10/25/16 | 2016 |
| SRR8364280 | O1 Ogawa | Clinical | 10/28/16 | 2016 |
| SRR8364286 | O1 Inaba | Clinical | 10/31/16 | 2016 |
| SRR8364287 | O1 Ogawa | Clinical | 11/8/16 | 2016 |
| SRR8364284 | O1 Inaba | Clinical | 11/9/16 | 2016 |
| SRR8364274 | O1 Ogawa | Clinical | 11/21/16 | 2016 |
| SRR8364285 | O1 Inaba | Clinical | 11/21/16 | 2016 |
| SRR8364275 | O1 Inaba | Clinical | 12/5/16 | 2016 |
| SRR8364329 | O1 Ogawa | Clinical | 12/5/16 | 2016 |
| SRR8364328 | O1 Ogawa | Clinical | 12/22/16 | 2016 |
| SRR8364331 | O1 Inaba | Clinical | 12/28/16 | 2016 |
| SRR8364330 | O1 Inaba | Clinical | 1/4/17 | 2017 |
| SRR8364333 | O1 Ogawa | Clinical | 1/5/17 | 2017 |
| SRR8364332 | O1 Ogawa | Clinical | 1/9/17 | 2017 |
| SRR8364335 | O1 Inaba | Clinical | 1/10/17 | 2017 |
| SRR8364334 | O1 Inaba | Clinical | 1/12/17 | 2017 |
| SRR8364326 | O1 Ogawa | Clinical | 1/13/17 | 2017 |
| SRR8364325 | O1 Ogawa | Clinical | 1/16/17 | 2017 |
| SRR8364402 | O1 Inaba | Clinical | 1/18/17 | 2017 |

| | | | | |
|---|---|---|---|---|
| SRR8364403 | O1 Ogawa | Clinical | 1/24/17 | 2017 |
| SRR8364404 | O1 Inaba | Clinical | 1/27/17 | 2017 |
| SRR8364405 | O1 Inaba | Clinical | 2/1/17 | 2017 |
| SRR8364409 | O1 Inaba | Clinical | 2/5/17 | 2017 |
| SRR8364406 | O1 Ogawa | Clinical | 2/13/17 | 2017 |
| SRR8364407 | O1 Inaba | Clinical | 2/20/17 | 2017 |
| SRR8364408 | O1 Inaba | Clinical | 2/20/17 | 2017 |
| SRR8364293 | O1 Inaba | Clinical | 2/21/17 | 2017 |
| SRR8364410 | O1 Inaba | Clinical | 2/21/17 | 2017 |
| SRR8364411 | O1 Inaba | Clinical | 2/21/17 | 2017 |
| SRR8364291 | O1 Inaba | Clinical | 2/24/17 | 2017 |
| SRR8364292 | O1 Inaba | Clinical | 2/24/17 | 2017 |
| SRR8364290 | O1 Inaba | Clinical | 2/27/17 | 2017 |
| SRR8364453 | O1 Inaba | Clinical | 3/2/17 | 2017 |
| SRR8364452 | O1 Inaba | Clinical | 3/3/17 | 2017 |
| SRR8364361 | O1 Inaba | Clinical | 3/5/17 | 2017 |
| SRR8364451 | O1 Inaba | Clinical | 3/6/17 | 2017 |
| SRR8364450 | O1 Inaba | Clinical | 3/14/17 | 2017 |
| SRR8364449 | O1 Inaba | Clinical | 3/17/17 | 2017 |
| SRR8364448 | O1 Inaba | Clinical | 3/23/17 | 2017 |
| SRR8364363 | O1 Inaba | Clinical | 3/28/17 | 2017 |
| SRR8364365 | O1 Inaba | Clinical | 3/28/17 | 2017 |
| SRR8364366 | O1 Inaba | Clinical | 3/28/17 | 2017 |
| SRR8364364 | O1 Inaba | Clinical | 3/30/17 | 2017 |
| SRR8364362 | O1 Inaba | Clinical | 4/4/17 | 2017 |
| SRR8364359 | O1 Inaba | Clinical | 4/7/17 | 2017 |
| SRR8364360 | O1 Inaba | Clinical | 4/11/17 | 2017 |
| SRR8364367 | O1 Inaba | Clinical | 4/13/17 | 2017 |
| SRR8364368 | O1 Inaba | Clinical | 4/17/17 | 2017 |
| SRR8364385 | O1 Inaba | Clinical | 4/17/17 | 2017 |
| SRR8364384 | O1 Inaba | Clinical | 4/18/17 | 2017 |
| SRR8364387 | O1 Inaba | Clinical | 4/20/17 | 2017 |
| SRR8364386 | O1 Inaba | Clinical | 4/21/17 | 2017 |
| SRR8364381 | O1 Inaba | Clinical | 4/24/17 | 2017 |
| SRR8364380 | O1 Inaba | Clinical | 4/27/17 | 2017 |
| SRR8364383 | O1 Inaba | Clinical | 4/28/17 | 2017 |
| SRR8364382 | O1 Inaba | Clinical | 5/5/17 | 2017 |
| SRR8364304 | O1 Inaba | Clinical | 5/17/17 | 2017 |
| SRR8364306 | O1 Inaba | Clinical | 5/17/17 | 2017 |
| SRR8364305 | O1 Inaba | Clinical | 5/18/17 | 2017 |
| SRR8364307 | O1 Inaba | Clinical | 5/18/17 | 2017 |
| SRR8364300 | O1 Inaba | Clinical | 5/19/17 | 2017 |
| SRR8364301 | O1 Inaba | Clinical | 5/20/17 | 2017 |

| | | | | |
|---|---|---|---|---|
| SRR8364302 | O1 Inaba | Clinical | 5/23/17 | 2017 |
| SRR8364303 | O1 Inaba | Clinical | 5/24/17 | 2017 |
| SRR8364296 | O1 Inaba | Clinical | 5/25/17 | 2017 |
| SRR8364297 | O1 Inaba | Clinical | 5/29/17 | 2017 |
| SRR8364389 | O1 Inaba | Clinical | 11/5/17 | 2017 |
| SRR8364388 | O1 Inaba | Clinical | 12/5/17 | 2017 |
| Env6956 | O1 Inaba | Environment | 2/26/18 | 2018 |
| EnvAC5418-5 | O1 Inaba | Environment | 3/12/18 | 2018 |
| VCO1902563 | O1 Inaba | Clinical | 4/27/18 | 2018 |
| VCO1902566 | O1 Inaba | Clinical | 5/3/18 | 2018 |
| VCO1902564 | O1 Inaba | Clinical | 5/4/18 | 2018 |
| VCO1902565 | O1 Inaba | Clinical | 5/4/18 | 2018 |
| VCO1902569 | O1 Inaba | Clinical | 5/6/18 | 2018 |
| VCO1902567 | O1 Inaba | Clinical | 5/8/18 | 2018 |
| VCO1902568 | O1 Inaba | Clinical | 5/9/18 | 2018 |
| VCO1902589 | O1 Inaba | Clinical | 5/11/18 | 2018 |
| VCO1902570 | O1 Inaba | Clinical | 5/22/18 | 2018 |
| VCO1902571 | O1 Inaba | Clinical | 6/1/18 | 2018 |
| VCO1902585 | O1 Inaba | Clinical | 6/1/18 | 2018 |
| VCO1902579 | O1 Inaba | Clinical | 6/6/18 | 2018 |
| VCO1902580 | O1 Inaba | Clinical | 6/7/18 | 2018 |
| VCO1902581 | O1 Inaba | Clinical | 6/8/18 | 2018 |
| VCO1902583 | O1 Inaba | Clinical | 6/10/18 | 2018 |
| VCO1902584 | O1 Inaba | Clinical | 6/11/18 | 2018 |
| VCO1902572 | O1 Inaba | Clinical | 6/19/18 | 2018 |
| VCO1902573 | O1 Inaba | Clinical | 6/20/18 | 2018 |
| VCO1902575 | O1 Inaba | Clinical | 6/20/18 | 2018 |
| VCO1902576 | O1 Inaba | Clinical | 6/20/18 | 2018 |
| VCO1902574 | O1 Inaba | Clinical | 6/21/18 | 2018 |
| VCO1902577 | O1 Inaba | Clinical | 6/21/18 | 2018 |
| VCO1902578 | O1 Inaba | Clinical | 6/30/18 | 2018 |
| VCO1902582 | O1 Inaba | Clinical | 6/30/18 | 2018 |
| VCO1902586 | O1 Inaba | Clinical | 7/3/18 | 2018 |
| VCO1902587 | O1 Inaba | Clinical | 7/4/18 | 2018 |
| VCO1902588 | O1 Inaba | Clinical | 7/5/18 | 2018 |
| EnvJ515 | O1 Ogawa | Environment | 7/23/18 | 2018 |
| VCO1902590 | O1 Inaba | Clinical | 9/1/18 | 2018 |
| VCN3833 | O1 Ogawa | Clinical-2022 | 10/3/22 | 2022 |
| VCN3834 | O1 Ogawa | Clinical-2022 | 10/4/22 | 2022 |

**Table S4.**

List of SNPs that differentiate VCN3833 and VCN38334, as compared to the genome of *Vibrio cholerae* O1 2010EL-1786 (reference strain).

| Gene | Chr | Genome position | Mutation Type | Mutation (AA) | Reference allele | Alternative allele | VCN3833 | VCN3834 |
|---|---|---|---|---|---|---|---|---|
| - | 1 | 62877 | intergenic | | A | G | 1 | 0 |
| carS | 1 | 879944 | nonsynonymous | Trp233 Leu | C | A | 1 | 0 |
| - | 1 | 1042459 | intergenic | | GAAATCAA | G | 0 | 1 |
| - | 2 | 52354 | intergenic | | C | CA | 1 | 0 |

**Table S5.**

Metadata and accession numbers of all (Haiti and global) toxigenic V. cholerae O1 strains in this

| Name | Country | Region | Year | # SRA/ENA |
|---|---|---|---|---|
| 2009V_1046 | Pakistan | Asia | 2009 | GCA_000237405.2 |
| 2009V_1085 | India | Asia | 2009 | GCA_000237425.2 |
| 2009V_1096 | India | Asia | 2009 | GCA_000237445.2 |
| 2009V_1116 | Pakistan | Asia | 2009 | GCA_000237465.2 |
| 2009V_1131 | India | Asia | 2009 | GCA_000237485.2 |
| A1552 | United States | Americas | 1992 | GCF_002892855_1 |
| AA-142 | Haiti | Americas | 2010 | JSDO00000000 |
| AA-143 | Haiti | Americas | 2010 | JSCC00000000 |
| AA-144 | Haiti | Americas | 2010 | JSSV00000000 |
| AA-145 | Haiti | Americas | 2010 | JSSW00000000 |
| AA-146 | Haiti | Americas | 2010 | JSSX00000000 |
| AA-147 | Haiti | Americas | 2010 | JSSY00000000 |
| AA-148 | Haiti | Americas | 2010 | JSSZ00000000 |
| AA-150 | Haiti | Americas | 2010 | JSTA00000000 |
| AA-151 | Haiti | Americas | 2010 | JSTB00000000 |
| AC5418-5 | Haiti | Americas | 2018 | AC5418-5 |
| B33 | Mozambique | Africa | 2004 | ACHZ00000000 |
| CIRS101 | Bangladesh | Asia | 2002 | ACVW00000000 |
| DRR394994 | Bangladesh | Asia | 2022 | DRR394994 |
| DRR394995 | Bangladesh | Asia | 2022 | DRR394995 |
| DRR394996 | Bangladesh | Asia | 2022 | DRR394996 |
| DRR394997 | Bangladesh | Asia | 2022 | DRR394997 |
| DRR394998 | Bangladesh | Asia | 2022 | DRR394998 |
| DRR394999 | Bangladesh | Asia | 2022 | DRR394999 |
| DRR395000 | Bangladesh | Asia | 2022 | DRR395000 |
| DRR395001 | Bangladesh | Asia | 2022 | DRR395001 |
| DRR395002 | Bangladesh | Asia | 2022 | DRR395002 |
| DRR395003 | Bangladesh | Asia | 2022 | DRR395003 |
| DRR395004 | Bangladesh | Asia | 2022 | DRR395004 |
| DRR395005 | Bangladesh | Asia | 2022 | DRR395005 |
| DRR395006 | Bangladesh | Asia | 2022 | DRR395006 |
| DRR395007 | Bangladesh | Asia | 2022 | DRR395007 |
| DRR395008 | Bangladesh | Asia | 2022 | DRR395008 |
| DRR395009 | Bangladesh | Asia | 2022 | DRR395009 |
| DRR395010 | Bangladesh | Asia | 2022 | DRR395010 |
| DRR395011 | Bangladesh | Asia | 2022 | DRR395011 |
| DRR395012 | Bangladesh | Asia | 2022 | DRR395012 |
| DRR395013 | Bangladesh | Asia | 2022 | DRR395013 |
| DRR395014 | Bangladesh | Asia | 2022 | DRR395014 |
| DRR395015 | Bangladesh | Asia | 2022 | DRR395015 |

| | | | | |
|---|---|---|---|---|
| env-131 | Haiti | Americas | 2012 | JSTC01000061.1 |
| env-326 | Haiti | Americas | 2012 | JSTG00000000 |
| env-90 | Haiti | Americas | 2012 | JSTI00000000 |
| env-94 | Haiti | Americas | 2012 | JSTK00000000 |
| Env4926 | Haiti | Americas | 2016 | Env4926 |
| Env5156 | Haiti | Americas | 2016 | Env5156 |
| Env6956 | Haiti | Americas | 2018 | Env6956 |
| EnvJ515 | Haiti | Americas | 2018 | EnvJ515 |
| ERR018111 | India | Asia | 2007 | ERR018111 |
| ERR018112 | India | Asia | 2006 | ERR018112 |
| ERR018113 | Bangladesh | Asia | 2001 | ERR018113 |
| ERR018114 | Bangladesh | Asia | 1999 | ERR018114 |
| ERR018115 | Bangladesh | Asia | 2001 | ERR018115 |
| ERR018116 | Bangladesh | Asia | 2001 | ERR018116 |
| ERR018119 | India | Asia | 1993 | ERR018119 |
| ERR018120 | Bangladesh | Asia | 1994 | ERR018120 |
| ERR018121 | Bangladesh | Asia | 2002 | ERR018121 |
| ERR018122 | Bangladesh | Asia | 1991 | ERR018122 |
| ERR018123 | India | Asia | 2004 | ERR018123 |
| ERR018124 | India | Asia | 1992 | ERR018124 |
| ERR018125 | India | Asia | 1991 | ERR018125 |
| ERR018126 | India | Asia | 1990 | ERR018126 |
| ERR018127 | India | Asia | 1989 | ERR018127 |
| ERR018128 | Bangladesh | Asia | 2006 | ERR018128 |
| ERR018130 | India | Asia | 2004 | ERR018130 |
| ERR018146 | Colombia | Americas | 1992 | ERR018146 |
| ERR018148 | Colombia | Americas | 1992 | ERR018148 |
| ERR018149 | Colombia | Americas | 1992 | ERR018149 |
| ERR018150 | Mozambique | Africa | 1991 | ERR018150 |
| ERR018151 | Mozambique | Africa | 1991 | ERR018151 |
| ERR018152 | Mozambique | Africa | 1991 | ERR018152 |
| ERR018153 | India | Asia | 1989 | ERR018153 |
| ERR018154 | India | Asia | 1989 | ERR018154 |
| ERR018156 | Colombia | Americas | 1992 | ERR018156 |
| ERR018158 | Argentina | Americas | 1992 | ERR018158 |
| ERR018159 | Mexico | Americas | 1991 | ERR018159 |
| ERR018160 | Bolivia | Americas | 1992 | ERR018160 |
| ERR018161 | Mexico | Americas | 1991 | ERR018161 |
| ERR018167 | Argentina | Americas | 1992 | ERR018167 |
| ERR018168 | Argentina | Americas | 1992 | ERR018168 |
| ERR018169 | Viet Nam | Asia | 1989 | ERR018169 |
| ERR018171 | Viet Nam | Asia | 1989 | ERR018171 |
| ERR018172 | Djibouti | Africa | 2007 | ERR018172 |

| | | | | |
|---|---|---|---|---|
| ERR018174 | Djibouti | Africa | 2007 | ERR018174 |
| ERR018175 | Djibouti | Africa | 2007 | ERR018175 |
| ERR018176 | Bangladesh | Asia | 1987 | ERR018176 |
| ERR018177 | Bangladesh | Asia | 1987 | ERR018177 |
| ERR018179 | Bangladesh | Asia | 1994 | ERR018179 |
| ERR018181 | Argentina | Americas | 1993 | ERR018181 |
| ERR018182 | Bangladesh | Asia | 2007 | ERR018182 |
| ERR018185 | India | Asia | 1984 | ERR018185 |
| ERR018186 | India | Asia | 1973 | ERR018186 |
| ERR018187 | India | Asia | 2009 | ERR018187 |
| ERR018188 | India | Asia | 1979 | ERR018188 |
| ERR018189 | India | Asia | 1980 | ERR018189 |
| ERR018192 | Bahrain | Middle East | 1978 | ERR018192 |
| ERR018193 | Malaysia | Asia | 1978 | ERR018193 |
| ERR018194 | Germany | Europe | 1975 | ERR018194 |
| ERR018195 | India | Asia | 1992 | ERR018195 |
| ERR019287 | Kenya | Africa | 2007 | ERR019287 |
| ERR019288 | Kenya | Africa | 2005 | ERR019288 |
| ERR019289 | Kenya | Africa | 2007 | ERR019289 |
| ERR019290 | Kenya | Africa | 2007 | ERR019290 |
| ERR019291 | Kenya | Africa | 2007 | ERR019291 |
| ERR019292 | Kenya | Africa | 2007 | ERR019292 |
| ERR019295 | Kenya | Africa | 2007 | ERR019295 |
| ERR019296 | Kenya | Africa | 2005 | ERR019296 |
| ERR019297 | Kenya | Africa | 2005 | ERR019297 |
| ERR019879 | Viet Nam | Asia | 1995 | ERR019879 |
| ERR019880 | Viet Nam | Asia | 2002 | ERR019880 |
| ERR019881 | India | Asia | 2004 | ERR019881 |
| ERR019882 | India | Asia | 2006 | ERR019882 |
| ERR019883 | Bangladesh | Asia | 1991 | ERR019883 |
| ERR019884 | Bangladesh | Asia | 2000 | ERR019884 |
| ERR019885 | Viet Nam | Asia | 2007 | ERR019885 |
| ERR025356 | Mozambique | Africa | 2005 | ERR025356 |
| ERR025357 | India | Asia | 2007 | ERR025357 |
| ERR025358 | India | Asia | 2007 | ERR025358 |
| ERR025359 | India | Asia | 2007 | ERR025359 |
| ERR025360 | India | Asia | 2007 | ERR025360 |
| ERR025361 | India | Asia | 2004 | ERR025361 |
| ERR025362 | Viet Nam | Asia | 2004 | ERR025362 |
| ERR025364 | Viet Nam | Asia | 2003 | ERR025364 |
| ERR025366 | India | Asia | 2007 | ERR025366 |
| ERR025367 | Mozambique | Africa | 2005 | ERR025367 |
| ERR025368 | India | Asia | 2007 | ERR025368 |

| | | | | |
|---|---|---|---|---|
| ERR025369 | India | Asia | 2006 | ERR025369 |
| ERR025370 | Tanzania | Africa | 2009 | ERR025370 |
| ERR025371 | India | Asia | 2007 | ERR025371 |
| ERR025372 | India | Asia | 2007 | ERR025372 |
| ERR025373 | Bangladesh | Asia | 2001 | ERR025373 |
| ERR025374 | India | Asia | 2005 | ERR025374 |
| ERR025375 | India | Asia | 2007 | ERR025375 |
| ERR025377 | Mozambique | Africa | 2005 | ERR025377 |
| ERR025378 | Peru | Americas | 1991 | ERR025378 |
| ERR025381 | Angola | Africa | 1989 | ERR025381 |
| ERR025382 | Indonesia | Asia | 1957 | ERR025382 |
| ERR025383 | Bangladesh | Asia | 1979 | ERR025383 |
| ERR025384 | India | Asia | 1977 | ERR025384 |
| ERR025385 | Bangladesh | Asia | 1971 | ERR025385 |
| ERR025386 | Bangladesh | Asia | 1979 | ERR025386 |
| ERR025388 | Peru | Americas | 1991 | ERR025388 |
| ERR025389 | Peru | Americas | 1991 | ERR025389 |
| ERR025390 | Peru | Americas | 1991 | ERR025390 |
| ERR025393 | Bangladesh | Asia | 2006 | ERR025393 |
| ERR025395 | Bangladesh | Asia | 2007 | ERR025395 |
| ERR025396 | Bangladesh | Asia | 1991 | ERR025396 |
| ERR028066 | Kenya | Africa | 2009 | ERR028066 |
| ERR028068 | Kenya | Africa | 2009 | ERR028068 |
| ERR028074 | Kenya | Africa | 2009 | ERR028074 |
| ERR028075 | Kenya | Africa | 2009 | ERR028075 |
| ERR028076 | Kenya | Africa | 2009 | ERR028076 |
| ERR037738 | Kenya | Africa | 2010 | ERR037738 |
| ERR037739 | Kenya | Africa | 2010 | ERR037739 |
| ERR037741 | Kenya | Africa | 2010 | ERR037741 |
| ERR042729 | Palestine | Middle East | 1994 | ERR042729 |
| ERR042733 | Gaza | Middle East | 1994 | ERR042733 |
| ERR042738 | Palestine | Middle East | 1994 | ERR042738 |
| ERR042745 | Israel | Middle East | 1970 | ERR042745 |
| ERR042748 | Israel | Middle East | 1970 | ERR042748 |
| ERR042749 | Israel | Middle East | 1970 | ERR042749 |
| ERR042754 | Mexico | Americas | 1992 | ERR042754 |
| ERR044779 | Mexico | Americas | 1991 | ERR044779 |
| ERR044780 | Mexico | Americas | 1991 | ERR044780 |
| ERR044781 | Mexico | Americas | 1991 | ERR044781 |
| ERR044782 | Mexico | Americas | 1992 | ERR044782 |
| ERR044783 | Mexico | Americas | 1992 | ERR044783 |
| ERR044784 | Mexico | Americas | 1993 | ERR044784 |
| ERR044785 | Mexico | Americas | 1993 | ERR044785 |

| | | | | |
|---|---|---|---|---|
| ERR044786 | Mexico | Americas | 1993 | ERR044786 |
| ERR044787 | Mexico | Americas | 1994 | ERR044787 |
| ERR044788 | Mexico | Americas | 1995 | ERR044788 |
| ERR044791 | Mexico | Americas | 1997 | ERR044791 |
| ERR044794 | Zambia | Africa | 1996 | ERR044794 |
| ERR044795 | Zambia | Africa | 2003 | ERR044795 |
| ERR044797 | Mozambique | Africa | 2004 | ERR044797 |
| ERR044798 | Zimbabwe | Africa | 2009 | ERR044798 |
| ERR044799 | Zimbabwe | Africa | 2009 | ERR044799 |
| ERR044800 | Zimbabwe | Africa | 2009 | ERR044800 |
| ERR051745 | Pakistan | Asia | 2010 | ERR051745 |
| ERR051746 | Pakistan | Asia | 2010 | ERR051746 |
| ERR051748 | Pakistan | Asia | 2010 | ERR051748 |
| ERR051749 | Pakistan | Asia | 2010 | ERR051749 |
| ERR051751 | Pakistan | Asia | 2010 | ERR051751 |
| ERR051752 | Pakistan | Asia | 2010 | ERR051752 |
| ERR051755 | Pakistan | Asia | 2010 | ERR051755 |
| ERR051756 | Pakistan | Asia | 2010 | ERR051756 |
| ERR051758 | Pakistan | Asia | 2010 | ERR051758 |
| ERR051759 | Pakistan | Asia | 2010 | ERR051759 |
| ERR051760 | Pakistan | Asia | 2010 | ERR051760 |
| ERR051761 | Pakistan | Asia | 2010 | ERR051761 |
| ERR051762 | Pakistan | Asia | 2010 | ERR051762 |
| ERR051763 | Pakistan | Asia | 2010 | ERR051763 |
| ERR051764 | Pakistan | Asia | 2010 | ERR051764 |
| ERR051765 | Pakistan | Asia | 2010 | ERR051765 |
| ERR051767 | Pakistan | Asia | 2010 | ERR051767 |
| ERR051768 | Pakistan | Asia | 2010 | ERR051768 |
| ERR051769 | Pakistan | Asia | 2010 | ERR051769 |
| ERR051770 | Pakistan | Asia | 2010 | ERR051770 |
| ERR051771 | Pakistan | Asia | 2010 | ERR051771 |
| ERR051772 | Pakistan | Asia | 2010 | ERR051772 |
| ERR051773 | Pakistan | Asia | 2010 | ERR051773 |
| ERR051775 | Pakistan | Asia | 2010 | ERR051775 |
| ERR051776 | Pakistan | Asia | 2010 | ERR051776 |
| ERR051777 | Pakistan | Asia | 2010 | ERR051777 |
| ERR051779 | Pakistan | Asia | 2010 | ERR051779 |
| ERR051780 | Pakistan | Asia | 2010 | ERR051780 |
| ERR051781 | Pakistan | Asia | 2010 | ERR051781 |
| ERR051782 | Pakistan | Asia | 2010 | ERR051782 |
| ERR051783 | Pakistan | Asia | 2010 | ERR051783 |
| ERR051784 | Pakistan | Asia | 2010 | ERR051784 |
| ERR051785 | Pakistan | Asia | 2010 | ERR051785 |

| | | | | |
|---|---|---|---|---|
| ERR051786 | Pakistan | Asia | 2010 | ERR051786 |
| ERR051787 | Pakistan | Asia | 2010 | ERR051787 |
| ERR051788 | Pakistan | Asia | 2010 | ERR051788 |
| ERR051789 | Pakistan | Asia | 2010 | ERR051789 |
| ERR051790 | Pakistan | Asia | 2010 | ERR051790 |
| ERR1024518 | Ghana | Africa | 2011 | ERR1024518 |
| ERR1024521 | Ghana | Africa | 2014 | ERR1024521 |
| ERR1024524 | Ghana | Africa | 2014 | ERR1024524 |
| ERR1024526 | Ghana | Africa | 2014 | ERR1024526 |
| ERR108516 | Mexico | Americas | 1998 | ERR108516 |
| ERR108517 | Mexico | Americas | 1999 | ERR108517 |
| ERR108518 | Mexico | Americas | 1999 | ERR108518 |
| ERR108522 | Mexico | Americas | 2000 | ERR108522 |
| ERR108535 | Mexico | Americas | 2004 | ERR108535 |
| ERR108536 | Mexico | Americas | 2005 | ERR108536 |
| ERR108537 | Mexico | Americas | 2006 | ERR108537 |
| ERR114415 | Kenya | Africa | 2009 | ERR114415 |
| ERR114416 | Kenya | Africa | 2009 | ERR114416 |
| ERR114418 | Kenya | Africa | 2009 | ERR114418 |
| ERR117473 | Kenya | Africa | 2010 | ERR117473 |
| ERR117480 | Kenya | Africa | 2009 | ERR117480 |
| ERR117482 | Kenya | Africa | 2009 | ERR117482 |
| ERR117484 | Kenya | Africa | 2010 | ERR117484 |
| ERR117485 | Kenya | Africa | 2010 | ERR117485 |
| ERR117490 | Kenya | Africa | 2010 | ERR117490 |
| ERR117555 | Kenya | Africa | 2009 | ERR117555 |
| ERR117556 | Kenya | Africa | 2009 | ERR117556 |
| ERR117569 | Kenya | Africa | 2010 | ERR117569 |
| ERR117571 | Kenya | Africa | 2010 | ERR117571 |
| ERR117572 | Kenya | Africa | 2009 | ERR117572 |
| ERR117573 | Kenya | Africa | 2010 | ERR117573 |
| ERR117577 | Kenya | Africa | 2010 | ERR117577 |
| ERR117578 | Kenya | Africa | 2010 | ERR117578 |
| ERR117579 | Kenya | Africa | 2007 | ERR117579 |
| ERR117581 | Kenya | Africa | 2010 | ERR117581 |
| ERR117583 | Kenya | Africa | 2010 | ERR117583 |
| ERR117586 | Kenya | Africa | 2010 | ERR117586 |
| ERR117587 | Kenya | Africa | 2010 | ERR117587 |
| ERR117588 | Kenya | Africa | 2010 | ERR117588 |
| ERR117590 | Kenya | Africa | 2010 | ERR117590 |
| ERR117591 | Kenya | Africa | 2010 | ERR117591 |
| ERR117592 | Kenya | Africa | 2009 | ERR117592 |
| ERR117593 | Kenya | Africa | 2007 | ERR117593 |

| | | | | |
|---|---|---|---|---|
| ERR117594 | Kenya | Africa | 2009 | ERR117594 |
| ERR117595 | Kenya | Africa | 2010 | ERR117595 |
| ERR117597 | Kenya | Africa | 2010 | ERR117597 |
| ERR117599 | Kenya | Africa | 2010 | ERR117599 |
| ERR117600 | Kenya | Africa | 2009 | ERR117600 |
| ERR117603 | Kenya | Africa | 2009 | ERR117603 |
| ERR117604 | Kenya | Africa | 2009 | ERR117604 |
| ERR117605 | Kenya | Africa | 2009 | ERR117605 |
| ERR117606 | Kenya | Africa | 2008 | ERR117606 |
| ERR117607 | Kenya | Africa | 2009 | ERR117607 |
| ERR117609 | Kenya | Africa | 2009 | ERR117609 |
| ERR117610 | Kenya | Africa | 2009 | ERR117610 |
| ERR117612 | Kenya | Africa | 2009 | ERR117612 |
| ERR117613 | Kenya | Africa | 2009 | ERR117613 |
| ERR163233 | Mexico | Americas | 1991 | ERR163233 |
| ERR163234 | Mexico | Americas | 1991 | ERR163234 |
| ERR163235 | Mexico | Americas | 1991 | ERR163235 |
| ERR163236 | Mexico | Americas | 1991 | ERR163236 |
| ERR163237 | Mexico | Americas | 1991 | ERR163237 |
| ERR163238 | Mexico | Americas | 1992 | ERR163238 |
| ERR163239 | Mexico | Americas | 1992 | ERR163239 |
| ERR163240 | Mexico | Americas | 1993 | ERR163240 |
| ERR163241 | Mexico | Americas | 1994 | ERR163241 |
| ERR163242 | Mexico | Americas | 1994 | ERR163242 |
| ERR163243 | Mexico | Americas | 1995 | ERR163243 |
| ERR163244 | Mexico | Americas | 1995 | ERR163244 |
| ERR163245 | Mexico | Americas | 2004 | ERR163245 |
| ERR163246 | Mexico | Americas | 2004 | ERR163246 |
| ERR163247 | Mexico | Americas | 2004 | ERR163247 |
| ERR163248 | Mexico | Americas | 2004 | ERR163248 |
| ERR163249 | Mexico | Americas | 2007 | ERR163249 |
| ERR163250 | Mexico | Americas | 2007 | ERR163250 |
| ERR163253 | Mexico | Americas | 2008 | ERR163253 |
| ERR163254 | Mexico | Americas | 2008 | ERR163254 |
| ERR163255 | Mexico | Americas | 2008 | ERR163255 |
| ERR163256 | Mexico | Americas | 2008 | ERR163256 |
| ERR163259 | Mexico | Americas | 2010 | ERR163259 |
| ERR163260 | Mexico | Americas | 2010 | ERR163260 |
| ERR163263 | Mexico | Americas | 2010 | ERR163263 |
| ERR163268 | Mexico | Americas | 2010 | ERR163268 |
| ERR163271 | Mexico | Americas | 2010 | ERR163271 |
| ERR164757 | Zambia | Africa | 1996 | ERR164757 |
| ERR164758 | Zambia | Africa | 1996 | ERR164758 |

| | | | | |
|---|---|---|---|---|
| ERR164759 | Zambia | Africa | 1997 | ERR164759 |
| ERR164760 | Zambia | Africa | 1997 | ERR164760 |
| ERR164763 | Zambia | Africa | 1997 | ERR164763 |
| ERR164765 | Zambia | Africa | 2003 | ERR164765 |
| ERR164766 | Zambia | Africa | 2003 | ERR164766 |
| ERR164768 | Zambia | Africa | 2004 | ERR164768 |
| ERR164773 | Zambia | Africa | 2004 | ERR164773 |
| ERR164776 | Zambia | Africa | 2004 | ERR164776 |
| ERR187761 | South Africa | Africa | 2001 | ERR1877613 |
| ERR187761 | Rwanda | Africa | 1998 | ERR1877614 |
| ERR187761 | Rwanda | Africa | 1998 | ERR1877615 |
| ERR187761 | Mozambique | Africa | 1992 | ERR1877616 |
| ERR187761 | Burundi | Africa | 1992 | ERR1877617 |
| ERR187761 | Burundi | Africa | 1992 | ERR1877618 |
| ERR187761 | Uganda | Africa | 1992 | ERR1877619 |
| ERR187763 | Argentina | Americas | 1992 | ERR1877636 |
| ERR187763 | Niger | Africa | 1999 | ERR1877637 |
| ERR187763 | Uganda | Africa | 2000 | ERR1877638 |
| ERR187763 | Uganda | Africa | 2000 | ERR1877639 |
| ERR187764 | Comoros | Africa | 2000 | ERR1877640 |
| ERR187764 | Rwanda | Africa | 2000 | ERR1877641 |
| ERR187764 | Rwanda | Africa | 2000 | ERR1877642 |
| ERR187764 | Cameroon | Africa | 2000 | ERR1877643 |
| ERR187764 | Comoros | Africa | 2001 | ERR1877644 |
| ERR187764 | Cameroon | Africa | 2000 | ERR1877645 |
| ERR187764 | Cote d'Ivoire | Africa | 2001 | ERR1877646 |
| ERR187764 | Cote d'Ivoire | Africa | 2001 | ERR1877647 |
| ERR187764 | Comoros | Africa | 2001 | ERR1877648 |
| ERR187764 | Djibouti | Africa | 2000 | ERR1877649 |
| ERR187793 | Djibouti | Africa | 2000 | ERR1877938 |
| ERR187793 | Madagascar | Africa | 2000 | ERR1877939 |
| ERR187794 | Madagascar | Africa | 2001 | ERR1877940 |
| ERR187794 | Burundi | Africa | 2001 | ERR1877941 |
| ERR187794 | Burundi | Africa | 2001 | ERR1877942 |
| ERR187794 | Indonesia | Asia | 2001 | ERR1877943 |
| ERR187794 | Togo | Africa | 2001 | ERR1877944 |
| ERR187794 | Chad | Africa | 2001 | ERR1877945 |
| ERR187794 | Chad | Africa | 2001 | ERR1877946 |
| ERR187794 | Chad | Africa | 2001 | ERR1877947 |
| ERR187794 | Burkina Fasc | Africa | 2001 | ERR1877948 |
| ERR187794 | Burkina Fasc | Africa | 2001 | ERR1877949 |
| ERR187795 | Cameroon | Africa | 2001 | ERR1877950 |
| ERR187795 | Democratic F | Africa | 2001 | ERR1877951 |

| | | | | |
|---|---|---|---|---|
| ERR1877952 | Democratic R | Africa | 2001 | ERR1877952 |
| ERR1877953 | Comoros | Africa | 2002 | ERR1877953 |
| ERR1877954 | Liberia | Africa | 2002 | ERR1877954 |
| ERR1877955 | Democratic R | Africa | 2002 | ERR1877955 |
| ERR1877956 | Benin | Africa | 2002 | ERR1877956 |
| ERR1877957 | Benin | Africa | 2002 | ERR1877957 |
| ERR1878088 | Liberia | Africa | 2002 | ERR1878088 |
| ERR1878089 | Cote d'Ivoire | Africa | 2002 | ERR1878089 |
| ERR1878090 | Democratic R | Africa | 2003 | ERR1878090 |
| ERR1878091 | Benin | Africa | 2003 | ERR1878091 |
| ERR1878092 | Benin | Africa | 2003 | ERR1878092 |
| ERR1878093 | Democratic R | Africa | 2003 | ERR1878093 |
| ERR1878094 | Somalia | Africa | 2003 | ERR1878094 |
| ERR1878095 | Somalia | Africa | 2003 | ERR1878095 |
| ERR1878096 | Iraq | Middle East | 2003 | ERR1878096 |
| ERR1878097 | Democratic R | Africa | 2003 | ERR1878097 |
| ERR1878098 | Liberia | Africa | 2003 | ERR1878098 |
| ERR1878099 | Liberia | Africa | 2003 | ERR1878099 |
| ERR1878100 | Cote d'Ivoire | Africa | 2003 | ERR1878100 |
| ERR1878101 | Democratic R | Africa | 2002 | ERR1878101 |
| ERR1878102 | Cote d'Ivoire | Africa | 2003 | ERR1878102 |
| ERR1878103 | Comoros | Africa | 2003 | ERR1878103 |
| ERR1878104 | Comoros | Africa | 2003 | ERR1878104 |
| ERR1878105 | Cameroon | Africa | 2004 | ERR1878105 |
| ERR1878106 | Cameroon | Africa | 2001 | ERR1878106 |
| ERR1878107 | Cameroon | Africa | 2002 | ERR1878107 |
| ERR1878108 | Cameroon | Africa | 2004 | ERR1878108 |
| ERR1878109 | Benin | Africa | 2004 | ERR1878109 |
| ERR1878110 | Benin | Africa | 2004 | ERR1878110 |
| ERR1878111 | Sierra Leone | Africa | 2004 | ERR1878111 |
| ERR1878112 | Guinea | Africa | 2004 | ERR1878112 |
| ERR1878113 | Niger | Africa | 2004 | ERR1878113 |
| ERR1878114 | Niger | Africa | 2004 | ERR1878114 |
| ERR1878115 | Sierra Leone | Africa | 2004 | ERR1878115 |
| ERR1878116 | Democratic R | Africa | 2004 | ERR1878116 |
| ERR1878117 | Senegal | Africa | 2004 | ERR1878117 |
| ERR1878127 | Senegal | Africa | 2004 | ERR1878127 |
| ERR1878128 | Cameroon | Africa | 2005 | ERR1878128 |
| ERR1878129 | Cameroon | Africa | 2005 | ERR1878129 |
| ERR1878130 | Equatorial Gu | Africa | 2005 | ERR1878130 |
| ERR1878131 | Equatorial Gu | Africa | 2005 | ERR1878131 |
| ERR1878132 | Democratic R | Africa | 2005 | ERR1878132 |
| ERR1878133 | Benin | Africa | 2005 | ERR1878133 |

| | | | | |
|---|---|---|---|---|
| ERR1878134 | Benin | Africa | 2005 | ERR1878134 |
| ERR1878135 | Pakistan | Asia | 2005 | ERR1878135 |
| ERR1878136 | Democratic R | Africa | 2005 | ERR1878136 |
| ERR1878146 | Nigeria | Africa | 2005 | ERR1878146 |
| ERR1878147 | Nigeria | Africa | 2005 | ERR1878147 |
| ERR1878148 | Democratic R | Africa | 2006 | ERR1878148 |
| ERR1878149 | Mali | Africa | 2006 | ERR1878149 |
| ERR1878150 | Mali | Africa | 2005 | ERR1878150 |
| ERR1878151 | India | Asia | 2006 | ERR1878151 |
| ERR1878152 | Cameroon | Africa | 2006 | ERR1878152 |
| ERR1878153 | Kenya | Africa | 2006 | ERR1878153 |
| ERR1878154 | Kenya | Africa | 2006 | ERR1878154 |
| ERR1878155 | Democratic R | Africa | 2006 | ERR1878155 |
| ERR1878544 | Niger | Africa | 2006 | ERR1878544 |
| ERR1878545 | Niger | Africa | 2006 | ERR1878545 |
| ERR1878546 | Nigeria | Africa | 2006 | ERR1878546 |
| ERR1878547 | Nigeria | Africa | 2006 | ERR1878547 |
| ERR1878548 | India | Asia | 2006 | ERR1878548 |
| ERR1878549 | Democratic R | Africa | 2007 | ERR1878549 |
| ERR1878550 | Democratic R | Africa | 2007 | ERR1878550 |
| ERR1878551 | Djibouti | Africa | 2007 | ERR1878551 |
| ERR1878552 | Democratic R | Africa | 2007 | ERR1878552 |
| ERR1878553 | South Sudan | Africa | 2007 | ERR1878553 |
| ERR1878554 | South Sudan | Africa | 2007 | ERR1878554 |
| ERR1878555 | Algeria | Africa | 1994 | ERR1878555 |
| ERR1878556 | Mauritania | Africa | 2005 | ERR1878556 |
| ERR1878557 | Mauritania | Africa | 2006 | ERR1878557 |
| ERR1878558 | Mauritania | Africa | 2006 | ERR1878558 |
| ERR1878559 | Mauritania | Africa | 2005 | ERR1878559 |
| ERR1878560 | Algeria | Africa | 1994 | ERR1878560 |
| ERR1878561 | Benin | Africa | 2007 | ERR1878561 |
| ERR1878562 | Burkina Faso | Africa | 2005 | ERR1878562 |
| ERR1878563 | Burkina Faso | Africa | 2005 | ERR1878563 |
| ERR1878564 | Democratic R | Africa | 2007 | ERR1878564 |
| ERR1878565 | Niger | Africa | 2008 | ERR1878565 |
| ERR1878566 | Niger | Africa | 2008 | ERR1878566 |
| ERR1878567 | Democratic R | Africa | 2008 | ERR1878567 |
| ERR1878568 | Democratic R | Africa | 2008 | ERR1878568 |
| ERR1878569 | South Sudan | Africa | 2008 | ERR1878569 |
| ERR1878570 | India | Asia | 2008 | ERR1878570 |
| ERR1878571 | Côte d'Ivoire | Africa | 2006 | ERR1878571 |
| ERR1878572 | Cote d'Ivoire | Africa | 2006 | ERR1878572 |
| ERR1878573 | Cameroon | Africa | 2008 | ERR1878573 |

| | | | | |
|---|---|---|---|---|
| ERR1878574 | Cameroon | Africa | 2008 | ERR1878574 |
| ERR1878575 | Pakistan | Asia | 2009 | ERR1878575 |
| ERR1878576 | Pakistan | Asia | 2009 | ERR1878576 |
| ERR1878577 | Pakistan | Asia | 2009 | ERR1878577 |
| ERR1878578 | Nigeria | Africa | 2009 | ERR1878578 |
| ERR1878579 | Nigeria | Africa | 2009 | ERR1878579 |
| ERR1878580 | Cameroon | Africa | 2009 | ERR1878580 |
| ERR1878581 | Zambia | Africa | 2010 | ERR1878581 |
| ERR1878582 | Zambia | Africa | 2010 | ERR1878582 |
| ERR1878583 | Nigeria | Africa | 2010 | ERR1878583 |
| ERR1878586 | Benin | Africa | 2010 | ERR1878586 |
| ERR1878587 | Benin | Africa | 2010 | ERR1878587 |
| ERR1878588 | Cameroon | Africa | 2010 | ERR1878588 |
| ERR1878589 | Nigeria | Africa | 2010 | ERR1878589 |
| ERR1878590 | Chad | Africa | 2010 | ERR1878590 |
| ERR1878591 | Chad | Africa | 2010 | ERR1878591 |
| ERR1878593 | Nigeria | Africa | 2011 | ERR1878593 |
| ERR1878594 | Nigeria | Africa | 2011 | ERR1878594 |
| ERR1878595 | Cameroon | Africa | 2010 | ERR1878595 |
| ERR1878596 | Cameroon | Africa | 2011 | ERR1878596 |
| ERR1878597 | Cameroon | Africa | 2011 | ERR1878597 |
| ERR1878598 | Cameroon | Africa | 2010 | ERR1878598 |
| ERR1878599 | Chad | Africa | 2011 | ERR1878599 |
| ERR1878600 | Chad | Africa | 2011 | ERR1878600 |
| ERR1878601 | Nigeria | Africa | 2011 | ERR1878601 |
| ERR1878602 | Central Afric | Africa | 2011 | ERR1878602 |
| ERR1878603 | Central Afric | Africa | 2011 | ERR1878603 |
| ERR1878604 | Guinea Bissa | Africa | 2012 | ERR1878604 |
| ERR1878605 | Guinea Bissa | Africa | 2012 | ERR1878605 |
| ERR1878607 | Democratic R | Africa | 2014 | ERR1878607 |
| ERR1878608 | Democratic R | Africa | 2014 | ERR1878608 |
| ERR1878609 | Nigeria | Africa | 2014 | ERR1878609 |
| ERR1878610 | Nigeria | Africa | 2014 | ERR1878610 |
| ERR1878611 | Niger | Africa | 2010 | ERR1878611 |
| ERR1878612 | Niger | Africa | 2010 | ERR1878612 |
| ERR1878613 | Ukraine | Europe | 1970 | ERR1878613 |
| ERR1878614 | Ukraine | Europe | 1974 | ERR1878614 |
| ERR1878615 | Russia | Asia | 1990 | ERR1878615 |
| ERR1878616 | Russia | Asia | 1990 | ERR1878616 |
| ERR1879384 | Cote d'Ivoire | Africa | 1988 | ERR1879384 |
| ERR1879385 | Kenya | Africa | 1998 | ERR1879385 |
| ERR1879386 | Tanzania | Africa | 1998 | ERR1879386 |
| ERR1879387 | Ethiopia | Africa | 1998 | ERR1879387 |

| | | | | |
|---|---|---|---|---|
| ERR1879388 | Romania | Europe | 1981 | ERR1879388 |
| ERR1879389 | Lebanon | Middle East | 1993 | ERR1879389 |
| ERR1879390 | Turkey | Asia | 1976 | ERR1879390 |
| ERR1879391 | Ghana | Africa | 1982 | ERR1879391 |
| ERR1879392 | Ethiopia | Africa | 1985 | ERR1879392 |
| ERR1879393 | Romania | Europe | 1994 | ERR1879393 |
| ERR1879429 | Pakistan | Asia | 1992 | ERR1879429 |
| ERR1879430 | Romania | Europe | 1991 | ERR1879430 |
| ERR1879431 | Romania | Europe | 1987 | ERR1879431 |
| ERR1879432 | Lebanon | Middle East | 1993 | ERR1879432 |
| ERR1879433 | Romania | Europe | 1990 | ERR1879433 |
| ERR1879434 | Senegal | Africa | 1994 | ERR1879434 |
| ERR1879435 | Cote d'Ivoire | Africa | 1988 | ERR1879435 |
| ERR1879436 | Tunisia | Africa | 1973 | ERR1879436 |
| ERR1879437 | Tunisia | Africa | 1981 | ERR1879437 |
| ERR1879438 | Italy | Europe | 1994 | ERR1879438 |
| ERR1879535 | Cote d'Ivoire | Africa | 1988 | ERR1879535 |
| ERR1879536 | Somalia | Africa | 1985 | ERR1879536 |
| ERR1879537 | Somalia | Africa | 1985 | ERR1879537 |
| ERR1879538 | Cambodia | Asia | 1993 | ERR1879538 |
| ERR1879539 | Kenya | Africa | 1998 | ERR1879539 |
| ERR1879540 | Kenya | Africa | 1998 | ERR1879540 |
| ERR1879541 | Italy | Europe | 1973 | ERR1879541 |
| ERR1879542 | Cote d'Ivoire | Africa | 1988 | ERR1879542 |
| ERR1879543 | Kenya | Africa | 1998 | ERR1879543 |
| ERR1879544 | Kenya | Africa | 1998 | ERR1879544 |
| ERR1879545 | Ethiopia | Africa | 1998 | ERR1879545 |
| ERR1879546 | Guinea | Africa | 1990 | ERR1879546 |
| ERR1879547 | Mozambique | Africa | 1990 | ERR1879547 |
| ERR1879548 | Cambodia | Asia | 1991 | ERR1879548 |
| ERR1879549 | Turkey | Asia | 1991 | ERR1879549 |
| ERR1879550 | Iraq | Middle East | 1991 | ERR1879550 |
| ERR1879551 | Viet Nam | Asia | 1983 | ERR1879551 |
| ERR1879552 | Sao Tome | Africa | 1989 | ERR1879552 |
| ERR1879553 | India | Asia | 1992 | ERR1879553 |
| ERR1879554 | French Guiar | Americas | 1993 | ERR1879554 |
| ERR1879559 | Myanmar | Asia | 1971 | ERR1879559 |
| ERR1879560 | Slovakia | Europe | 1970 | ERR1879560 |
| ERR1879561 | Israel | Middle East | 1970 | ERR1879561 |
| ERR1879562 | India | Asia | 1962 | ERR1879562 |
| ERR1879563 | Lebanon | Middle East | 1970 | ERR1879563 |
| ERR1879564 | Philippines | Asia | 1961 | ERR1879564 |
| ERR1879565 | China | Asia | 1961 | ERR1879565 |

| | | | | |
|---|---|---|---|---|
| ERR187956( | India | Asia | 1964 | ERR1879566 |
| ERR1879567 | Cambodia | Asia | 1963 | ERR1879567 |
| ERR1879568 | Iran | Middle East | 1965 | ERR1879568 |
| ERR1879569 | France | Europe | 1970 | ERR1879569 |
| ERR187957( | Thailand | Asia | 1993 | ERR1879570 |
| ERR1879571 | Sri Lanka | Asia | 1981 | ERR1879571 |
| ERR1879572 | Djibouti | Africa | 1993 | ERR1879572 |
| ERR1879573 | Philippines | Asia | 1963 | ERR1879573 |
| ERR1879574 | Romania | Europe | 1991 | ERR1879574 |
| ERR1879575 | Somalia | Africa | 1994 | ERR1879575 |
| ERR187957( | Nepal | Asia | 1994 | ERR1879576 |
| ERR1879577 | Albania | Europe | 1994 | ERR1879577 |
| ERR1879578 | French Guiar | Americas | 1994 | ERR1879578 |
| ERR1879579 | Ecuador | Americas | 1995 | ERR1879579 |
| ERR187958( | Malaysia | Asia | 1973 | ERR1879580 |
| ERR1879581 | Viet Nam | Asia | 1966 | ERR1879581 |
| ERR1879582 | Viet Nam | Asia | 1968 | ERR1879582 |
| ERR1879583 | Viet Nam | Asia | 1969 | ERR1879583 |
| ERR1879584 | Philippines | Asia | 1973 | ERR1879584 |
| ERR1879585 | Italy | Europe | 1974 | ERR1879585 |
| ERR187958( | Senegal | Africa | 1974 | ERR1879586 |
| ERR1879587 | Myanmar | Asia | 1972 | ERR1879587 |
| ERR1879588 | Iraq | Middle East | 1995 | ERR1879588 |
| ERR1879593 | Iraq | Middle East | 1995 | ERR1879593 |
| ERR1879594 | Iraq | Middle East | 1995 | ERR1879594 |
| ERR1879595 | Sudan | Africa | 1996 | ERR1879595 |
| ERR1879597 | India | Asia | 1996 | ERR1879597 |
| ERR1879598 | Mauritania | Africa | 1996 | ERR1879598 |
| ERR1879599 | Mauritania | Africa | 1996 | ERR1879599 |
| ERR187960( | Sudan | Africa | 1996 | ERR1879600 |
| ERR1879601 | Iraq | Middle East | 1966 | ERR1879601 |
| ERR1879602 | Iraq | Middle East | 1966 | ERR1879602 |
| ERR1879603 | Iran | Middle East | 1965 | ERR1879603 |
| ERR1879604 | Iran | Middle East | 1965 | ERR1879604 |
| ERR1879605 | Iran | Middle East | 1965 | ERR1879605 |
| ERR187960( | Iran | Middle East | 1965 | ERR1879606 |
| ERR1879607 | Iran | Middle East | 1965 | ERR1879607 |
| ERR1879624 | Israel | Middle East | 1970 | ERR1879624 |
| ERR1879625 | Israel | Middle East | 1970 | ERR1879625 |
| ERR187962( | Israel | Middle East | 1970 | ERR1879626 |
| ERR1879627 | Jordan | Middle East | 1970 | ERR1879627 |
| ERR1879628 | Lebanon | Middle East | 1970 | ERR1879628 |
| ERR1879629 | New Guinea | Oceania | 1962 | ERR1879629 |

| | | | | |
|---|---|---|---|---|
| ERR187963( | Pakistan | Asia | 1964 | ERR1879630 |
| ERR1879631 | Slovakia | Europe | 1970 | ERR1879631 |
| ERR1879632 | Slovakia | Europe | 1970 | ERR1879632 |
| ERR187963: | Thailand | Asia | 1963 | ERR1879633 |
| ERR187963⁄ | Viet Nam | Asia | 1964 | ERR1879634 |
| ERR187963( | Burundi | Africa | 1997 | ERR1879636 |
| ERR187963 | India | Asia | 1997 | ERR1879637 |
| ERR187963{ | Chad | Africa | 1998 | ERR1879638 |
| ERR187964⁄ | Cambodia | Asia | 1999 | ERR1879644 |
| ERR187964: | Laos | Asia | 1999 | ERR1879645 |
| ERR187964( | Burkina Fasc | Africa | 1999 | ERR1879646 |
| ERR187964 | Democratic F | Africa | 1999 | ERR1879647 |
| ERR187964{ | Afghanistan | Asia | 1999 | ERR1879648 |
| ERR187964{ | Afghanistan | Asia | 1999 | ERR1879649 |
| ERR187965( | Cameroon | Africa | 1999 | ERR1879650 |
| ERR1879651 | Cameroon | Africa | 1993 | ERR1879651 |
| ERR1879652 | Djibouti | Africa | 1993 | ERR1879652 |
| ERR187965: | South Africa | Africa | 1996 | ERR1879653 |
| ERR187965⁄ | South Africa | Africa | 1996 | ERR1879654 |
| ERR187965: | Tanzania | Africa | 1997 | ERR1879655 |
| ERR187965( | Nigeria | Africa | 1997 | ERR1879656 |
| ERR187965 | Nigeria | Africa | 1997 | ERR1879657 |
| ERR187965{ | Malawi | Africa | 1997 | ERR1879658 |
| ERR1880761 | Malawi | Africa | 1997 | ERR1880761 |
| ERR1880762 | Guinea Bissa | Africa | 1997 | ERR1880762 |
| ERR188076: | Mozambique | Africa | 1997 | ERR1880763 |
| ERR188076⁄ | Zambia | Africa | 1998 | ERR1880764 |
| ERR188076: | Zambia | Africa | 1998 | ERR1880765 |
| ERR188076( | Tanzania | Africa | 1998 | ERR1880766 |
| ERR188076 | Tanzania | Africa | 1998 | ERR1880767 |
| ERR188076{ | South Africa | Africa | 2002 | ERR1880768 |
| ERR188076{ | South Africa | Africa | 2002 | ERR1880769 |
| ERR188077( | Mali | Africa | 2003 | ERR1880770 |
| ERR1880771 | Mali | Africa | 2003 | ERR1880771 |
| ERR1880772 | Guinea Bissa | Africa | 2008 | ERR1880772 |
| ERR188077: | Guinea Bissa | Africa | 2008 | ERR1880773 |
| ERR188077⁄ | South Africa | Africa | 2001 | ERR1880774 |
| ERR188077: | South Africa | Africa | 2001 | ERR1880775 |
| ERR188077( | South Africa | Africa | 2001 | ERR1880776 |
| ERR188077 | South Africa | Africa | 2001 | ERR1880777 |
| ERR188077{ | South Africa | Africa | 2001 | ERR1880778 |
| ERR188077{ | South Africa | Africa | 2001 | ERR1880779 |
| ERR188078( | South Africa | Africa | 2001 | ERR1880780 |

| | | | | |
|---|---|---|---|---|
| ERR1880781 | South Africa | Africa | 2001 | ERR1880781 |
| ERR1880782 | South Africa | Africa | 2001 | ERR1880782 |
| ERR1880783 | South Africa | Africa | 2001 | ERR1880783 |
| ERR1880784 | South Africa | Africa | 2001 | ERR1880784 |
| ERR1880785 | South Africa | Africa | 2001 | ERR1880785 |
| ERR1880786 | South Africa | Africa | 1980 | ERR1880786 |
| ERR1880787 | South Africa | Africa | 2001 | ERR1880787 |
| ERR1880788 | South Africa | Africa | 2001 | ERR1880788 |
| ERR1880789 | South Africa | Africa | 1998 | ERR1880789 |
| ERR1880790 | South Africa | Africa | 2001 | ERR1880790 |
| ERR1880791 | South Africa | Africa | 1980 | ERR1880791 |
| ERR1880792 | South Africa | Africa | 1980 | ERR1880792 |
| ERR1880793 | Angola | Africa | 1972 | ERR1880793 |
| ERR1880794 | Turkey | Asia | 1994 | ERR1880794 |
| ERR1880795 | Turkey | Asia | 1994 | ERR1880795 |
| ERR1880796 | Spain | Europe | 1971 | ERR1880796 |
| ERR1880797 | Turkey | Asia | 1980 | ERR1880797 |
| ERR1880798 | Sao Tome | Africa | 1989 | ERR1880798 |
| ERR1880799 | India | Asia | 1996 | ERR1880799 |
| ERR1880800 | India | Asia | 1996 | ERR1880800 |
| ERR1880801 | India | Asia | 1997 | ERR1880801 |
| ERR1880802 | India | Asia | 1997 | ERR1880802 |
| ERR1880803 | India | Asia | 1995 | ERR1880803 |
| ERR1880804 | India | Asia | 1993 | ERR1880804 |
| ERR1880805 | India | Asia | 1993 | ERR1880805 |
| ERR1880806 | India | Asia | 1994 | ERR1880806 |
| ERR1880807 | India | Asia | 1994 | ERR1880807 |
| ERR1880808 | India | Asia | 1995 | ERR1880808 |
| ERR1880809 | India | Asia | 2000 | ERR1880809 |
| ERR1880810 | India | Asia | 2000 | ERR1880810 |
| ERR1880811 | India | Asia | 1998 | ERR1880811 |
| ERR1880812 | India | Asia | 1998 | ERR1880812 |
| ERR1880813 | India | Asia | 1999 | ERR1880813 |
| ERR1880814 | India | Asia | 2001 | ERR1880814 |
| ERR1880815 | Togo | Africa | 1975 | ERR1880815 |
| ERR1880816 | India | Asia | 1980 | ERR1880816 |
| ERR1880817 | India | Asia | 2002 | ERR1880817 |
| ERR1880818 | India | Asia | 2002 | ERR1880818 |
| ERR1880819 | India | Asia | 2003 | ERR1880819 |
| ERR1880820 | India | Asia | 2003 | ERR1880820 |
| ERR1880821 | India | Asia | 2007 | ERR1880821 |
| ERR1880822 | India | Asia | 2007 | ERR1880822 |
| ERR1880823 | India | Asia | 2008 | ERR1880823 |

| | | | | |
|---|---|---|---|---|
| ERR1880824 | India | Asia | 2008 | ERR1880824 |
| ERR1880825 | India | Asia | 2009 | ERR1880825 |
| ERR1880826 | India | Asia | 2009 | ERR1880826 |
| ERR1880827 | India | Asia | 2010 | ERR1880827 |
| ERR1880828 | India | Asia | 2010 | ERR1880828 |
| ERR1880829 | India | Asia | 2004 | ERR1880829 |
| ERR1880830 | India | Asia | 2004 | ERR1880830 |
| ERR1880831 | India | Asia | 2005 | ERR1880831 |
| ERR1880832 | India | Asia | 2005 | ERR1880832 |
| ERR1880833 | India | Asia | 2006 | ERR1880833 |
| ERR1880834 | India | Asia | 2006 | ERR1880834 |
| ERR1880835 | India | Asia | 1999 | ERR1880835 |
| ERR1880836 | India | Asia | 2001 | ERR1880836 |
| ERR1880837 | India | Asia | 1990 | ERR1880837 |
| ERR1880838 | India | Asia | 1989 | ERR1880838 |
| ERR1880839 | India | Asia | 1990 | ERR1880839 |
| ERR1880840 | India | Asia | 1991 | ERR1880840 |
| ERR1880841 | India | Asia | 1991 | ERR1880841 |
| ERR1880842 | India | Asia | 1989 | ERR1880842 |
| ERR1880843 | India | Asia | 1992 | ERR1880843 |
| ERR1880844 | India | Asia | 1992 | ERR1880844 |
| ERR2008340 | Mexico | Americas | 1991 | ERR2008340 |
| ERR2008341 | Mexico | Americas | 1991 | ERR2008341 |
| ERR2008342 | Mexico | Americas | 1991 | ERR2008342 |
| ERR2008343 | Mexico | Americas | 1991 | ERR2008343 |
| ERR2008344 | Mexico | Americas | 1991 | ERR2008344 |
| ERR2008345 | Mexico | Americas | 1991 | ERR2008345 |
| ERR2008346 | Mexico | Americas | 1991 | ERR2008346 |
| ERR2008347 | Mexico | Americas | 1991 | ERR2008347 |
| ERR2008348 | Mexico | Americas | 1992 | ERR2008348 |
| ERR2008349 | Mexico | Americas | 1992 | ERR2008349 |
| ERR2008350 | Mexico | Americas | 1992 | ERR2008350 |
| ERR2008352 | Mexico | Americas | 1999 | ERR2008352 |
| ERR2008353 | Mexico | Americas | 1999 | ERR2008353 |
| ERR2008358 | Mexico | Americas | 1992 | ERR2008358 |
| ERR2008359 | Mexico | Americas | 1992 | ERR2008359 |
| ERR2008360 | Mexico | Americas | 1992 | ERR2008360 |
| ERR2008361 | Mexico | Americas | 1992 | ERR2008361 |
| ERR2008362 | Mexico | Americas | 1992 | ERR2008362 |
| ERR2008363 | Mexico | Americas | 1992 | ERR2008363 |
| ERR2008364 | Mexico | Americas | 1992 | ERR2008364 |
| ERR2008365 | Mexico | Americas | 1993 | ERR2008365 |
| ERR2008366 | Mexico | Americas | 1993 | ERR2008366 |

| | | | | |
|---|---|---|---|---|
| ERR2008386 | Mexico | Americas | 1993 | ERR2008386 |
| ERR2008387 | Mexico | Americas | 1993 | ERR2008387 |
| ERR2008388 | Mexico | Americas | 1993 | ERR2008388 |
| ERR2008389 | Mexico | Americas | 1993 | ERR2008389 |
| ERR2008390 | Mexico | Americas | 1993 | ERR2008390 |
| ERR2008391 | Mexico | Americas | 1993 | ERR2008391 |
| ERR2008392 | Mexico | Americas | 1994 | ERR2008392 |
| ERR2008393 | Mexico | Americas | 1994 | ERR2008393 |
| ERR2008394 | Mexico | Americas | 1994 | ERR2008394 |
| ERR2008395 | Mexico | Americas | 1994 | ERR2008395 |
| ERR2008396 | Mexico | Americas | 1994 | ERR2008396 |
| ERR2008397 | Mexico | Americas | 1994 | ERR2008397 |
| ERR2008398 | Mexico | Americas | 1995 | ERR2008398 |
| ERR2008399 | Mexico | Americas | 1995 | ERR2008399 |
| ERR2008400 | Mexico | Americas | 1995 | ERR2008400 |
| ERR2008401 | Mexico | Americas | 1995 | ERR2008401 |
| ERR2008402 | Mexico | Americas | 1995 | ERR2008402 |
| ERR2008403 | Mexico | Americas | 1995 | ERR2008403 |
| ERR2008404 | Mexico | Americas | 1995 | ERR2008404 |
| ERR2008405 | Mexico | Americas | 1995 | ERR2008405 |
| ERR2008406 | Mexico | Americas | 1995 | ERR2008406 |
| ERR2008407 | Mexico | Americas | 1995 | ERR2008407 |
| ERR2008408 | Mexico | Americas | 1995 | ERR2008408 |
| ERR2008409 | Mexico | Americas | 1995 | ERR2008409 |
| ERR2008410 | Mexico | Americas | 1996 | ERR2008410 |
| ERR2008411 | Mexico | Americas | 1996 | ERR2008411 |
| ERR2008412 | Mexico | Americas | 2000 | ERR2008412 |
| ERR2008413 | Mexico | Americas | 1998 | ERR2008413 |
| ERR2008414 | Mexico | Americas | 2000 | ERR2008414 |
| ERR2008418 | Mexico | Americas | 1996 | ERR2008418 |
| ERR2008419 | Mexico | Americas | 1997 | ERR2008419 |
| ERR2008420 | Mexico | Americas | 1997 | ERR2008420 |
| ERR2008421 | Mexico | Americas | 1996 | ERR2008421 |
| ERR2008422 | Mexico | Americas | 1997 | ERR2008422 |
| ERR2008423 | Mexico | Americas | 1997 | ERR2008423 |
| ERR2008424 | Mexico | Americas | 1997 | ERR2008424 |
| ERR2008425 | Mexico | Americas | 1997 | ERR2008425 |
| ERR2008426 | Mexico | Americas | 1997 | ERR2008426 |
| ERR2008430 | Mexico | Americas | 1998 | ERR2008430 |
| ERR2008433 | Mexico | Americas | 2000 | ERR2008433 |
| ERR2008434 | Mexico | Americas | 2000 | ERR2008434 |
| ERR2008746 | Mexico | Americas | 1997 | ERR2008746 |
| ERR2008748 | Mexico | Americas | 1998 | ERR2008748 |

| | | | | |
|---|---|---|---|---|
| ERR200874[illegible] | Mexico | Americas | 1998 | ERR2008749 |
| ERR200875[illegible] | Mexico | Americas | 1998 | ERR2008750 |
| ERR2008751 | Mexico | Americas | 1997 | ERR2008751 |
| ERR200875[illegible] | Mexico | Americas | 1998 | ERR2008752 |
| ERR200875[illegible] | Bolivia | Americas | 1991 | ERR2008756 |
| ERR200875[illegible] | Brazil | Americas | 1991 | ERR2008757 |
| ERR200875[illegible] | Brazil | Americas | 1991 | ERR2008758 |
| ERR200875[illegible] | Brazil | Americas | 1991 | ERR2008759 |
| ERR200876[illegible] | Colombia | Americas | 1991 | ERR2008760 |
| ERR2008761 | Colombia | Americas | 1992 | ERR2008761 |
| ERR200876[illegible] | Peru | Americas | 1991 | ERR2008762 |
| ERR214473[illegible] | Mozambique | Africa | 2010 | ERS1938068 |
| ERR214475[illegible] | Mozambique | Africa | 2010 | ERS1938072 |
| ERR214475[illegible] | Mozambique | Africa | 2002 | ERS1938058 |
| ERR214476[illegible] | Mozambique | Africa | 2003 | ERS1938085 |
| ERR214477[illegible] | Mozambique | Africa | 2003 | ERS1938079 |
| ERR2144781 | Mozambique | Africa | 2002 | ERS1938051 |
| ERR214478[illegible] | Mozambique | Africa | 2003 | ERS1938060 |
| ERR214479[illegible] | Mozambique | Africa | 2002 | ERS1938059 |
| ERR226558[illegible] | Kenya | Africa | 2015 | ERR2265589 |
| ERR226559[illegible] | Bangladesh | Asia | 2011 | ERR2265590 |
| ERR2265591 | South Sudan | Africa | 2014 | ERR2265591 |
| ERR226564[illegible] | South Sudan | Africa | 2014 | ERR2265645 |
| ERR226564[illegible] | South Sudan | Africa | 2014 | ERR2265646 |
| ERR226564[illegible] | South Sudan | Africa | 2014 | ERR2265647 |
| ERR226564[illegible] | South Sudan | Africa | 2014 | ERR2265648 |
| ERR226564[illegible] | India | Asia | 2014 | ERR2265649 |
| ERR226565[illegible] | Democratic R | Africa | 2015 | ERR2265650 |
| ERR2265651 | Democratic R | Africa | 2015 | ERR2265651 |
| ERR226565[illegible] | India | Asia | 2015 | ERR2265652 |
| ERR226565[illegible] | South Sudan | Africa | 2015 | ERR2265653 |
| ERR226565[illegible] | South Sudan | Africa | 2015 | ERR2265654 |
| ERR226565[illegible] | Iraq | Middle East | 2015 | ERR2265655 |
| ERR226565[illegible] | Iraq | Middle East | 2015 | ERR2265656 |
| ERR226565[illegible] | Iraq | Middle East | 2015 | ERR2265657 |
| ERR226565[illegible] | Iraq | Middle East | 2015 | ERR2265658 |
| ERR226565[illegible] | Iraq | Middle East | 2015 | ERR2265659 |
| ERR226566[illegible] | Iraq | Middle East | 2015 | ERR2265660 |
| ERR2265661 | Democratic R | Africa | 2015 | ERR2265661 |
| ERR226566[illegible] | Iraq | Middle East | 2007 | ERR2265662 |
| ERR226566[illegible] | Iraq | Middle East | 2007 | ERR2265663 |
| ERR226566[illegible] | Iraq | Middle East | 2007 | ERR2265664 |
| ERR226566[illegible] | Iraq | Middle East | 2007 | ERR2265665 |

| | | | | |
|---|---|---|---|---|
| ERR226566( | Iraq | Middle East | 2007 | ERR2265666 |
| ERR226566, | South Sudan | Africa | 2016 | ERR2265667 |
| ERR226566{ | South Sudan | Africa | 2016 | ERR2265668 |
| ERR226566( | South Sudan | Africa | 2017 | ERR2265669 |
| ERR226567( | South Sudan | Africa | 2017 | ERR2265670 |
| ERR226567] | South Sudan | Africa | 2017 | ERR2265671 |
| ERR226567: | South Sudan | Africa | 2017 | ERR2265672 |
| ERR226567: | South Sudan | Africa | 2017 | ERR2265673 |
| ERR226567< | Yemen | Middle East | 2016 | ERR2265674 |
| ERR226567: | Yemen | Middle East | 2016 | ERR2265675 |
| ERR226567( | Yemen | Middle East | 2016 | ERR2265676 |
| ERR226567, | Yemen | Middle East | 2016 | ERR2265677 |
| ERR226567{ | Yemen | Middle East | 2016 | ERR2265678 |
| ERR226961: | Yemen | Middle East | 2016 | ERR2269613 |
| ERR226961< | Yemen | Middle East | 2016 | ERR2269614 |
| ERR226961: | Yemen | Middle East | 2016 | ERR2269615 |
| ERR226961( | Yemen | Middle East | 2017 | ERR2269616 |
| ERR226961, | Yemen | Middle East | 2017 | ERR2269617 |
| ERR226961{ | Yemen | Middle East | 2017 | ERR2269618 |
| ERR226961( | Yemen | Middle East | 2017 | ERR2269619 |
| ERR226962( | Yemen | Middle East | 2017 | ERR2269620 |
| ERR226962] | Yemen | Middle East | 2017 | ERR2269621 |
| ERR226962: | Yemen | Middle East | 2017 | ERR2269622 |
| ERR226964( | Yemen | Middle East | 2017 | ERR2269640 |
| ERR226964] | Yemen | Middle East | 2017 | ERR2269641 |
| ERR226964: | Yemen | Middle East | 2017 | ERR2269642 |
| ERR226964: | Yemen | Middle East | 2017 | ERR2269643 |
| ERR226964< | Yemen | Middle East | 2017 | ERR2269644 |
| ERR226964: | Yemen | Middle East | 2017 | ERR2269645 |
| ERR226964( | Yemen | Middle East | 2017 | ERR2269646 |
| ERR226964, | Yemen | Middle East | 2017 | ERR2269647 |
| ERR226964{ | Yemen | Middle East | 2017 | ERR2269648 |
| ERR226964( | Yemen | Middle East | 2017 | ERR2269649 |
| ERR226965( | Yemen | Middle East | 2017 | ERR2269650 |
| ERR226970( | Yemen | Middle East | 2017 | ERR2269709 |
| ERR226971( | Yemen | Middle East | 2017 | ERR2269710 |
| ERR226971] | Yemen | Middle East | 2017 | ERR2269711 |
| ERR226971: | Yemen | Middle East | 2017 | ERR2269712 |
| ERR226971: | Yemen | Middle East | 2017 | ERR2269713 |
| ERR226971< | Yemen | Middle East | 2017 | ERR2269714 |
| ERR226971: | Yemen | Middle East | 2017 | ERR2269715 |
| ERR226971( | Yemen | Middle East | 2017 | ERR2269716 |
| ERR226971, | Yemen | Middle East | 2017 | ERR2269717 |

| | | | | |
|---|---|---|---|---|
| ERR2269718 | Yemen | Middle East | 2017 | ERR2269718 |
| ERR2269808 | India | Asia | 2010 | ERR2269808 |
| ERR2269809 | Yemen | Middle East | 2017 | ERR2269809 |
| ERR2269810 | Yemen | Middle East | 2017 | ERR2269810 |
| ERR2269811 | Yemen | Middle East | 2017 | ERR2269811 |
| ERR2269832 | Yemen | Middle East | 2017 | ERR2269832 |
| ERR2269833 | Yemen | Middle East | 2017 | ERR2269833 |
| ERR2269834 | Yemen | Middle East | 2017 | ERR2269834 |
| ERR2269835 | Iran | Middle East | 2012 | ERR2269835 |
| ERR2269836 | Iran | Middle East | 2013 | ERR2269836 |
| ERR2269837 | Iran | Middle East | 2013 | ERR2269837 |
| ERR2269838 | Iran | Middle East | 2015 | ERR2269838 |
| ERR2269921 | India | Asia | 2010 | ERR2269921 |
| ERR2269922 | India | Asia | 2010 | ERR2269922 |
| ERR2269923 | India | Asia | 2011 | ERR2269923 |
| ERR2269924 | India | Asia | 2011 | ERR2269924 |
| ERR2269925 | India | Asia | 2011 | ERR2269925 |
| ERR2269926 | India | Asia | 2011 | ERR2269926 |
| ERR2269927 | India | Asia | 2012 | ERR2269927 |
| ERR2269928 | India | Asia | 2012 | ERR2269928 |
| ERR2269929 | India | Asia | 2012 | ERR2269929 |
| ERR2269930 | India | Asia | 2013 | ERR2269930 |
| ERR2269944 | India | Asia | 2013 | ERR2269944 |
| ERR2269945 | India | Asia | 2013 | ERR2269945 |
| ERR2269946 | India | Asia | 2013 | ERR2269946 |
| ERR2269947 | India | Asia | 2014 | ERR2269947 |
| ERR2269948 | India | Asia | 2014 | ERR2269948 |
| ERR2269949 | India | Asia | 2014 | ERR2269949 |
| ERR2269950 | India | Asia | 2015 | ERR2269950 |
| ERR2269951 | India | Asia | 2015 | ERR2269951 |
| ERR2269952 | India | Asia | 2015 | ERR2269952 |
| ERR2269953 | India | Asia | 2015 | ERR2269953 |
| ERR2270655 | India | Asia | 2016 | ERR2270655 |
| ERR2270656 | India | Asia | 2016 | ERR2270656 |
| ERR2270657 | India | Asia | 2016 | ERR2270657 |
| ERR2270658 | India | Asia | 2016 | ERR2270658 |
| ERR2270659 | India | Asia | 2017 | ERR2270659 |
| ERR2270660 | India | Asia | 2017 | ERR2270660 |
| ERR2270661 | India | Asia | 2017 | ERR2270661 |
| ERR2270662 | India | Asia | 2017 | ERR2270662 |
| ERR2275156 | Kenya | Africa | 2015 | ERS1572783 |
| ERR2275157 | Kenya | Africa | 2015 | ERS1572784 |
| ERR2275158 | Kenya | Africa | 2015 | ERS1572785 |

| | | | | |
|---|---|---|---|---|
| ERR2275159 | Kenya | Africa | 2015 | ERS1572793 |
| ERR2275160 | Kenya | Africa | 2012 | ERS1572798 |
| ERR2275161 | Kenya | Africa | 2012 | ERS1572803 |
| ERR2275162 | Kenya | Africa | 2012 | ERS1572809 |
| ERR2275163 | Kenya | Africa | 2010 | ERS1572813 |
| ERR2275164 | Kenya | Africa | 2016 | ERS1572815 |
| ERR2442217 | Tanzania | Africa | 2015 | ERS2318682 |
| ERR2442219 | Tanzania | Africa | 2015 | ERS2318685 |
| ERR2442221 | Tanzania | Africa | 2015 | ERS2318718 |
| ERR2442225 | Tanzania | Africa | 2015 | ERS2318693 |
| ERR2442226 | Tanzania | Africa | 2015 | ERS2318708 |
| ERR2442229 | Tanzania | Africa | 2015 | ERS2318689 |
| ERR2442231 | Tanzania | Africa | 2015 | ERS2318680 |
| ERR2442232 | Tanzania | Africa | 2015 | ERS2318712 |
| ERR2442234 | Tanzania | Africa | 2015 | ERS2318697 |
| ERR2442236 | Tanzania | Africa | 2015 | ERS2318700 |
| ERR2442238 | Tanzania | Africa | 2012 | ERS2318703 |
| ERR2442245 | Tanzania | Africa | 2011 | ERS2318704 |
| ERR2442246 | Tanzania | Africa | 2015 | ERS2318681 |
| ERR2442252 | Tanzania | Africa | 2012 | ERS2318705 |
| ERR3342504 | Zimbabwe | Africa | 2018 | ERR3342504 |
| ERR3342505 | Zimbabwe | Africa | 2018 | ERR3342505 |
| ERR3342506 | Zimbabwe | Africa | 2018 | ERR3342506 |
| ERR3342507 | Zimbabwe | Africa | 2018 | ERR3342507 |
| ERR3342508 | Zimbabwe | Africa | 2018 | ERR3342508 |
| ERR3342509 | Zimbabwe | Africa | 2018 | ERR3342509 |
| ERR3342510 | Zimbabwe | Africa | 2018 | ERR3342510 |
| ERR3342511 | Zimbabwe | Africa | 2018 | ERR3342511 |
| ERR3342512 | Zimbabwe | Africa | 2018 | ERR3342512 |
| ERR3342513 | Zimbabwe | Africa | 2018 | ERR3342513 |
| ERR3342514 | Zimbabwe | Africa | 2018 | ERR3342514 |
| ERR3342515 | Zimbabwe | Africa | 2018 | ERR3342515 |
| ERR3342516 | Zimbabwe | Africa | 2018 | ERR3342516 |
| ERR351174 | India | Asia | 2009 | ERR351174 |
| ERR351175 | India | Asia | 2009 | ERR351175 |
| ERR351177 | India | Asia | 2009 | ERS353751 |
| ERR351179 | India | Asia | 2009 | ERR351179 |
| ERR351182 | India | Asia | 2009 | ERS353756 |
| ERR351185 | India | Asia | 2009 | ERR351185 |
| ERR351186 | India | Asia | 2009 | ERS353760 |
| ERR351188 | India | Asia | 2009 | ERR351188 |
| ERR351191 | India | Asia | 2009 | ERR351191 |
| ERR351192 | India | Asia | 2009 | ERR351192 |

| | | | | |
|---|---|---|---|---|
| ERR351193 | India | Asia | 2009 | ERR351193 |
| ERR351198 | India | Asia | 2009 | ERS353772 |
| ERR351199 | India | Asia | 2009 | ERR351199 |
| ERR351202 | India | Asia | 2009 | ERR351202 |
| ERR351203 | India | Asia | 2009 | ERR351203 |
| ERR351205 | India | Asia | 2009 | ERR351205 |
| ERR351206 | India | Asia | 2009 | ERR351206 |
| ERR351207 | India | Asia | 2009 | ERR351207 |
| ERR351208 | India | Asia | 2009 | ERR351208 |
| ERR351210 | India | Asia | 2009 | ERR351210 |
| ERR3771803 | Argentina | Americas | 1996 | ERS2493998 |
| ERR3771804 | Argentina | Americas | 1996 | ERS2493999 |
| ERR3771805 | Argentina | Americas | 1996 | ERS2494000 |
| ERR3771807 | Argentina | Americas | 1996 | ERS2493974 |
| ERR3771810 | Argentina | Americas | 1993 | ERS2493977 |
| ERR3771814 | Argentina | Americas | 1993 | ERS2493981 |
| ERR3771815 | Argentina | Americas | 1993 | ERS2493982 |
| ERR3771816 | Argentina | Americas | 1993 | ERS2493983 |
| ERR3771817 | Argentina | Americas | 1993 | ERS2493985 |
| ERR3771818 | Argentina | Americas | 1993 | ERS2493986 |
| ERR3771819 | Argentina | Americas | 1993 | ERS2493987 |
| ERR3771820 | Argentina | Americas | 1993 | ERS2493988 |
| ERR3771821 | Argentina | Americas | 1993 | ERS2493989 |
| ERR3771823 | Argentina | Americas | 1992 | ERS2494001 |
| ERR3771824 | Argentina | Americas | 1993 | ERS2494002 |
| ERR3771825 | Argentina | Americas | 1992 | ERS2494003 |
| ERR3771826 | Argentina | Americas | 1993 | ERS2494004 |
| ERR3771827 | Argentina | Americas | 1993 | ERS2494005 |
| ERR3771828 | Argentina | Americas | 1993 | ERS2494006 |
| ERR3771829 | Argentina | Americas | 1992 | ERS2494008 |
| ERR3771830 | Argentina | Americas | 1992 | ERS2494009 |
| ERR3771831 | Argentina | Americas | 1993 | ERS2494010 |
| ERR3771832 | Argentina | Americas | 1993 | ERS2494011 |
| ERR3771833 | Argentina | Americas | 1993 | ERS2494012 |
| ERR3771834 | Argentina | Americas | 1993 | ERS2494013 |
| ERR3771835 | Argentina | Americas | 1993 | ERS2494014 |
| ERR3771836 | Argentina | Americas | 1993 | ERS2494015 |
| ERR3771837 | Argentina | Americas | 1993 | ERS2494016 |
| ERR3771838 | Argentina | Americas | 1993 | ERS2494017 |
| ERR3771839 | Argentina | Americas | 1993 | ERS2494018 |
| ERR3771840 | Argentina | Americas | 1993 | ERS2494019 |
| ERR3771841 | Argentina | Americas | 1993 | ERS2494020 |
| ERR3771843 | Argentina | Americas | 1993 | ERS2494022 |

| | | | | |
|---|---|---|---|---|
| ERR3771844 | Argentina | Americas | 1993 | ERS2494023 |
| ERR3771845 | Argentina | Americas | 1993 | ERS2494024 |
| ERR3771846 | Argentina | Americas | 1993 | ERS2494025 |
| ERR3771847 | Argentina | Americas | 1993 | ERS2494026 |
| ERR3771848 | Argentina | Americas | 1993 | ERS2494027 |
| ERR3771849 | Argentina | Americas | 1993 | ERS2494028 |
| ERR3771850 | Argentina | Americas | 1993 | ERS2494029 |
| ERR3771851 | Argentina | Americas | 1993 | ERS2494030 |
| ERR3771852 | Argentina | Americas | 1993 | ERS2494031 |
| ERR3771854 | Argentina | Americas | 1993 | ERS2494033 |
| ERR3771855 | Argentina | Americas | 1993 | ERS2494034 |
| ERR3771856 | Argentina | Americas | 1993 | ERS2494035 |
| ERR3771857 | Argentina | Americas | 1993 | ERS2494036 |
| ERR3771858 | Argentina | Americas | 1993 | ERS2494037 |
| ERR3771859 | Argentina | Americas | 1993 | ERS2494038 |
| ERR3771860 | Argentina | Americas | 1993 | ERS2493884 |
| ERR3771861 | Argentina | Americas | 1993 | ERS2493885 |
| ERR3771862 | Argentina | Americas | 1993 | ERS2493886 |
| ERR3771863 | Argentina | Americas | 1993 | ERS2493887 |
| ERR3771864 | Argentina | Americas | 1993 | ERS2493888 |
| ERR3771865 | Argentina | Americas | 1993 | ERS2493889 |
| ERR3771866 | Argentina | Americas | 1993 | ERS2493890 |
| ERR3771867 | Argentina | Americas | 1993 | ERS2493891 |
| ERR3771868 | Argentina | Americas | 1993 | ERS2493892 |
| ERR3771869 | Argentina | Americas | 1993 | ERS2493893 |
| ERR3771870 | Argentina | Americas | 1993 | ERS2493894 |
| ERR3771871 | Argentina | Americas | 1993 | ERS2493895 |
| ERR3771872 | Argentina | Americas | 1993 | ERS2493896 |
| ERR3771873 | Argentina | Americas | 1993 | ERS2493897 |
| ERR3771874 | Argentina | Americas | 1993 | ERS2493898 |
| ERR3771875 | Argentina | Americas | 1993 | ERS2493899 |
| ERR3771876 | Argentina | Americas | 1993 | ERS2493900 |
| ERR3771877 | Argentina | Americas | 1993 | ERS2493901 |
| ERR3771878 | Argentina | Americas | 1993 | ERS2493902 |
| ERR3771879 | Argentina | Americas | 1993 | ERS2493903 |
| ERR3771880 | Argentina | Americas | 1993 | ERS2493904 |
| ERR3771881 | Argentina | Americas | 1993 | ERS2493905 |
| ERR3771882 | Argentina | Americas | 1993 | ERS2493906 |
| ERR3771883 | Argentina | Americas | 1997 | ERS2493907 |
| ERR3771884 | Argentina | Americas | 1997 | ERS2493908 |
| ERR3771885 | Argentina | Americas | 1996 | ERS2493909 |
| ERR3771886 | Argentina | Americas | 1996 | ERS2493910 |
| ERR3771887 | Argentina | Americas | 1997 | ERS2493911 |

| | | | | |
|---|---|---|---|---|
| ERR3771888 | Argentina | Americas | 1997 | ERS2493912 |
| ERR3771889 | Argentina | Americas | 1997 | ERS2493913 |
| ERR3771890 | Argentina | Americas | 1996 | ERS2493914 |
| ERR3771891 | Argentina | Americas | 1997 | ERS2493915 |
| ERR3771893 | Argentina | Americas | 1997 | ERS2493917 |
| ERR3771894 | Argentina | Americas | 1993 | ERS2493918 |
| ERR3771895 | Argentina | Americas | 1993 | ERS2493919 |
| ERR3771896 | Argentina | Americas | 1993 | ERS2493921 |
| ERR3771897 | Argentina | Americas | 1993 | ERS2493922 |
| ERR3771898 | Argentina | Americas | 1997 | ERS2493923 |
| ERR3771899 | Argentina | Americas | 1993 | ERS2493924 |
| ERR3771900 | Argentina | Americas | 1993 | ERS2493925 |
| ERR3771901 | Argentina | Americas | 1993 | ERS2493926 |
| ERR3771902 | Argentina | Americas | 1993 | ERS2493927 |
| ERR3771903 | Argentina | Americas | 1993 | ERS2493928 |
| ERR3771904 | Argentina | Americas | 1993 | ERS2493929 |
| ERR3771905 | Argentina | Americas | 1993 | ERS2493930 |
| ERR3771906 | Argentina | Americas | 1993 | ERS2493931 |
| ERR3771907 | Argentina | Americas | 1993 | ERS2493932 |
| ERR3771909 | Argentina | Americas | 1993 | ERS2493934 |
| ERR3771910 | Argentina | Americas | 1993 | ERS2493935 |
| ERR3771911 | Argentina | Americas | 1993 | ERS2493936 |
| ERR3771912 | Argentina | Americas | 1993 | ERS2493937 |
| ERR3771913 | Argentina | Americas | 1993 | ERS2493938 |
| ERR3771914 | Argentina | Americas | 1993 | ERS2493939 |
| ERR3771915 | Argentina | Americas | 1993 | ERS2493940 |
| ERR3771916 | Argentina | Americas | 1993 | ERS2493941 |
| ERR3771917 | Argentina | Americas | 1993 | ERS2493942 |
| ERR3771918 | Argentina | Americas | 1993 | ERS2493943 |
| ERR3771919 | Argentina | Americas | 1993 | ERS2493944 |
| ERR3771920 | Argentina | Americas | 1993 | ERS2493945 |
| ERR3771921 | Argentina | Americas | 1993 | ERS2493946 |
| ERR3771922 | Argentina | Americas | 1993 | ERS2493947 |
| ERR3771923 | Argentina | Americas | 1993 | ERS2493948 |
| ERR3771924 | Argentina | Americas | 1993 | ERS2493949 |
| ERR3771925 | Argentina | Americas | 1993 | ERS2493950 |
| ERR3771926 | Argentina | Americas | 1993 | ERS2493951 |
| ERR3771927 | Argentina | Americas | 1993 | ERS2493952 |
| ERR3771929 | Argentina | Americas | 1993 | ERS2493954 |
| ERR3771932 | Argentina | Americas | 1993 | ERS2493957 |
| ERR3771933 | Argentina | Americas | 1993 | ERS2493958 |
| ERR3771934 | Argentina | Americas | 1993 | ERS2493959 |
| ERR3771935 | Argentina | Americas | 1993 | ERS2493960 |

| | | | | |
|---|---|---|---|---|
| ERR3771936 | Argentina | Americas | 1993 | ERS2493961 |
| ERR3771937 | Argentina | Americas | 1993 | ERS2493962 |
| ERR3771938 | Argentina | Americas | 1993 | ERS2493963 |
| ERR3771939 | Argentina | Americas | 1993 | ERS2493964 |
| ERR3771940 | Argentina | Americas | 1993 | ERS2493965 |
| ERR3771941 | Argentina | Americas | 1993 | ERS2493966 |
| ERR3771942 | Argentina | Americas | 1993 | ERS2493967 |
| ERR3771943 | Argentina | Americas | 1993 | ERS2493968 |
| ERR3771944 | Argentina | Americas | 1993 | ERS2493970 |
| ERR3771945 | Argentina | Americas | 1993 | ERS2493971 |
| ERR3771946 | Argentina | Americas | 1993 | ERS2493972 |
| ERR3771947 | Argentina | Americas | 1993 | ERS2493991 |
| ERR3771948 | Argentina | Americas | 1993 | ERS2493992 |
| ERR3771949 | Argentina | Americas | 1993 | ERS2493993 |
| ERR3771950 | Argentina | Americas | 1993 | ERS2493994 |
| ERR3771951 | Argentina | Americas | 1993 | ERS2493995 |
| ERR3771952 | Argentina | Americas | 1997 | ERS2493996 |
| ERR3771953 | Argentina | Americas | 1997 | ERS2493997 |
| ERR3771954 | Argentina | Americas | 1993 | ERS2493788 |
| ERR3771955 | Argentina | Americas | 1993 | ERS2493789 |
| ERR3771956 | Argentina | Americas | 1993 | ERS2493791 |
| ERR3771957 | Argentina | Americas | 1992 | ERS2493794 |
| ERR3771958 | Argentina | Americas | 1992 | ERS2493795 |
| ERR3771959 | Argentina | Americas | 1992 | ERS2493796 |
| ERR3771960 | Argentina | Americas | 1992 | ERS2493797 |
| ERR3771961 | Argentina | Americas | 1993 | ERS2493798 |
| ERR3771962 | Argentina | Americas | 1993 | ERS2493799 |
| ERR3771963 | Argentina | Americas | 1992 | ERS2493800 |
| ERR3771964 | Argentina | Americas | 1993 | ERS2493801 |
| ERR3771965 | Argentina | Americas | 1992 | ERS2493802 |
| ERR3771966 | Argentina | Americas | 1992 | ERS2493803 |
| ERR3771967 | Argentina | Americas | 1992 | ERS2493804 |
| ERR3771968 | Argentina | Americas | 1992 | ERS2493805 |
| ERR3771969 | Argentina | Americas | 1992 | ERS2493806 |
| ERR3771970 | Argentina | Americas | 1992 | ERS2493807 |
| ERR3771971 | Argentina | Americas | 1992 | ERS2493808 |
| ERR3771972 | Argentina | Americas | 1992 | ERS2493809 |
| ERR3771973 | Argentina | Americas | 1992 | ERS2493810 |
| ERR3771974 | Argentina | Americas | 1992 | ERS2493811 |
| ERR3771975 | Argentina | Americas | 1993 | ERS2493812 |
| ERR3771976 | Argentina | Americas | 1992 | ERS2493813 |
| ERR3771977 | Argentina | Americas | 1993 | ERS2493814 |
| ERR3771978 | Argentina | Americas | 1992 | ERS2493815 |

| | | | | |
|---|---|---|---|---|
| ERR3771979 | Argentina | Americas | 1992 | ERS2493816 |
| ERR3771980 | Argentina | Americas | 1997 | ERS2493817 |
| ERR3771981 | Argentina | Americas | 1993 | ERS2493818 |
| ERR3771982 | Argentina | Americas | 1993 | ERS2493819 |
| ERR3771983 | Argentina | Americas | 1992 | ERS2493820 |
| ERR3771984 | Argentina | Americas | 1992 | ERS2493821 |
| ERR3771985 | Argentina | Americas | 1992 | ERS2493822 |
| ERR3771986 | Argentina | Americas | 1993 | ERS2493823 |
| ERR3771987 | Argentina | Americas | 1993 | ERS2493824 |
| ERR3771988 | Argentina | Americas | 1993 | ERS2493825 |
| ERR3771989 | Argentina | Americas | 1992 | ERS2493826 |
| ERR3771990 | Argentina | Americas | 1993 | ERS2493827 |
| ERR3771991 | Argentina | Americas | 1992 | ERS2493828 |
| ERR3771992 | Argentina | Americas | 1993 | ERS2493829 |
| ERR3771993 | Argentina | Americas | 1993 | ERS2493830 |
| ERR3771994 | Argentina | Americas | 1993 | ERS2493831 |
| ERR3771995 | Argentina | Americas | 1993 | ERS2493832 |
| ERR3771996 | Argentina | Americas | 1993 | ERS2493833 |
| ERR3771997 | Argentina | Americas | 1993 | ERS2493834 |
| ERR3771998 | Argentina | Americas | 1993 | ERS2493835 |
| ERR3771999 | Argentina | Americas | 1993 | ERS2493836 |
| ERR3772000 | Argentina | Americas | 1993 | ERS2493837 |
| ERR3772001 | Argentina | Americas | 1993 | ERS2493840 |
| ERR3772002 | Argentina | Americas | 1993 | ERS2493841 |
| ERR3772003 | Argentina | Americas | 1993 | ERS2493842 |
| ERR3772005 | Argentina | Americas | 1993 | ERS2493844 |
| ERR3772006 | Argentina | Americas | 1993 | ERS2493845 |
| ERR3772007 | Argentina | Americas | 1993 | ERS2493846 |
| ERR3772008 | Argentina | Americas | 1993 | ERS2493847 |
| ERR3772009 | Argentina | Americas | 1993 | ERS2493848 |
| ERR3772010 | Argentina | Americas | 1993 | ERS2493849 |
| ERR3772011 | Argentina | Americas | 1993 | ERS2493850 |
| ERR3772012 | Argentina | Americas | 1993 | ERS2493851 |
| ERR3772013 | Argentina | Americas | 1993 | ERS2493852 |
| ERR3772014 | Argentina | Americas | 1993 | ERS2493853 |
| ERR3772015 | Argentina | Americas | 1993 | ERS2493854 |
| ERR3772016 | Argentina | Americas | 1993 | ERS2493855 |
| ERR3772017 | Argentina | Americas | 1993 | ERS2493856 |
| ERR3772018 | Argentina | Americas | 1993 | ERS2493857 |
| ERR3772019 | Argentina | Americas | 1993 | ERS2493858 |
| ERR3772020 | Argentina | Americas | 1993 | ERS2493859 |
| ERR3772021 | Argentina | Americas | 1993 | ERS2493860 |
| ERR3772022 | Argentina | Americas | 1993 | ERS2493861 |

| | | | | |
|---|---|---|---|---|
| ERR3772023 | Argentina | Americas | 1993 | ERS2493862 |
| ERR3772024 | Argentina | Americas | 1993 | ERS2493863 |
| ERR3772026 | Argentina | Americas | 1993 | ERS2493865 |
| ERR3772027 | Argentina | Americas | 1993 | ERS2493866 |
| ERR3772028 | Argentina | Americas | 1993 | ERS2493867 |
| ERR3772029 | Argentina | Americas | 1993 | ERS2493868 |
| ERR3772030 | Argentina | Americas | 1997 | ERS2493869 |
| ERR3772031 | Argentina | Americas | 1993 | ERS2493870 |
| ERR3772032 | Argentina | Americas | 1993 | ERS2493871 |
| ERR3772033 | Argentina | Americas | 1993 | ERS2493872 |
| ERR3772034 | Argentina | Americas | 1993 | ERS2493873 |
| ERR3772035 | Argentina | Americas | 1993 | ERS2493874 |
| ERR3772036 | Argentina | Americas | 1993 | ERS2493875 |
| ERR3772037 | Argentina | Americas | 1993 | ERS2493876 |
| ERR3772038 | Argentina | Americas | 1993 | ERS2493877 |
| ERR3772039 | Argentina | Americas | 1993 | ERS2493879 |
| ERR3772040 | Argentina | Americas | 1993 | ERS2493880 |
| ERR3772041 | Argentina | Americas | 1993 | ERS2493881 |
| ERR3772042 | Argentina | Americas | 1993 | ERS2493882 |
| ERR3772043 | Argentina | Americas | 1993 | ERS2493883 |
| ERR3772044 | Argentina | Americas | 1997 | ERS2493594 |
| ERR3772045 | Argentina | Americas | 1997 | ERS2493595 |
| ERR3772046 | Argentina | Americas | 1996 | ERS2493596 |
| ERR3772047 | Argentina | Americas | 1997 | ERS2493597 |
| ERR3772048 | Argentina | Americas | 1997 | ERS2493598 |
| ERR3772049 | Argentina | Americas | 1997 | ERS2493599 |
| ERR3772052 | Argentina | Americas | 1997 | ERS2493602 |
| ERR3772053 | Argentina | Americas | 1997 | ERS2493603 |
| ERR3772055 | Argentina | Americas | 1996 | ERS2493605 |
| ERR3772056 | Argentina | Americas | 1997 | ERS2493606 |
| ERR3772057 | Argentina | Americas | 1997 | ERS2493607 |
| ERR3772058 | Argentina | Americas | 1997 | ERS2493608 |
| ERR3772059 | Argentina | Americas | 1996 | ERS2493609 |
| ERR3772060 | Argentina | Americas | 1997 | ERS2493610 |
| ERR3772062 | Argentina | Americas | 1997 | ERS2493612 |
| ERR3772065 | Argentina | Americas | 1997 | ERS2493615 |
| ERR3772066 | Argentina | Americas | 1997 | ERS2493616 |
| ERR3772067 | Argentina | Americas | 1997 | ERS2493617 |
| ERR3772068 | Argentina | Americas | 1997 | ERS2493618 |
| ERR3772070 | Argentina | Americas | 1997 | ERS2493620 |
| ERR3772071 | Argentina | Americas | 1997 | ERS2493621 |
| ERR3772072 | Argentina | Americas | 1997 | ERS2493622 |
| ERR3772073 | Argentina | Americas | 1997 | ERS2493623 |

| | | | | |
|---|---|---|---|---|
| ERR3772075 | Argentina | Americas | 1997 | ERS2493625 |
| ERR3772076 | Argentina | Americas | 1997 | ERS2493626 |
| ERR3772077 | Argentina | Americas | 1997 | ERS2493627 |
| ERR3772078 | Argentina | Americas | 1997 | ERS2493628 |
| ERR3772079 | Argentina | Americas | 1997 | ERS2493629 |
| ERR3772080 | Argentina | Americas | 1997 | ERS2493630 |
| ERR3772081 | Argentina | Americas | 1997 | ERS2493631 |
| ERR3772082 | Argentina | Americas | 1996 | ERS2493632 |
| ERR3772087 | Argentina | Americas | 1996 | ERS2493637 |
| ERR3772088 | Argentina | Americas | 1996 | ERS2493638 |
| ERR3772089 | Argentina | Americas | 1996 | ERS2493639 |
| ERR3772090 | Argentina | Americas | 1996 | ERS2493640 |
| ERR3772091 | Argentina | Americas | 1996 | ERS2493641 |
| ERR3772092 | Argentina | Americas | 1996 | ERS2493642 |
| ERR3772095 | Argentina | Americas | 1996 | ERS2493645 |
| ERR3772096 | Argentina | Americas | 1996 | ERS2493646 |
| ERR3772099 | Argentina | Americas | 1992 | ERS2493649 |
| ERR3772101 | Argentina | Americas | 1992 | ERS2493651 |
| ERR3772102 | Argentina | Americas | 1992 | ERS2493652 |
| ERR3772103 | Argentina | Americas | 1992 | ERS2493654 |
| ERR3772104 | Argentina | Americas | 1992 | ERS2493655 |
| ERR3772106 | Argentina | Americas | 1992 | ERS2493657 |
| ERR3772107 | Argentina | Americas | 1992 | ERS2493658 |
| ERR3772110 | Argentina | Americas | 1996 | ERS2493662 |
| ERR3772114 | Argentina | Americas | 1996 | ERS2493668 |
| ERR3772116 | Argentina | Americas | 1996 | ERS2493670 |
| ERR3772119 | Argentina | Americas | 1996 | ERS2493673 |
| ERR3772120 | Argentina | Americas | 1996 | ERS2493674 |
| ERR3772122 | Argentina | Americas | 1996 | ERS2493676 |
| ERR3772123 | Argentina | Americas | 1996 | ERS2493677 |
| ERR3772126 | Argentina | Americas | 1996 | ERS2493680 |
| ERR3772128 | Argentina | Americas | 1993 | ERS2493682 |
| ERR3772129 | Argentina | Americas | 1993 | ERS2493683 |
| ERR3772131 | Argentina | Americas | 1992 | ERS2493685 |
| ERR3772134 | Argentina | Americas | 1992 | ERS2493688 |
| ERR3772135 | Argentina | Americas | 1992 | ERS2493689 |
| ERR3772136 | Argentina | Americas | 1992 | ERS2493690 |
| ERR3772138 | Argentina | Americas | 1992 | ERS2493692 |
| ERR3772139 | Argentina | Americas | 1992 | ERS2493693 |
| ERR3772140 | Argentina | Americas | 1992 | ERS2493694 |
| ERR3772141 | Argentina | Americas | 1992 | ERS2493695 |
| ERR3772142 | Argentina | Americas | 1992 | ERS2493696 |
| ERR3772145 | Argentina | Americas | 1992 | ERS2493699 |

| | | | | |
|---|---|---|---|---|
| ERR3772147 | Argentina | Americas | 1993 | ERS2493701 |
| ERR3772150 | Argentina | Americas | 1993 | ERS2493704 |
| ERR3772151 | Argentina | Americas | 1993 | ERS2493705 |
| ERR3772152 | Argentina | Americas | 1993 | ERS2493706 |
| ERR3772153 | Argentina | Americas | 1993 | ERS2493707 |
| ERR3772154 | Argentina | Americas | 1993 | ERS2493708 |
| ERR3772155 | Argentina | Americas | 1993 | ERS2493709 |
| ERR3772156 | Argentina | Americas | 1993 | ERS2493710 |
| ERR3772157 | Argentina | Americas | 1993 | ERS2493717 |
| ERR3772160 | Argentina | Americas | 1993 | ERS2493720 |
| ERR3772161 | Argentina | Americas | 1993 | ERS2493721 |
| ERR3772162 | Argentina | Americas | 1993 | ERS2493722 |
| ERR3772163 | Argentina | Americas | 1993 | ERS2493723 |
| ERR3772164 | Argentina | Americas | 1993 | ERS2493724 |
| ERR3772165 | Argentina | Americas | 1992 | ERS2493725 |
| ERR3772168 | Argentina | Americas | 1993 | ERS2493728 |
| ERR3772169 | Argentina | Americas | 1993 | ERS2493729 |
| ERR3772170 | Argentina | Americas | 1993 | ERS2493730 |
| ERR3772172 | Argentina | Americas | 1993 | ERS2493732 |
| ERR3772173 | Argentina | Americas | 1993 | ERS2493711 |
| ERR3772174 | Argentina | Americas | 1992 | ERS2493712 |
| ERR3772175 | Argentina | Americas | 1992 | ERS2493713 |
| ERR3772178 | Argentina | Americas | 1992 | ERS2493716 |
| ERR3772179 | Argentina | Americas | 1992 | ERS2493733 |
| ERR3772180 | Argentina | Americas | 1992 | ERS2493734 |
| ERR3772181 | Argentina | Americas | 1992 | ERS2493735 |
| ERR3772182 | Argentina | Americas | 1992 | ERS2493736 |
| ERR3772183 | Argentina | Americas | 1992 | ERS2493737 |
| ERR3772184 | Argentina | Americas | 1992 | ERS2493738 |
| ERR3772185 | Argentina | Americas | 1992 | ERS2493739 |
| ERR3772186 | Argentina | Americas | 1992 | ERS2493740 |
| ERR3772187 | Argentina | Americas | 1992 | ERS2493741 |
| ERR3772188 | Argentina | Americas | 1992 | ERS2493742 |
| ERR3772189 | Argentina | Americas | 1992 | ERS2493744 |
| ERR3772190 | Argentina | Americas | 1992 | ERS2493745 |
| ERR3772191 | Argentina | Americas | 1992 | ERS2493746 |
| ERR3772192 | Argentina | Americas | 1992 | ERS2493748 |
| ERR3772193 | Argentina | Americas | 1993 | ERS2493749 |
| ERR3772194 | Argentina | Americas | 1992 | ERS2493750 |
| ERR3772195 | Argentina | Americas | 1992 | ERS2493751 |
| ERR3772196 | Argentina | Americas | 1993 | ERS2493752 |
| ERR3772197 | Argentina | Americas | 1992 | ERS2493753 |
| ERR3772198 | Argentina | Americas | 1992 | ERS2493754 |

| | | | | |
|---|---|---|---|---|
| ERR3772199 | Argentina | Americas | 1992 | ERS2493755 |
| ERR3772200 | Argentina | Americas | 1992 | ERS2493756 |
| ERR3772201 | Argentina | Americas | 1992 | ERS2493757 |
| ERR3772202 | Argentina | Americas | 1992 | ERS2493758 |
| ERR3772203 | Argentina | Americas | 1992 | ERS2493759 |
| ERR3772204 | Argentina | Americas | 1992 | ERS2493760 |
| ERR3772205 | Argentina | Americas | 1992 | ERS2493761 |
| ERR3772206 | Argentina | Americas | 1992 | ERS2493762 |
| ERR3772207 | Argentina | Americas | 1992 | ERS2493763 |
| ERR3772208 | Argentina | Americas | 1992 | ERS2493764 |
| ERR3772209 | Argentina | Americas | 1992 | ERS2493765 |
| ERR3772210 | Argentina | Americas | 1992 | ERS2493766 |
| ERR3772211 | Argentina | Americas | 1992 | ERS2493767 |
| ERR3772212 | Argentina | Americas | 1993 | ERS2493768 |
| ERR3772213 | Argentina | Americas | 1993 | ERS2493769 |
| ERR3772214 | Argentina | Americas | 1992 | ERS2493770 |
| ERR3772215 | Argentina | Americas | 1993 | ERS2493772 |
| ERR3772216 | Argentina | Americas | 1992 | ERS2493773 |
| ERR3772217 | Argentina | Americas | 1992 | ERS2493776 |
| ERR3772218 | Argentina | Americas | 1992 | ERS2493777 |
| ERR3772219 | Argentina | Americas | 1993 | ERS2493778 |
| ERR3772220 | Argentina | Americas | 1992 | ERS2493780 |
| ERR3772221 | Argentina | Americas | 1993 | ERS2493782 |
| ERR3772222 | Argentina | Americas | 1993 | ERS2493784 |
| ERR3772223 | Argentina | Americas | 1993 | ERS2493786 |
| ERR3772224 | Argentina | Americas | 1993 | ERS2494039 |
| ERR3772225 | Argentina | Americas | 1993 | ERS2494040 |
| ERR3772226 | Argentina | Americas | 1993 | ERS2494041 |
| ERR3772227 | Argentina | Americas | 1993 | ERS2494042 |
| ERR3772228 | Argentina | Americas | 1993 | ERS2494043 |
| ERR3772229 | Argentina | Americas | 1993 | ERS2494044 |
| ERR3772230 | Argentina | Americas | 1993 | ERS2494045 |
| ERR3772231 | Argentina | Americas | 1993 | ERS2494046 |
| ERR3772232 | Argentina | Americas | 1993 | ERS2494047 |
| ERR3772233 | Argentina | Americas | 1993 | ERS2494048 |
| ERR3772234 | Argentina | Americas | 1993 | ERS2494049 |
| ERR3772235 | Argentina | Americas | 1993 | ERS2494050 |
| ERR3772236 | Argentina | Americas | 1993 | ERS2494051 |
| ERR3772237 | Argentina | Americas | 1993 | ERS2494052 |
| ERR3772238 | Argentina | Americas | 1993 | ERS2494053 |
| ERR3772239 | Argentina | Americas | 1993 | ERS2494054 |
| ERR3772240 | Argentina | Americas | 1993 | ERS2494055 |
| ERR3772241 | Argentina | Americas | 1993 | ERS2494056 |

| | | | | |
|---|---|---|---|---|
| ERR3772242 | Argentina | Americas | 1993 | ERS2494057 |
| ERR3772243 | Argentina | Americas | 1993 | ERS2494058 |
| ERR3772244 | Argentina | Americas | 1993 | ERS2494059 |
| ERR3772245 | Argentina | Americas | 1993 | ERS2494060 |
| ERR3772246 | Argentina | Americas | 1993 | ERS2494061 |
| ERR3772247 | Argentina | Americas | 1993 | ERS2494062 |
| ERR3772248 | Argentina | Americas | 1993 | ERS2494063 |
| ERR3772249 | Argentina | Americas | 1993 | ERS2494064 |
| ERR3772250 | Argentina | Americas | 1993 | ERS2494065 |
| ERR3772251 | Argentina | Americas | 1993 | ERS2494066 |
| ERR3772252 | Argentina | Americas | 1993 | ERS2494067 |
| ERR3772253 | Argentina | Americas | 1993 | ERS2494068 |
| ERR3772254 | Argentina | Americas | 1993 | ERS2494069 |
| ERR3772255 | Argentina | Americas | 1993 | ERS2494070 |
| ERR3772256 | Argentina | Americas | 1993 | ERS2494071 |
| ERR3772257 | Argentina | Americas | 1993 | ERS2494072 |
| ERR3772258 | Argentina | Americas | 1994 | ERS2494073 |
| ERR3772260 | Argentina | Americas | 1992 | ERS2494075 |
| ERR3772261 | Argentina | Americas | 1992 | ERS2494076 |
| ERR3772262 | Argentina | Americas | 1993 | ERS2494077 |
| ERR3772263 | Argentina | Americas | 1996 | ERS2494078 |
| ERR3772264 | Argentina | Americas | 1993 | ERS2494079 |
| ERR3772265 | Argentina | Americas | 1994 | ERS2494080 |
| ERR3772266 | Argentina | Americas | 1993 | ERS2494081 |
| ERR3772267 | Argentina | Americas | 1994 | ERS2494082 |
| ERR3772268 | Argentina | Americas | 1993 | ERS2494083 |
| ERR3772270 | Argentina | Americas | 1993 | ERS2494085 |
| ERR3772276 | Argentina | Americas | 1996 | ERS2494091 |
| ERR386629 | Democratic R | Africa | 2009 | ERR386629 |
| ERR386647 | Democratic R | Africa | 2012 | ERR386647 |
| ERR386656 | Zambia | Africa | 2012 | ERR386656 |
| ERR386658 | Guinea | Africa | 2012 | ERR386658 |
| ERR386661 | Zambia | Africa | 2012 | ERR386661 |
| ERR386663 | Zambia | Africa | 2012 | ERR386663 |
| ERR386666 | Guinea | Africa | 2012 | ERR386666 |
| ERR386672 | Democratic R | Africa | 2009 | ERR386672 |
| ERR386683 | Democratic R | Africa | 2009 | ERR386683 |
| ERR386688 | Democratic R | Africa | 2012 | ERR386688 |
| ERR386690 | Democratic R | Africa | 2009 | ERR386690 |
| ERR386695 | Democratic R | Africa | 2012 | ERR386695 |
| ERR386698 | Democratic R | Africa | 2009 | ERR386698 |
| ERR386704 | Guinea | Africa | 2012 | ERR386704 |
| ERR386711 | Democratic R | Africa | 2012 | ERR386711 |

| | | | | |
|---|---|---|---|---|
| ERR386712 | Democratic F | Africa | 2012 | ERR386712 |
| ERR466829 | Mexico | Americas | 2013 | ERR466829 |
| ERR466830 | Mexico | Americas | 2013 | ERR466830 |
| ERR466831 | Mexico | Americas | 2013 | ERR466831 |
| ERR466832 | Mexico | Americas | 2013 | ERR466832 |
| ERR466833 | Mexico | Americas | 2013 | ERR466833 |
| ERR466834 | Mexico | Americas | 2013 | ERR466834 |
| ERR466835 | Mexico | Americas | 2013 | ERR466835 |
| ERR466836 | Mexico | Americas | 2013 | ERR466836 |
| ERR466837 | Mexico | Americas | 2013 | ERR466837 |
| ERR466838 | Mexico | Americas | 2013 | ERR466838 |
| ERR466839 | Mexico | Americas | 2013 | ERR466839 |
| ERR466840 | Mexico | Americas | 2013 | ERR466840 |
| ERR466841 | Mexico | Americas | 2013 | ERR466841 |
| ERR466842 | Mexico | Americas | 2013 | ERR466842 |
| ERR466843 | Mexico | Americas | 2013 | ERR466843 |
| ERR466844 | Mexico | Americas | 2013 | ERR466844 |
| ERR466845 | Mexico | Americas | 2013 | ERR466845 |
| ERR466846 | Mexico | Americas | 2013 | ERR466846 |
| ERR466847 | Mexico | Americas | 2013 | ERR466847 |
| ERR466848 | Mexico | Americas | 2013 | ERR466848 |
| ERR471120 | Mexico | Americas | 2013 | ERR471120 |
| ERR471121 | Mexico | Americas | 2013 | ERR471121 |
| ERR471122 | Mexico | Americas | 2013 | ERR471122 |
| ERR471123 | Mexico | Americas | 2013 | ERR471123 |
| ERR471124 | Mexico | Americas | 2013 | ERR471124 |
| ERR471125 | Mexico | Americas | 2013 | ERR471125 |
| ERR471126 | Mexico | Americas | 2013 | ERR471126 |
| ERR471127 | Mexico | Americas | 2013 | ERR471127 |
| ERR471128 | Mexico | Americas | 2013 | ERR471128 |
| ERR471129 | Mexico | Americas | 2013 | ERR471129 |
| ERR572514 | Democratic F | Africa | 2009 | ERR572514 |
| ERR572538 | Democratic F | Africa | 2009 | ERR572538 |
| ERR572548 | Democratic F | Africa | 2012 | ERR572548 |
| ERR572559 | Democratic F | Africa | 2013 | ERR572559 |
| ERR572563 | Democratic F | Africa | 2013 | ERR572563 |
| ERR572571 | Togo | Africa | 2010 | ERR572571 |
| ERR572572 | Togo | Africa | 2010 | ERR572572 |
| ERR572574 | Togo | Africa | 2010 | ERR572574 |
| ERR572580 | Togo | Africa | 2010 | ERR572580 |
| ERR572582 | Togo | Africa | 2011 | ERR572582 |
| ERR572585 | Togo | Africa | 2011 | ERR572585 |
| ERR572589 | Togo | Africa | 2011 | ERR572589 |

| | | | | |
|---|---|---|---|---|
| ERR572590 | Togo | Africa | 2012 | ERR572590 |
| ERR572592 | Togo | Africa | 2012 | ERR572592 |
| ERR572756 | Democratic F | Africa | 2008 | ERR572756 |
| ERR572757 | Democratic F | Africa | 2008 | ERR572757 |
| ERR572772 | Democratic F | Africa | 2011 | ERR572772 |
| ERR572781 | Democratic F | Africa | 2011 | ERR572781 |
| ERR572807 | Democratic F | Africa | 2012 | ERR572807 |
| ERR572810 | Democratic F | Africa | 2001 | ERR572810 |
| ERR572821 | Democratic F | Africa | 2011 | ERR572821 |
| ERR572836 | Democratic F | Africa | 2013 | ERR572836 |
| ERR572837 | Democratic F | Africa | 2013 | ERR572837 |
| ERR572839 | Togo | Africa | 2010 | ERR572839 |
| ERR572840 | Togo | Africa | 2011 | ERR572840 |
| ERR572842 | Togo | Africa | 2012 | ERR572842 |
| ERR576950 | Chile | Americas | 1991 | ERR576950 |
| ERR576951 | China | Asia | 2001 | ERR576951 |
| ERR576970 | Thailand | Asia | 1990 | ERR576970 |
| ERR576971 | China | Asia | 1961 | ERR576971 |
| ERR576972 | China | Asia | 1961 | ERR576972 |
| ERR576973 | Peru | Americas | 1991 | ERR576973 |
| ERR576974 | China | Asia | 2010 | ERR576974 |
| ERR576975 | China | Asia | 2001 | ERR576975 |
| ERR576976 | China | Asia | 1998 | ERR576976 |
| ERR576977 | China | Asia | 1993 | ERR576977 |
| ERR576978 | Mauritania | Africa | 1986 | ERR576978 |
| ERR576979 | China | Asia | 1964 | ERR576979 |
| ERR576980 | China | Asia | 1963 | ERR576980 |
| ERR576981 | Indonesia | Asia | 1961 | ERR576981 |
| ERR576982 | Indonesia | Asia | 1961 | ERR576982 |
| ERR576983 | Indonesia | Asia | 1961 | ERR576983 |
| ERR576984 | China | Asia | 1999 | ERR576984 |
| ERR576985 | China | Asia | 1994 | ERR576985 |
| ERR576986 | China | Asia | 1993 | ERR576986 |
| ERR576987 | China | Asia | 1988 | ERR576987 |
| ERR576988 | China | Asia | 1984 | ERR576988 |
| ERR576995 | China | Asia | 1983 | ERR576995 |
| ERR576996 | China | Asia | 1983 | ERR576996 |
| ERR577139 | China | Asia | 1982 | ERR577139 |
| ERR577140 | China | Asia | 1981 | ERR577140 |
| ERR577141 | China | Asia | 1980 | ERR577141 |
| ERR577142 | China | Asia | 1979 | ERR577142 |
| ERR577143 | China | Asia | 1978 | ERR577143 |
| ERR579049 | China | Asia | 1977 | ERR579049 |

| | | | | |
|---|---|---|---|---|
| ERR579050 | China | Asia | 1974 | ERR579050 |
| ERR579051 | China | Asia | 1973 | ERR579051 |
| ERR579052 | China | Asia | 1969 | ERR579052 |
| ERR579053 | China | Asia | 1965 | ERR579053 |
| ERR579054 | China | Asia | 1964 | ERR579054 |
| ERR579055 | China | Asia | 1962 | ERR579055 |
| ERR579056 | China | Asia | 1966 | ERR579056 |
| ERR579057 | China | Asia | 1961 | ERR579057 |
| ERR579058 | China | Asia | 1961 | ERR579058 |
| ERR579059 | China | Asia | 1961 | ERR579059 |
| ERR579060 | China | Asia | 1961 | ERR579060 |
| ERR579061 | China | Asia | 1988 | ERR579061 |
| ERR579062 | China | Asia | 1979 | ERR579062 |
| ERR579063 | China | Asia | 1964 | ERR579063 |
| ERR579064 | China | Asia | 1987 | ERR579064 |
| ERR579065 | China | Asia | 1981 | ERR579065 |
| ERR579066 | China | Asia | 1962 | ERR579066 |
| ERR579067 | China | Asia | 1986 | ERR579067 |
| ERR579068 | China | Asia | 1995 | ERR579068 |
| ERR579069 | China | Asia | 1998 | ERR579069 |
| ERR579070 | China | Asia | 1989 | ERR579070 |
| ERR579072 | China | Asia | 1980 | ERR579072 |
| ERR579073 | China | Asia | 1996 | ERR579073 |
| ERR579074 | China | Asia | 1978 | ERR579074 |
| ERR579075 | China | Asia | 2008 | ERR579075 |
| ERR579076 | China | Asia | 2000 | ERR579076 |
| ERR579077 | China | Asia | 1998 | ERR579077 |
| ERR579078 | China | Asia | 1997 | ERR579078 |
| ERR579079 | China | Asia | 1993 | ERR579079 |
| ERR579080 | China | Asia | 1992 | ERR579080 |
| ERR579081 | China | Asia | 1991 | ERR579081 |
| ERR579082 | China | Asia | 1990 | ERR579082 |
| ERR579083 | China | Asia | 1979 | ERR579083 |
| ERR579085 | China | Asia | 2002 | ERR579085 |
| ERR579086 | China | Asia | 2001 | ERR579086 |
| ERR579087 | China | Asia | 1978 | ERR579087 |
| ERR579088 | China | Asia | 1984 | ERR579088 |
| ERR579089 | China | Asia | 2005 | ERR579089 |
| ERR579090 | China | Asia | 2008 | ERR579090 |
| ERR579091 | China | Asia | 2000 | ERR579091 |
| ERR579092 | China | Asia | 1998 | ERR579092 |
| ERR579093 | China | Asia | 1998 | ERR579093 |
| ERR579114 | China | Asia | 1998 | ERR579114 |

| | | | | |
|---|---|---|---|---|
| ERR579309 | China | Asia | 2001 | ERR579309 |
| ERR579311 | China | Asia | 1994 | ERR579311 |
| ERR579313 | China | Asia | 2001 | ERR579313 |
| ERR579919 | China | Asia | 1985 | ERR579919 |
| ERR579986 | China | Asia | 1994 | ERR579986 |
| ERR580006 | Peru | Americas | 1991 | ERR580006 |
| ERR713549( | Algeria | Africa | 2018 | ERR7135490 |
| ERR7135491 | Algeria | Africa | 2018 | ERR7135491 |
| ERR713549 | Algeria | Africa | 2018 | ERR7135492 |
| ERR713549 | Algeria | Africa | 2018 | ERR7135493 |
| ERR713549 | Algeria | Africa | 2018 | ERR7135494 |
| ERR976391 | Angola | Africa | 1990 | ERR976391 |
| ERR976392 | Malawi | Africa | 1990 | ERR976392 |
| ERR976393 | Malawi | Africa | 1990 | ERR976393 |
| ERR976394 | Malawi | Africa | 1989 | ERR976394 |
| ERR976395 | Algeria | Africa | 1987 | ERR976395 |
| ERR976396 | Angola | Africa | 1990 | ERR976396 |
| ERR976397 | Egypt | Africa | 1966 | ERR976397 |
| ERR976398 | Sierra Leone | Africa | 1970 | ERR976398 |
| ERR976399 | Ethiopia | Africa | 1970 | ERR976399 |
| ERR976400 | Cameroon | Africa | 1970 | ERR976400 |
| ERR976401 | Chad | Africa | 1971 | ERR976401 |
| ERR976402 | Mali | Africa | 1970 | ERR976402 |
| ERR976403 | Guinea | Africa | 1970 | ERR976403 |
| ERR976404 | Democratic F | Africa | 1988 | ERR976404 |
| ERR976405 | Algeria | Africa | 1983 | ERR976405 |
| ERR976406 | Algeria | Africa | 1975 | ERR976406 |
| ERR976407 | Algeria | Africa | 1982 | ERR976407 |
| ERR976408 | Algeria | Africa | 1982 | ERR976408 |
| ERR976409 | Algeria | Africa | 1986 | ERR976409 |
| ERR976410 | Democratic F | Africa | 1984 | ERR976410 |
| ERR976411 | Cote d'Ivoire | Africa | 1984 | ERR976411 |
| ERR976412 | Algeria | Africa | 1986 | ERR976412 |
| ERR976413 | Algeria | Africa | 1974 | ERR976413 |
| ERR976414 | Rwanda | Africa | 1988 | ERR976414 |
| ERR976415 | Chad | Africa | 1972 | ERR976415 |
| ERR976416 | Chad | Africa | 1972 | ERR976416 |
| ERR976418 | Burkina Fasc | Africa | 1974 | ERR976418 |
| ERR976419 | Burkina Fasc | Africa | 1974 | ERR976419 |
| ERR976420 | Cameroon | Africa | 1970 | ERR976420 |
| ERR976421 | Cameroon | Africa | 1970 | ERR976421 |
| ERR976422 | Cameroon | Africa | 1970 | ERR976422 |
| ERR976423 | Cameroon | Africa | 1970 | ERR976423 |

| | | | | |
|---|---|---|---|---|
| ERR976424 | Cameroon | Africa | 1970 | ERR976424 |
| ERR976425 | Cote d'Ivoire | Africa | 1973 | ERR976425 |
| ERR976426 | Cote d'Ivoire | Africa | 1973 | ERR976426 |
| ERR976427 | Cote d'Ivoire | Africa | 1973 | ERR976427 |
| ERR976428 | Cote d'Ivoire | Africa | 1970 | ERR976428 |
| ERR976429 | Ghana | Africa | 1971 | ERR976429 |
| ERR976430 | Ghana | Africa | 1971 | ERR976430 |
| ERR976431 | Nigeria | Africa | 1970 | ERR976431 |
| ERR976432 | Nigeria | Africa | 1970 | ERR976432 |
| ERR976433 | Senegal | Africa | 1973 | ERR976433 |
| ERR976434 | Senegal | Africa | 1974 | ERR976434 |
| ERR976435 | Senegal | Africa | 1972 | ERR976435 |
| ERR976436 | Senegal | Africa | 1971 | ERR976436 |
| ERR976437 | Chad | Africa | 1974 | ERR976437 |
| ERR976438 | Chad | Africa | 1974 | ERR976438 |
| ERR976439 | Chad | Africa | 1972 | ERR976439 |
| ERR976440 | Chad | Africa | 1974 | ERR976440 |
| ERR976441 | Tunisia | Africa | 1974 | ERR976441 |
| ERR976442 | Burkina Fasc | Africa | 1974 | ERR976442 |
| ERR976443 | Burkina Fasc | Africa | 1974 | ERR976443 |
| ERR976444 | Algeria | Africa | 1974 | ERR976444 |
| ERR976445 | Morocco | Africa | 1974 | ERR976445 |
| ERR976446 | Morocco | Africa | 1972 | ERR976446 |
| ERR976447 | Morocco | Africa | 1972 | ERR976447 |
| ERR976448 | Morocco | Africa | 1972 | ERR976448 |
| ERR976449 | Morocco | Africa | 1972 | ERR976449 |
| ERR976450 | Morocco | Africa | 1972 | ERR976450 |
| ERR976451 | Comoros | Africa | 1975 | ERR976451 |
| ERR976452 | Comoros | Africa | 1975 | ERR976452 |
| ERR976453 | Comoros | Africa | 1975 | ERR976453 |
| ERR976454 | Tunisia | Africa | 1971 | ERR976454 |
| ERR976455 | Tunisia | Africa | 1971 | ERR976455 |
| ERR976456 | Angola | Africa | 1970 | ERR976456 |
| ERR976457 | Angola | Africa | 1970 | ERR976457 |
| ERR976458 | Cote d'Ivoire | Africa | 1970 | ERR976458 |
| ERR976459 | Benin | Africa | 1970 | ERR976459 |
| ERR976460 | Benin | Africa | 1970 | ERR976460 |
| ERR976463 | Ethiopia | Africa | 1970 | ERR976463 |
| ERR976464 | Ghana | Africa | 1970 | ERR976464 |
| ERR976465 | Ghana | Africa | 1970 | ERR976465 |
| ERR976466 | Guinea | Africa | 1970 | ERR976466 |
| ERR976467 | Guinea | Africa | 1970 | ERR976467 |
| ERR976468 | Liberia | Africa | 1970 | ERR976468 |

| | | | | |
|---|---|---|---|---|
| ERR976469 | Liberia | Africa | 1970 | ERR976469 |
| ERR976470 | Libya | Africa | 1970 | ERR976470 |
| ERR976471 | Libya | Africa | 1970 | ERR976471 |
| ERR976472 | Mali | Africa | 1970 | ERR976472 |
| ERR976473 | Niger | Africa | 1970 | ERR976473 |
| ERR976474 | Niger | Africa | 1970 | ERR976474 |
| ERR976475 | Sierra Leone | Africa | 1970 | ERR976475 |
| ERR976476 | Algeria | Africa | 1990 | ERR976476 |
| ERR976477 | Sierra Leone | Africa | 1986 | ERR976477 |
| ERR976478 | Djibouti | Africa | 1985 | ERR976478 |
| ERR976479 | Burkina Fasc | Africa | 1984 | ERR976479 |
| ERR976480 | Mali | Africa | 1984 | ERR976480 |
| ERR976481 | Mali | Africa | 1985 | ERR976481 |
| ERR976482 | Djibouti | Africa | 1985 | ERR976482 |
| ERR976483 | Benin | Africa | 1985 | ERR976483 |
| ERR976484 | Democratic F | Africa | 1988 | ERR976484 |
| ERR976485 | Burkina Fasc | Africa | 1984 | ERR976485 |
| ERR976486 | Burkina Fasc | Africa | 1984 | ERR976486 |
| ERR976487 | Democratic F | Africa | 1984 | ERR976487 |
| ERR976488 | Bolivia | Americas | 1991 | ERR976488 |
| ERR976489 | Brazil | Americas | 1991 | ERR976489 |
| ERR976492 | Brazil | Americas | 1991 | ERR976492 |
| ERR976493 | Brazil | Americas | 1992 | ERR976493 |
| ERR976494 | Bolivia | Americas | 1992 | ERR976494 |
| ERR976495 | Bolivia | Americas | 1992 | ERR976495 |
| ERR976496 | Bolivia | Americas | 1992 | ERR976496 |
| ERR976497 | Bolivia | Americas | 1991 | ERR976497 |
| ERR976498 | Angola | Africa | 1988 | ERR976498 |
| ERR976499 | Angola | Africa | 1988 | ERR976499 |
| ERR976500 | Angola | Africa | 1988 | ERR976500 |
| ERR976501 | Angola | Africa | 1988 | ERR976501 |
| ERR976502 | Angola | Africa | 1988 | ERR976502 |
| ERR976503 | Angola | Africa | 1988 | ERR976503 |
| ERR976504 | Guinea Bissa | Africa | 1994 | ERR976504 |
| ERR976505 | Portugal | Europe | 1971 | ERR976505 |
| ERR976506 | Portugal | Europe | 1971 | ERR976506 |
| ERR976507 | Portugal | Europe | 1971 | ERR976507 |
| ERR976508 | Portugal | Europe | 1974 | ERR976508 |
| ERR976510 | Portugal | Europe | 1974 | ERR976510 |
| ERR976511 | Cote d'Ivoire | Africa | 1988 | ERR976511 |
| ERR976512 | Cote d'Ivoire | Africa | 1988 | ERR976512 |
| ERR976513 | Benin | Africa | 1991 | ERR976513 |
| ERR976514 | Benin | Africa | 1991 | ERR976514 |

| | | | | |
|---|---|---|---|---|
| ERR976515 | Benin | Africa | 1991 | ERR976515 |
| ERR976516 | Niger | Africa | 1991 | ERR976516 |
| ERR976517 | Morocco | Africa | 1991 | ERR976517 |
| ERR976518 | Morocco | Africa | 1991 | ERR976518 |
| ERR976519 | Côte d'Ivoire | Africa | 1991 | ERR976519 |
| ERR976520 | Morocco | Africa | 1991 | ERR976520 |
| ERR976521 | Democratic F | Africa | 1991 | ERR976521 |
| ERR976522 | Angola | Africa | 1992 | ERR976522 |
| ERR976523 | Malawi | Africa | 1992 | ERR976523 |
| ERR976524 | Democratic F | Africa | 1992 | ERR976524 |
| ERR976525 | Democratic F | Africa | 1993 | ERR976525 |
| ERR976526 | Democratic F | Africa | 1993 | ERR976526 |
| ERR976527 | Morocco | Africa | 1993 | ERR976527 |
| ERR976528 | Rwanda | Africa | 1993 | ERR976528 |
| ERR976529 | Burundi | Africa | 1993 | ERR976529 |
| ERR976530 | Burundi | Africa | 1993 | ERR976530 |
| ERR976531 | Tanzania | Africa | 1993 | ERR976531 |
| ERR976532 | Angola | Africa | 1994 | ERR976532 |
| ERR976533 | Angola | Africa | 1994 | ERR976533 |
| ERR976534 | Cote d'Ivoire | Africa | 1993 | ERR976534 |
| ERR976535 | Cote d'Ivoire | Africa | 1994 | ERR976535 |
| ERR976536 | Cote d'Ivoire | Africa | 1993 | ERR976536 |
| ERR976537 | Guinea | Africa | 1994 | ERR976537 |
| ERR976538 | Guinea | Africa | 1994 | ERR976538 |
| ERR976539 | Guinea | Africa | 1994 | ERR976539 |
| ERR976540 | Democratic F | Africa | 1994 | ERR976540 |
| ERR976541 | Morocco | Africa | 1994 | ERR976541 |
| ERR976542 | Guinea | Africa | 1994 | ERR976542 |
| ERR976543 | Liberia | Africa | 1994 | ERR976543 |
| ERR976544 | Democratic F | Africa | 1994 | ERR976544 |
| ERR976545 | Liberia | Africa | 1994 | ERR976545 |
| ERR976546 | Liberia | Africa | 1994 | ERR976546 |
| ERR976547 | Gabon | Africa | 1997 | ERR976547 |
| ERR976548 | Democratic F | Africa | 1997 | ERR976548 |
| ERR976549 | Djibouti | Africa | 1997 | ERR976549 |
| ERR976550 | Democratic F | Africa | 1997 | ERR976550 |
| ERR976551 | Djibouti | Africa | 1997 | ERR976551 |
| ERR976552 | Uganda | Africa | 1998 | ERR976552 |
| ERR976553 | Uganda | Africa | 1998 | ERR976553 |
| ERR976554 | Cameroon | Africa | 1997 | ERR976554 |
| ERR976555 | Cameroon | Africa | 1997 | ERR976555 |
| ERR976556 | Comoros | Africa | 1998 | ERR976556 |
| ERR976557 | Comoros | Africa | 1998 | ERR976557 |

| | | | | |
|---|---|---|---|---|
| ERR976558 | Comoros | Africa | 1998 | ERR976558 |
| ERR976559 | Democratic F | Africa | 1998 | ERR976559 |
| ERR976560 | Democratic F | Africa | 1998 | ERR976560 |
| ERR976561 | Cameroon | Africa | 1998 | ERR976561 |
| ERR976562 | Cameroon | Africa | 1998 | ERR976562 |
| ERR976563 | Tanzania | Africa | 1998 | ERR976563 |
| ERR976564 | Tanzania | Africa | 1998 | ERR976564 |
| ERR976565 | Djibouti | Africa | 1997 | ERR976565 |
| ERR976566 | Rwanda | Africa | 1998 | ERR976566 |
| ERR976567 | Rwanda | Africa | 1998 | ERR976567 |
| ERR976568 | Cameroon | Africa | 1998 | ERR976568 |
| ERR976569 | Rwanda | Africa | 1998 | ERR976569 |
| ERR976570 | Cameroon | Africa | 1998 | ERR976570 |
| ERR976571 | Sudan | Africa | 1998 | ERR976571 |
| ERR976572 | Burkina Fasc | Africa | 1998 | ERR976572 |
| ERR976573 | Burkina Fasc | Africa | 1998 | ERR976573 |
| ERR976574 | Guinea | Africa | 1998 | ERR976574 |
| ERR976575 | Sudan | Africa | 1998 | ERR976575 |
| ERR976576 | Cameroon | Africa | 1998 | ERR976576 |
| ERR976577 | Cameroon | Africa | 1998 | ERR976577 |
| ERR976578 | Cameroon | Africa | 1998 | ERR976578 |
| ERR976579 | Cameroon | Africa | 1998 | ERR976579 |
| ERR976580 | Sierra Leone | Africa | 1998 | ERR976580 |
| ERR976581 | Côte d'Ivoire | Africa | 1998 | ERR976581 |
| ERR976582 | Cote d'Ivoire | Africa | 1998 | ERR976582 |
| ERR976583 | Somalia | Africa | 1999 | ERR976583 |
| ERR976584 | Uganda | Africa | 1999 | ERR976584 |
| ERR976585 | Mozambique | Africa | 1999 | ERR976585 |
| ERR976586 | Mozambique | Africa | 1999 | ERR976586 |
| ERR976587 | Mozambique | Africa | 1999 | ERR976587 |
| ERR976588 | Madagascar | Africa | 1999 | ERR976588 |
| ERR976589 | Madagascar | Africa | 1999 | ERR976589 |
| ERR976590 | Madagascar | Africa | 1999 | ERR976590 |
| ERR976591 | Madagascar | Africa | 2000 | ERR976591 |
| ERR976592 | Madagascar | Africa | 2000 | ERR976592 |
| ERR976593 | Madagascar | Africa | 2000 | ERR976593 |
| ERR976594 | Madagascar | Africa | 2000 | ERR976594 |
| ERR976595 | Madagascar | Africa | 2000 | ERR976595 |
| ERR976596 | Madagascar | Africa | 2000 | ERR976596 |
| ERR976597 | Djibouti | Africa | 1994 | ERR976597 |
| ERR976598 | Djibouti | Africa | 1994 | ERR976598 |
| ERR976599 | Djibouti | Africa | 1994 | ERR976599 |
| ERR998656 | Chad | Africa | 1994 | ERR998656 |

| | | | | |
|---|---|---|---|---|
| ERR998657 | Liberia | Africa | 1994 | ERR998657 |
| ERR998658 | Liberia | Africa | 1994 | ERR998658 |
| ERR998659 | Democratic F | Africa | 1994 | ERR998659 |
| ERR998660 | Morocco | Africa | 1994 | ERR998660 |
| ERR998661 | Morocco | Africa | 1994 | ERR998661 |
| ERR998662 | Chad | Africa | 1994 | ERR998662 |
| ERR998663 | Niger | Africa | 1994 | ERR998663 |
| ERR998664 | Angola | Africa | 1995 | ERR998664 |
| ERR998665 | Angola | Africa | 1995 | ERR998665 |
| ERR998666 | Cameroon | Africa | 1995 | ERR998666 |
| ERR998667 | Democratic F | Africa | 1995 | ERR998667 |
| ERR998668 | Democratic F | Africa | 1995 | ERR998668 |
| ERR998669 | Democratic F | Africa | 1995 | ERR998669 |
| ERR998670 | Guinea | Africa | 1995 | ERR998670 |
| ERR998671 | Burkina Fasc | Africa | 1995 | ERR998671 |
| ERR998672 | Burkina Fasc | Africa | 1995 | ERR998672 |
| ERR998673 | Mali | Africa | 1995 | ERR998673 |
| ERR998674 | Mali | Africa | 1995 | ERR998674 |
| ERR998675 | Sierra Leone | Africa | 1995 | ERR998675 |
| ERR998676 | Guinea | Africa | 1995 | ERR998676 |
| ERR998677 | Guinea | Africa | 1995 | ERR998677 |
| ERR998678 | Liberia | Africa | 1995 | ERR998678 |
| ERR998679 | Liberia | Africa | 1995 | ERR998679 |
| ERR998681 | Democratic F | Africa | 1995 | ERR998681 |
| ERR998682 | Cote d'Ivoire | Africa | 1995 | ERR998682 |
| ERR998683 | Cote d'Ivoire | Africa | 1995 | ERR998683 |
| ERR998684 | Cote d'Ivoire | Africa | 1995 | ERR998684 |
| ERR998685 | Cote d'Ivoire | Africa | 1995 | ERR998685 |
| ERR998686 | Niger | Africa | 1995 | ERR998686 |
| ERR998687 | Burkina Fasc | Africa | 1995 | ERR998687 |
| ERR998688 | Burkina Fasc | Africa | 1995 | ERR998688 |
| ERR998689 | Niger | Africa | 1996 | ERR998689 |
| ERR998690 | Senegal | Africa | 1996 | ERR998690 |
| ERR998691 | Senegal | Africa | 1996 | ERR998691 |
| ERR998692 | Nigeria | Africa | 1996 | ERR998692 |
| ERR998693 | Senegal | Africa | 1996 | ERR998693 |
| ERR998694 | Democratic F | Africa | 1996 | ERR998694 |
| ERR998695 | Senegal | Africa | 1996 | ERR998695 |
| ERR998696 | Democratic F | Africa | 1996 | ERR998696 |
| ERR998697 | Democratic F | Africa | 1996 | ERR998697 |
| ERR998698 | Democratic F | Africa | 1996 | ERR998698 |
| ERR998699 | Liberia | Africa | 1996 | ERR998699 |
| ERR998700 | Chad | Africa | 1996 | ERR998700 |

| | | | | |
|---|---|---|---|---|
| ERR998701 | Chad | Africa | 1996 | ERR998701 |
| ERR998702 | Chad | Africa | 1996 | ERR998702 |
| ERR998703 | Cameroon | Africa | 1996 | ERR998703 |
| ERR998704 | Mali | Africa | 1996 | ERR998704 |
| ERR998705 | Nigeria | Africa | 1996 | ERR998705 |
| ERR998706 | Nigeria | Africa | 1996 | ERR998706 |
| ERR998707 | Nigeria | Africa | 1996 | ERR998707 |
| ERR998708 | Chad | Africa | 1996 | ERR998708 |
| ERR998709 | Nigeria | Africa | 1996 | ERR998709 |
| ERR998710 | Senegal | Africa | 1996 | ERR998710 |
| ERR998711 | Chad | Africa | 1996 | ERR998711 |
| ERR998712 | Cameroon | Africa | 1996 | ERR998712 |
| ERR998713 | Rwanda | Africa | 1996 | ERR998713 |
| ERR998714 | Democratic F | Africa | 1996 | ERR998714 |
| ERR998715 | Rwanda | Africa | 1996 | ERR998715 |
| ERR998716 | Rwanda | Africa | 1996 | ERR998716 |
| ERR998717 | Senegal | Africa | 1996 | ERR998717 |
| ERR998718 | Senegal | Africa | 1996 | ERR998718 |
| ERR998719 | Senegal | Africa | 1996 | ERR998719 |
| ERR998720 | Senegal | Africa | 1996 | ERR998720 |
| ERR998721 | Rwanda | Africa | 1996 | ERR998721 |
| ERR998722 | Rwanda | Africa | 1996 | ERR998722 |
| ERR998723 | Democratic F | Africa | 1997 | ERR998723 |
| ERR998724 | Democratic F | Africa | 1997 | ERR998724 |
| ERR998725 | Cameroon | Africa | 1997 | ERR998725 |
| ERR998726 | Cameroon | Africa | 1997 | ERR998726 |
| ERR998727 | Cameroon | Africa | 1997 | ERR998727 |
| ERR998728 | Cameroon | Africa | 1997 | ERR998728 |
| ERR998729 | Democratic F | Africa | 1997 | ERR998729 |
| ERR998730 | Democratic F | Africa | 1997 | ERR998730 |
| ERR998731 | Cameroon | Africa | 1997 | ERR998731 |
| ERR998732 | Somalia | Africa | 1997 | ERR998732 |
| ERR998733 | Democratic F | Africa | 1997 | ERR998733 |
| ERR998734 | Democratic F | Africa | 1997 | ERR998734 |
| ERR998735 | Central Afric | Africa | 1997 | ERR998735 |
| ERR998736 | Central Afric | Africa | 1997 | ERR998736 |
| ERR998737 | Central Afric | Africa | 1997 | ERR998737 |
| ERR998738 | Central Afric | Africa | 1997 | ERR998738 |
| ERR998739 | Central Afric | Africa | 1997 | ERR998739 |
| ERR998740 | Central Afric | Africa | 1997 | ERR998740 |
| ERR998741 | Democratic F | Africa | 1997 | ERR998741 |
| ERR998742 | Central Afric | Africa | 1997 | ERR998742 |
| ERR998743 | Central Afric | Africa | 1997 | ERR998743 |

| | | | | |
|---|---|---|---|---|
| ERR998744 | Central Afric | Africa | 1997 | ERR998744 |
| ERR998745 | Central Afric | Africa | 1997 | ERR998745 |
| ERR998746 | Democratic F | Africa | 1997 | ERR998746 |
| ERR998747 | Central Afric | Africa | 1997 | ERR998747 |
| ERR998748 | Central Afric | Africa | 1997 | ERR998748 |
| ERR998749 | Benin | Africa | 1997 | ERR998749 |
| ERR998750 | Benin | Africa | 1997 | ERR998750 |
| HC-07 | Haiti | Americas | 2012 | JSTL00000000 |
| HC-08 | Haiti | Americas | 2012 | JSTM00000000 |
| HC-10 | Haiti | Americas | 2012 | JSTN00000000 |
| HC-11 | Haiti | Americas | 2012 | JSTO00000000 |
| HC-12 | Haiti | Americas | 2012 | JSTP00000000 |
| HC-15 | Haiti | Americas | 2012 | JSTQ00000000 |
| HC-16 | Haiti | Americas | 2012 | JSTR00000000 |
| HC-17 | Haiti | Americas | 2012 | JSTS00000000 |
| HC-18 | Haiti | Americas | 2012 | JSTT00000000 |
| HC-19 | Haiti | Americas | 2012 | JSTU00000000 |
| HC-21 | Haiti | Americas | 2012 | JSTV00000000 |
| HC-22 | Haiti | Americas | 2012 | JSTW00000000 |
| HC-24 | Haiti | Americas | 2012 | JSTX00000000 |
| HC-31 | Haiti | Americas | 2012 | JSTZ00000000 |
| HC-32 | Haiti | Americas | 2012 | JSUA00000000 |
| HC-33 | Haiti | Americas | 2012 | JSUB00000000 |
| HC-34 | Haiti | Americas | 2012 | JSUC00000000 |
| HC-35 | Haiti | Americas | 2012 | JSUD00000000 |
| INDRE91_1 | Mexico | Americas | 1991 | GCA_000176435.1 |
| M66 | Indonesia | Asia | 1937 | CP001233/CP001234 |
| MJ1236 | Bangladesh | Asia | 1994 | CP001485/CP001486 |
| N16961 | Bangladesh | Asia | 1975 | GCA_900205735 |
| RC9 | Kenya | Africa | 1985 | ACHX00000000 |
| SAMN0874 | Uganda | Africa | 2016 | SAMN08744330 |
| SAMN0874 | Uganda | Africa | 2016 | SAMN08744331 |
| SAMN0874 | Uganda | Africa | 2015 | SAMN08744332 |
| SRR1519251 | Democratic F | Africa | 2017 | SRR15192510 |
| SRR1519251 | Democratic F | Africa | 2017 | SRR15192511 |
| SRR1519251 | Democratic F | Africa | 2017 | SRR15192512 |
| SRR1519251 | Democratic F | Africa | 2017 | SRR15192513 |
| SRR1519251 | Democratic F | Africa | 2017 | SRR15192514 |
| SRR1519251 | Democratic F | Africa | 2017 | SRR15192515 |
| SRR1519251 | Democratic F | Africa | 2017 | SRR15192516 |
| SRR1519251 | Democratic F | Africa | 2017 | SRR15192517 |
| SRR1519251 | Democratic F | Africa | 2017 | SRR15192518 |
| SRR1519251 | Democratic F | Africa | 2017 | SRR15192519 |

| | | | | |
|---|---|---|---|---|
| SRR1519252 | Democratic F | Africa | 2017 | SRR15192520 |
| SRR1519252 | Democratic F | Africa | 2017 | SRR15192521 |
| SRR1519252 | Democratic F | Africa | 2017 | SRR15192522 |
| SRR1519252 | Democratic F | Africa | 2017 | SRR15192523 |
| SRR1519252 | Democratic F | Africa | 2017 | SRR15192524 |
| SRR1519252 | Democratic F | Africa | 2017 | SRR15192525 |
| SRR1519252 | Democratic F | Africa | 2017 | SRR15192526 |
| SRR1519252 | Democratic F | Africa | 2017 | SRR15192527 |
| SRR1519252 | Democratic F | Africa | 2017 | SRR15192528 |
| SRR1519252 | Democratic F | Africa | 2017 | SRR15192529 |
| SRR1519253 | Democratic F | Africa | 2017 | SRR15192530 |
| SRR1519253 | Democratic F | Africa | 2017 | SRR15192531 |
| SRR1519253 | Democratic F | Africa | 2017 | SRR15192532 |
| SRR1519253 | Democratic F | Africa | 2017 | SRR15192533 |
| SRR1728307 | Zambia | Africa | 2016 | SRR17283076 |
| SRR2107446 | Democratic F | Africa | 2022 | SRR21074466 |
| SRR2107446 | Democratic F | Africa | 2022 | SRR21074467 |
| SRR227303 | Bangladesh | Asia | 2010 | SRR227303 |
| SRR227307 | Zambia | Africa | 2004 | SRR227307 |
| SRR227309 | Zambia | Africa | 2004 | SRR227309 |
| SRR227311 | Zimbabwe | Africa | 2009 | SRR227311 |
| SRR227312 | Mexico | Americas | 1991 | SRR227312 |
| SRR227318 | Mexico | Americas | 2008 | SRR227318 |
| SRR227324 | Thailand | Asia | 2010 | SRR227324 |
| SRR227335 | Bangladesh | Asia | 2010 | SRR227335 |
| SRR227336 | Peru | Americas | 1991 | SRR227336 |
| SRR308665 | Nepal | Asia | 2010 | SRR308665 |
| SRR308690 | Nepal | Asia | 2010 | SRR308690 |
| SRR308691 | Nepal | Asia | 2010 | SRR308691 |
| SRR308692 | Nepal | Asia | 2010 | SRR308692 |
| SRR308693 | Nepal | Asia | 2010 | SRR308693 |
| SRR308703 | Nepal | Asia | 2010 | SRR308703 |
| SRR308704 | Nepal | Asia | 2010 | SRR308704 |
| SRR308705 | Nepal | Asia | 2010 | SRR308705 |
| SRR308706 | Nepal | Asia | 2010 | SRR308706 |
| SRR308707 | Nepal | Asia | 2010 | SRR308707 |
| SRR308708 | Nepal | Asia | 2010 | SRR308708 |
| SRR308709 | Nepal | Asia | 2010 | SRR308709 |
| SRR308713 | Nepal | Asia | 2010 | SRR308713 |
| SRR308715 | Nepal | Asia | 2010 | SRR308715 |
| SRR308716 | Nepal | Asia | 2010 | SRR308716 |
| SRR308717 | Nepal | Asia | 2010 | SRR308717 |
| SRR308720 | Nepal | Asia | 2010 | SRR308720 |

| | | | | |
|---|---|---|---|---|
| SRR308721 | Nepal | Asia | 2010 | SRR308721 |
| SRR308722 | Nepal | Asia | 2010 | SRR308722 |
| SRR308723 | Nepal | Asia | 2010 | SRR308723 |
| SRR308724 | Nepal | Asia | 2010 | SRR308724 |
| SRR308725 | Nepal | Asia | 2010 | SRR308725 |
| SRR308726 | Nepal | Asia | 2010 | SRR308726 |
| SRR308727 | Nepal | Asia | 2010 | SRR308727 |
| SRR350623 | Mexico | Americas | 2000 | SRR350623 |
| SRR491124 | Bangladesh | Asia | 2010 | SRR491124 |
| SRR491132 | Bangladesh | Asia | 2010 | SRR491132 |
| SRR491153 | Bangladesh | Asia | 2011 | SRR491153 |
| SRR491158 | Bangladesh | Asia | 2010 | SRR491158 |
| SRR491161 | Brazil | Americas | 1992 | SRR491161 |
| SRR491169 | Bangladesh | Asia | 2010 | SRR491169 |
| SRR491170 | Bangladesh | Asia | 2010 | SRR491170 |
| SRR491171 | Bangladesh | Asia | 2010 | SRR491171 |
| SRR491172 | Nepal | Asia | 2003 | SRR491172 |
| SRR491173 | Nepal | Asia | 2003 | SRR491173 |
| SRR491175 | Bangladesh | Asia | 2010 | SRR491175 |
| SRR491176 | Bangladesh | Asia | 2010 | SRR491176 |
| SRR491177 | Bangladesh | Asia | 2010 | SRR491177 |
| SRR491178 | Bangladesh | Asia | 2010 | SRR491178 |
| SRR495447 | Bangladesh | Asia | 2010 | SRR495447 |
| SRR495764 | Bangladesh | Asia | 2010 | SRR495764 |
| SRR770779 | Haiti | Americas | 2010 | SRX248208 |
| SRR771214 | Haiti | Americas | 2010 | SRX248274 |
| SRR771222 | Haiti | Americas | 2010 | SRX248275 |
| SRR771360 | Haiti | Americas | 2010 | SRX248280 |
| SRR771582 | Haiti | Americas | 2010 | SRX248403 |
| SRR771645 | Haiti | Americas | 2010 | SRX248599 |
| SRR772254 | Haiti | Americas | 2010 | SRR772254 |
| SRR772256 | Haiti | Americas | 2010 | SRX249015 |
| SRR772892 | Haiti | Americas | 2011 | SRX249313 |
| SRR772893 | Haiti | Americas | 2011 | SRX249314 |
| SRR773027 | Haiti | Americas | 2011 | SRX249334 |
| SRR773028 | Haiti | Americas | 2011 | SRX249335 |
| SRR773104 | Haiti | Americas | 2011 | SRX249353 |
| SRR773107 | Haiti | Americas | 2011 | SRX249364 |
| SRR773175 | Haiti | Americas | 2011 | SRX249365 |
| SRR773179 | Haiti | Americas | 2011 | SRX249366 |
| SRR773315 | Haiti | Americas | 2011 | SRX249382 |
| SRR773317 | Haiti | Americas | 2011 | SRX249390 |
| SRR773321 | Haiti | Americas | 2011 | SRX249393 |

| | | | | |
|---|---|---|---|---|
| SRR773389 | Haiti | Americas | 2011 | SRX249394 |
| SRR773393 | Haiti | Americas | 2011 | SRX249405 |
| SRR773397 | Haiti | Americas | 2012 | SRX249408 |
| SRR773655 | Cameroon | Africa | 2010 | SRR773655 |
| SRR773656 | Haiti | Americas | 2010 | SRX249464 |
| SRR773657 | Haiti | Americas | 2010 | SRX249465 |
| SRR773658 | Haiti | Americas | 2010 | SRX249466 |
| SRR773660 | Haiti | Americas | 2010 | SRX249468 |
| SRR774784 | South Africa | Africa | 2009 | SRR774784 |
| SRR8364252 | Haiti | Americas | 2013 | SRR8364252 |
| SRR8364253 | Haiti | Americas | 2013 | SRR8364253 |
| SRR8364254 | Haiti | Americas | 2013 | SRR8364254 |
| SRR8364255 | Haiti | Americas | 2014 | SRR8364255 |
| SRR8364256 | Haiti | Americas | 2014 | SRR8364256 |
| SRR8364257 | Haiti | Americas | 2013 | SRR8364257 |
| SRR8364258 | Haiti | Americas | 2013 | SRR8364258 |
| SRR8364259 | Haiti | Americas | 2015 | SRR8364259 |
| SRR8364260 | Haiti | Americas | 2015 | SRR8364260 |
| SRR8364261 | Haiti | Americas | 2015 | SRR8364261 |
| SRR8364262 | Haiti | Americas | 2013 | SRR8364262 |
| SRR8364263 | Haiti | Americas | 2013 | SRR8364263 |
| SRR8364264 | Haiti | Americas | 2013 | SRR8364264 |
| SRR8364265 | Haiti | Americas | 2013 | SRR8364265 |
| SRR8364266 | Haiti | Americas | 2013 | SRR8364266 |
| SRR8364267 | Haiti | Americas | 2013 | SRR8364267 |
| SRR8364268 | Haiti | Americas | 2013 | SRR8364268 |
| SRR8364269 | Haiti | Americas | 2013 | SRR8364269 |
| SRR8364270 | Haiti | Americas | 2013 | SRR8364270 |
| SRR8364271 | Haiti | Americas | 2013 | SRR8364271 |
| SRR8364272 | Haiti | Americas | 2016 | SRR8364272 |
| SRR8364273 | Haiti | Americas | 2016 | SRR8364273 |
| SRR8364274 | Haiti | Americas | 2016 | SRR8364274 |
| SRR8364275 | Haiti | Americas | 2016 | SRR8364275 |
| SRR8364276 | Haiti | Americas | 2016 | SRR8364276 |
| SRR8364277 | Haiti | Americas | 2016 | SRR8364277 |
| SRR8364278 | Haiti | Americas | 2016 | SRR8364278 |
| SRR8364279 | Haiti | Americas | 2016 | SRR8364279 |
| SRR8364280 | Haiti | Americas | 2016 | SRR8364280 |
| SRR8364281 | Haiti | Americas | 2016 | SRR8364281 |
| SRR8364282 | Haiti | Americas | 2016 | SRR8364282 |
| SRR8364283 | Haiti | Americas | 2016 | SRR8364283 |
| SRR8364284 | Haiti | Americas | 2016 | SRR8364284 |
| SRR8364285 | Haiti | Americas | 2016 | SRR8364285 |

| | | | | |
|---|---|---|---|---|
| SRR8364286 | Haiti | Americas | 2016 | SRR8364286 |
| SRR8364287 | Haiti | Americas | 2016 | SRR8364287 |
| SRR8364288 | Haiti | Americas | 2013 | SRR8364288 |
| SRR8364289 | Haiti | Americas | 2013 | SRR8364289 |
| SRR8364290 | Haiti | Americas | 2017 | SRR8364290 |
| SRR8364291 | Haiti | Americas | 2017 | SRR8364291 |
| SRR8364292 | Haiti | Americas | 2017 | SRR8364292 |
| SRR8364293 | Haiti | Americas | 2017 | SRR8364293 |
| SRR8364294 | Haiti | Americas | 2013 | SRR8364294 |
| SRR8364295 | Haiti | Americas | 2013 | SRR8364295 |
| SRR8364296 | Haiti | Americas | 2017 | SRR8364296 |
| SRR8364297 | Haiti | Americas | 2017 | SRR8364297 |
| SRR8364298 | Haiti | Americas | 2013 | SRR8364298 |
| SRR8364299 | Haiti | Americas | 2013 | SRR8364299 |
| SRR8364300 | Haiti | Americas | 2017 | SRR8364300 |
| SRR8364301 | Haiti | Americas | 2017 | SRR8364301 |
| SRR8364302 | Haiti | Americas | 2017 | SRR8364302 |
| SRR8364303 | Haiti | Americas | 2017 | SRR8364303 |
| SRR8364304 | Haiti | Americas | 2017 | SRR8364304 |
| SRR8364305 | Haiti | Americas | 2017 | SRR8364305 |
| SRR8364306 | Haiti | Americas | 2017 | SRR8364306 |
| SRR8364307 | Haiti | Americas | 2017 | SRR8364307 |
| SRR8364308 | Haiti | Americas | 2016 | SRR8364308 |
| SRR8364309 | Haiti | Americas | 2013 | SRR8364309 |
| SRR8364310 | Haiti | Americas | 2013 | SRR8364310 |
| SRR8364311 | Haiti | Americas | 2013 | SRR8364311 |
| SRR8364312 | Haiti | Americas | 2013 | SRR8364312 |
| SRR8364313 | Haiti | Americas | 2013 | SRR8364313 |
| SRR8364314 | Haiti | Americas | 2013 | SRR8364314 |
| SRR8364315 | Haiti | Americas | 2013 | SRR8364315 |
| SRR8364316 | Haiti | Americas | 2013 | SRR8364316 |
| SRR8364317 | Haiti | Americas | 2013 | SRR8364317 |
| SRR8364318 | Haiti | Americas | 2013 | SRR8364318 |
| SRR8364319 | Haiti | Americas | 2016 | SRR8364319 |
| SRR8364320 | Haiti | Americas | 2016 | SRR8364320 |
| SRR8364321 | Haiti | Americas | 2016 | SRR8364321 |
| SRR8364322 | Haiti | Americas | 2016 | SRR8364322 |
| SRR8364323 | Haiti | Americas | 2016 | SRR8364323 |
| SRR8364324 | Haiti | Americas | 2016 | SRR8364324 |
| SRR8364325 | Haiti | Americas | 2017 | SRR8364325 |
| SRR8364326 | Haiti | Americas | 2017 | SRR8364326 |
| SRR8364327 | Haiti | Americas | 2016 | SRR8364327 |
| SRR8364328 | Haiti | Americas | 2016 | SRR8364328 |

| | | | | |
|---|---|---|---|---|
| SRR8364329 | Haiti | Americas | 2016 | SRR8364329 |
| SRR8364330 | Haiti | Americas | 2017 | SRR8364330 |
| SRR8364331 | Haiti | Americas | 2016 | SRR8364331 |
| SRR8364332 | Haiti | Americas | 2017 | SRR8364332 |
| SRR8364333 | Haiti | Americas | 2017 | SRR8364333 |
| SRR8364334 | Haiti | Americas | 2017 | SRR8364334 |
| SRR8364335 | Haiti | Americas | 2017 | SRR8364335 |
| SRR8364336 | Haiti | Americas | 2016 | SRR8364336 |
| SRR8364337 | Haiti | Americas | 2013 | SRR8364337 |
| SRR8364338 | Haiti | Americas | 2013 | SRR8364338 |
| SRR8364339 | Haiti | Americas | 2013 | SRR8364339 |
| SRR8364340 | Haiti | Americas | 2013 | SRR8364340 |
| SRR8364341 | Haiti | Americas | 2013 | SRR8364341 |
| SRR8364342 | Haiti | Americas | 2013 | SRR8364342 |
| SRR8364343 | Haiti | Americas | 2013 | SRR8364343 |
| SRR8364344 | Haiti | Americas | 2013 | SRR8364344 |
| SRR8364345 | Haiti | Americas | 2013 | SRR8364345 |
| SRR8364346 | Haiti | Americas | 2013 | SRR8364346 |
| SRR8364347 | Haiti | Americas | 2014 | SRR8364347 |
| SRR8364348 | Haiti | Americas | 2014 | SRR8364348 |
| SRR8364349 | Haiti | Americas | 2014 | SRR8364349 |
| SRR8364350 | Haiti | Americas | 2014 | SRR8364350 |
| SRR8364351 | Haiti | Americas | 2014 | SRR8364351 |
| SRR8364352 | Haiti | Americas | 2014 | SRR8364352 |
| SRR8364353 | Haiti | Americas | 2015 | SRR8364353 |
| SRR8364354 | Haiti | Americas | 2014 | SRR8364354 |
| SRR8364355 | Haiti | Americas | 2015 | SRR8364355 |
| SRR8364356 | Haiti | Americas | 2015 | SRR8364356 |
| SRR8364357 | Haiti | Americas | 2013 | SRR8364357 |
| SRR8364358 | Haiti | Americas | 2013 | SRR8364358 |
| SRR8364359 | Haiti | Americas | 2017 | SRR8364359 |
| SRR8364360 | Haiti | Americas | 2017 | SRR8364360 |
| SRR8364361 | Haiti | Americas | 2017 | SRR8364361 |
| SRR8364362 | Haiti | Americas | 2017 | SRR8364362 |
| SRR8364363 | Haiti | Americas | 2017 | SRR8364363 |
| SRR8364364 | Haiti | Americas | 2017 | SRR8364364 |
| SRR8364365 | Haiti | Americas | 2017 | SRR8364365 |
| SRR8364366 | Haiti | Americas | 2017 | SRR8364366 |
| SRR8364367 | Haiti | Americas | 2017 | SRR8364367 |
| SRR8364368 | Haiti | Americas | 2017 | SRR8364368 |
| SRR8364369 | Haiti | Americas | 2013 | SRR8364369 |
| SRR8364370 | Haiti | Americas | 2013 | SRR8364370 |
| SRR8364371 | Haiti | Americas | 2013 | SRR8364371 |

| | | | | |
|---|---|---|---|---|
| SRR8364372 | Haiti | Americas | 2013 | SRR8364372 |
| SRR8364373 | Haiti | Americas | 2014 | SRR8364373 |
| SRR8364374 | Haiti | Americas | 2014 | SRR8364374 |
| SRR8364375 | Haiti | Americas | 2014 | SRR8364375 |
| SRR8364376 | Haiti | Americas | 2014 | SRR8364376 |
| SRR8364377 | Haiti | Americas | 2014 | SRR8364377 |
| SRR8364378 | Haiti | Americas | 2014 | SRR8364378 |
| SRR8364379 | Haiti | Americas | 2016 | SRR8364379 |
| SRR8364380 | Haiti | Americas | 2017 | SRR8364380 |
| SRR8364381 | Haiti | Americas | 2017 | SRR8364381 |
| SRR8364382 | Haiti | Americas | 2017 | SRR8364382 |
| SRR8364383 | Haiti | Americas | 2017 | SRR8364383 |
| SRR8364384 | Haiti | Americas | 2017 | SRR8364384 |
| SRR8364385 | Haiti | Americas | 2017 | SRR8364385 |
| SRR8364386 | Haiti | Americas | 2017 | SRR8364386 |
| SRR8364387 | Haiti | Americas | 2017 | SRR8364387 |
| SRR8364388 | Haiti | Americas | 2017 | SRR8364388 |
| SRR8364389 | Haiti | Americas | 2017 | SRR8364389 |
| SRR8364390 | Haiti | Americas | 2015 | SRR8364390 |
| SRR8364391 | Haiti | Americas | 2015 | SRR8364391 |
| SRR8364392 | Haiti | Americas | 2015 | SRR8364392 |
| SRR8364393 | Haiti | Americas | 2015 | SRR8364393 |
| SRR8364394 | Haiti | Americas | 2015 | SRR8364394 |
| SRR8364395 | Haiti | Americas | 2015 | SRR8364395 |
| SRR8364396 | Haiti | Americas | 2015 | SRR8364396 |
| SRR8364397 | Haiti | Americas | 2015 | SRR8364397 |
| SRR8364398 | Haiti | Americas | 2015 | SRR8364398 |
| SRR8364399 | Haiti | Americas | 2015 | SRR8364399 |
| SRR8364400 | Haiti | Americas | 2016 | SRR8364400 |
| SRR8364401 | Haiti | Americas | 2016 | SRR8364401 |
| SRR8364402 | Haiti | Americas | 2017 | SRR8364402 |
| SRR8364403 | Haiti | Americas | 2017 | SRR8364403 |
| SRR8364404 | Haiti | Americas | 2017 | SRR8364404 |
| SRR8364405 | Haiti | Americas | 2017 | SRR8364405 |
| SRR8364406 | Haiti | Americas | 2017 | SRR8364406 |
| SRR8364407 | Haiti | Americas | 2017 | SRR8364407 |
| SRR8364408 | Haiti | Americas | 2017 | SRR8364408 |
| SRR8364409 | Haiti | Americas | 2017 | SRR8364409 |
| SRR8364410 | Haiti | Americas | 2017 | SRR8364410 |
| SRR8364411 | Haiti | Americas | 2017 | SRR8364411 |
| SRR8364412 | Haiti | Americas | 2016 | SRR8364412 |
| SRR8364413 | Haiti | Americas | 2015 | SRR8364413 |
| SRR8364414 | Haiti | Americas | 2015 | SRR8364414 |

| | | | | |
|---|---|---|---|---|
| SRR8364415 | Haiti | Americas | 2015 | SRR8364415 |
| SRR8364416 | Haiti | Americas | 2015 | SRR8364416 |
| SRR8364417 | Haiti | Americas | 2015 | SRR8364417 |
| SRR8364418 | Haiti | Americas | 2015 | SRR8364418 |
| SRR8364419 | Haiti | Americas | 2015 | SRR8364419 |
| SRR8364420 | Haiti | Americas | 2015 | SRR8364420 |
| SRR8364421 | Haiti | Americas | 2015 | SRR8364421 |
| SRR8364422 | Haiti | Americas | 2015 | SRR8364422 |
| SRR8364423 | Haiti | Americas | 2016 | SRR8364423 |
| SRR8364424 | Haiti | Americas | 2016 | SRR8364424 |
| SRR8364425 | Haiti | Americas | 2015 | SRR8364425 |
| SRR8364426 | Haiti | Americas | 2015 | SRR8364426 |
| SRR8364427 | Haiti | Americas | 2015 | SRR8364427 |
| SRR8364428 | Haiti | Americas | 2015 | SRR8364428 |
| SRR8364429 | Haiti | Americas | 2015 | SRR8364429 |
| SRR8364430 | Haiti | Americas | 2013 | SRR8364430 |
| SRR8364431 | Haiti | Americas | 2015 | SRR8364431 |
| SRR8364432 | Haiti | Americas | 2015 | SRR8364432 |
| SRR8364433 | Haiti | Americas | 2015 | SRR8364433 |
| SRR8364434 | Haiti | Americas | 2013 | SRR8364434 |
| SRR8364435 | Haiti | Americas | 2015 | SRR8364435 |
| SRR8364436 | Haiti | Americas | 2013 | SRR8364436 |
| SRR8364437 | Haiti | Americas | 2013 | SRR8364437 |
| SRR8364438 | Haiti | Americas | 2015 | SRR8364438 |
| SRR8364439 | Haiti | Americas | 2015 | SRR8364439 |
| SRR8364440 | Haiti | Americas | 2015 | SRR8364440 |
| SRR8364441 | Haiti | Americas | 2015 | SRR8364441 |
| SRR8364442 | Haiti | Americas | 2015 | SRR8364442 |
| SRR8364443 | Haiti | Americas | 2015 | SRR8364443 |
| SRR8364444 | Haiti | Americas | 2015 | SRR8364444 |
| SRR8364445 | Haiti | Americas | 2015 | SRR8364445 |
| SRR8364446 | Haiti | Americas | 2015 | SRR8364446 |
| SRR8364447 | Haiti | Americas | 2015 | SRR8364447 |
| SRR8364448 | Haiti | Americas | 2017 | SRR8364448 |
| SRR8364449 | Haiti | Americas | 2017 | SRR8364449 |
| SRR8364450 | Haiti | Americas | 2017 | SRR8364450 |
| SRR8364451 | Haiti | Americas | 2017 | SRR8364451 |
| SRR8364452 | Haiti | Americas | 2017 | SRR8364452 |
| SRR8364453 | Haiti | Americas | 2017 | SRR8364453 |
| VCN3833 | Haiti | Americas | 2022 | VCN3833 |
| VCN3834 | Haiti | Americas | 2022 | VCN3834 |
| VCO190256 | Haiti | Americas | 2018 | VCO1902563 |
| VCO190256 | Haiti | Americas | 2018 | VCO1902564 |

| | | | | |
|---|---|---|---|---|
| VCO190256 | Haiti | Americas | 2018 | VCO1902565 |
| VCO190256 | Haiti | Americas | 2018 | VCO1902566 |
| VCO190256 | Haiti | Americas | 2018 | VCO1902567 |
| VCO190256 | Haiti | Americas | 2018 | VCO1902568 |
| VCO190256 | Haiti | Americas | 2018 | VCO1902569 |
| VCO190257 | Haiti | Americas | 2018 | VCO1902570 |
| VCO190257 | Haiti | Americas | 2018 | VCO1902571 |
| VCO190257 | Haiti | Americas | 2018 | VCO1902572 |
| VCO190257 | Haiti | Americas | 2018 | VCO1902573 |
| VCO190257 | Haiti | Americas | 2018 | VCO1902574 |
| VCO190257 | Haiti | Americas | 2018 | VCO1902575 |
| VCO190257 | Haiti | Americas | 2018 | VCO1902576 |
| VCO190257 | Haiti | Americas | 2018 | VCO1902577 |
| VCO190257 | Haiti | Americas | 2018 | VCO1902578 |
| VCO190257 | Haiti | Americas | 2018 | VCO1902579 |
| VCO190258 | Haiti | Americas | 2018 | VCO1902580 |
| VCO190258 | Haiti | Americas | 2018 | VCO1902581 |
| VCO190258 | Haiti | Americas | 2018 | VCO1902582 |
| VCO190258 | Haiti | Americas | 2018 | VCO1902583 |
| VCO190258 | Haiti | Americas | 2018 | VCO1902584 |
| VCO190258 | Haiti | Americas | 2018 | VCO1902585 |
| VCO190258 | Haiti | Americas | 2018 | VCO1902586 |
| VCO190258 | Haiti | Americas | 2018 | VCO1902587 |
| VCO190258 | Haiti | Americas | 2018 | VCO1902588 |
| VCO190258 | Haiti | Americas | 2018 | VCO1902589 |
| VCO190259 | Haiti | Americas | 2018 | VCO1902590 |
| VCO990258 | Haiti | Americas | 2018 | VCO9902589 |

**Table S6.**

List of SNPs that differentiate VCN3833 and VCN38334 from EnvJ515 environmental Ogawa strain. All strains SNPs were compared to the genome of *Vibrio cholerae* O1 2010EL-1786 (used as reference strain).

| Gene | Chr | Genome position | Mutation Type | Mutation (AA) | Reference allele | Alternative allele | VCN3833 | VCN3834 | EnvJ515 |
|---|---|---|---|---|---|---|---|---|---|
| **-** | 1 | 62877 | intergenic | | A | G | 1 | 0 | 0 |
| *istA* | 1 | 150844 | nonsynonymous | Ala231Val | G | A | 0 | 0 | 1 |
| *vpsE* | 1 | 460621 | nonsynonymous | His34Tyr | C | T | 1 | 1 | 0 |
| *moA* | 1 | 571070 | nonsynonymous | Thr310Asn | C | A | 1 | - | 0 |
| *fadJ* | 1 | 594746 | nonsynonymous | Ala517Val | C | T | 1 | 1 | 0 |
| *carS* | 1 | 879944 | nonsynonymous | Trp233Leu | C | A | 1 | 0 | 0 |
| **-** | 1 | 1042459 | intergenic | | GAAATCAA | G | 0 | 1 | 0 |
| **-** | 1 | 1465310 | intergenic | | TA | T | 1 | - | 0 |
| *mfd* | 1 | 1503970 | nonsynonymous | Val709Ala | T | C | 1 | 1 | 0 |
| HJ37_RS07360 | 1 | 1658252 | frameshift | Ala74fs | C | CGGCG | 1 | 1 | 0 |
| **-** | 1 | 1802518 | intergenic | | CA | C | 0 | - | 1 |
| *ftsY* | 1 | 2417048 | inframe deletion | Glu67_Thr68del | GCTGTTT | G | 1 | - | 0 |
| *rpsG* | 1 | 2803192 | synonymous | Glu63Glu | A | G | 1 | 1 | 0 |
| *tuf* | 1 | 2806713 | synonymous | Asp315Asp | T | C | 0 | 0 | 1 |
| HJ37_RS14100 | 2 | 30347 | inframe deletion | Pro345_Glu372del | TACCAGAACCAGAACCAGAACCAGAACCAGAACCAGAACCAGAACCAGAACCAGAACCAGAACCAGAACCAGAACCAGAACCAGA | T | 1 | - | 0 |
| **-** | 2 | 49152 | intergenic | | C | CA | 1 | 1 | 0 |

| | | | | | | | | | |
|---|---|---|---|---|---|---|---|---|---|
| **-** | 2 | 52354 | intergenic | | C | CA | 1 | 0 | 1 |
| **-** | 2 | 260714 | intergenic | | CAAAGTTACGCAAAG | C | 1 | - | 0 |
| **-** | 2 | 603535 | intergenic | | A | G | 0 | 0 | 1 |
| HJ37_RS17585 | 2 | 731831 | nonsynonymous | Gly589Asp | G | A | 0 | 0 | 1 |
